# Quantifying inflammatory resolution in human menstruation reveals disease-specific failure modes and enables a non-invasive diagnostic for endometriosis

**DOI:** 10.64898/2025.12.30.25343168

**Authors:** Stephen Gire, Stephen Palmer, Abhishek Ja, Ridhi Tariyal

## Abstract

Inflammatory resolution is essential for tissue health, yet its dynamics remain difficult to study in humans. Menstruation is a recurrent, non-pathological inflammatory process that provides a natural window into inflammation and repair. We developed and validated a standardized menstrual sampling and RNA-seq workflow, analyzing more than 1,000 samples from over 300 individuals. We show that menstrual transcriptomes are dominated by two major biological confounders: heterogeneous tissue composition and rapid temporal progression. We introduce tissue-aware transcriptional axes that quantify uterine enrichment and an Inflammatory Resolution Score (IRS) that positions samples along an inflammation-to-repair trajectory independent of tissue admixture. In healthy individuals, IRS defines a conserved resolution trajectory across early menstruation. Applying this framework to endometriosis and autoimmune disease reveals reproducible deviations from the healthy trajectory with distinct transcriptional programs and altered pathway coordination. Finally, we demonstrate translational relevance by developing a non-invasive endometriosis classifier grounded in resolution biology that generalizes across symptomatic populations and shifts following surgical intervention. This work establishes menstruation as a tractable human model system for quantifying inflammatory resolution and detecting disease-associated disruptions.

## INTRODUCTION

Understanding inflammatory and immune-mediated diseases is limited by the lack of human model systems that allow direct observation of inflammatory initiation and resolution *in vivo*. Most molecular measurements are obtained at isolated clinical time points or from invasive procedures, providing only static snapshots of processes that are inherently dynamic^1,2^. As a result, key features of inflammatory progression, persistence, and repair remain difficult to study systematically in humans, constraining both biological insight and translational progress^3^.

Menstruation is a recurrent, non-pathological inflammatory process that involves coordinated immune activation, tissue breakdown, vascular remodeling, and repair. Each menstrual cycle represents a naturally occurring instance of acute inflammation followed by resolution, unfolding over a well-defined temporal window^4–6^. Menstrual effluence contains a mixture of shed endometrial tissue, blood, stromal and epithelial cells, immune populations, and microbial transcripts, capturing multiple layers of inflammatory biology that are difficult to access through peripheral blood or episodic biopsy^7–9^. Despite this, menstruation has rarely been treated as a quantitative model system for studying inflammatory dynamics.

A major barrier to leveraging menstrual transcriptomes has been the presence of strong biological and technical confounders that obscure interpretable signal. Menstrual effluence is compositionally heterogeneous, with variable contributions from uterine and cervicovaginal tissues that depend on bleeding phenotype, collection method, and individual physiology^10,11^. In addition, transcriptional profiles change rapidly across early menstruation, such that samples collected even one day apart reflect different biological states. Without explicit modeling of tissue composition and temporal progression, naïve analyses are dominated by these effects, masking underlying inflammatory and repair programs^12,13^.

These challenges are particularly relevant for diseases in which inflammation and immune dysregulation intersect with menstruation^14^. Endometriosis, which affects approximately one in ten individuals with a uterus, is characterized by chronic inflammation, aberrant immune responses, and altered tissue remodeling, yet diagnosis is often delayed for years and typically requires surgical confirmation^15^. Existing biomarkers lack sensitivity or specificity, and disease heterogeneity further complicates detection^16^. More broadly, conditions such as fibroids, adenomyosis, abnormal uterine bleeding, reproductive tract infections, and systemic immune-mediated diseases may similarly perturb menstrual inflammatory dynamics but remain poorly characterized at the molecular level.

A framework is needed that treats menstruation as a dynamic biological system rather than a static specimen^17^. Such a framework must enable reproducible molecular profiling at scale, explicitly account for tissue heterogeneity, and quantify progression along an inflammatory-to-repair trajectory. Here, we develop and validate a standardized menstrual sampling and RNA-seq approach and introduce tissue-aware modeling strategies that disentangle composition-driven variation from biological signal. Building on this foundation, we define a quantitative measure of inflammatory resolution that captures conserved temporal dynamics of menstrual repair and apply this framework to identify disease-associated deviations in endometriosis, and we utilize this framework to develop and validate a gene classifier for endometriosis. Together, this work establishes menstruation as an underutilized human model system for studying inflammatory resolution and provides a quantitative foundation for investigating immune-mediated disease biology.

## RESULTS

Analyses throughout the Results draw on multiple, partially overlapping datasets with prespecified inclusion criteria tailored to each biological question (see Methods).

### 1. Standardized at-home collection enables tissue-aware analysis of menstrual effluence

To study menstruation as a molecular model system under real-world conditions, we developed a standardized at-home collection and preservation workflow that supports high-quality RNA sequencing from menstrual effluence. The workflow integrates controlled tampon-based collection, immediate stabilization, ambient-temperature shipping, and centralized processing, and follows quality management practices aligned with the International Organization for Standardization (ISO). The Supplementary Methods describe the device design, stabilization chemistry, and quality procedures in detail.

Using this ISO-aligned workflow, we reproducibly profiled tampon-collected menstrual effluence under decentralized, real-world collection conditions, enabling quantitative modeling of menstrual biology. Participants collected menstrual and cervicovaginal samples in collection kits containing stabilization buffer (Norgen Biotek) and returned them by ambient shipment for centralized processing (Fig. 1a). From a subset of 601 menstrual tampons, we generated 1,135 RNA-sequenced libraries that consistently met predefined quality thresholds for downstream biological analysis (Supp. Table 3). RNA integrity remained stable for up to 14 days at ambient temperature, and unrecoverable failure rates remained below 3 percent, with biological rather than pre-analytical factors driving residual variability (Supp. Figs. 1–2). Because menstrual effluence contains both host and microbial material, we assessed background microbial signal and device-associated contamination and observed low and stable microbial signal in unused devices and preservation buffer (Supplementary Fig. 5).

**Figure 1.**
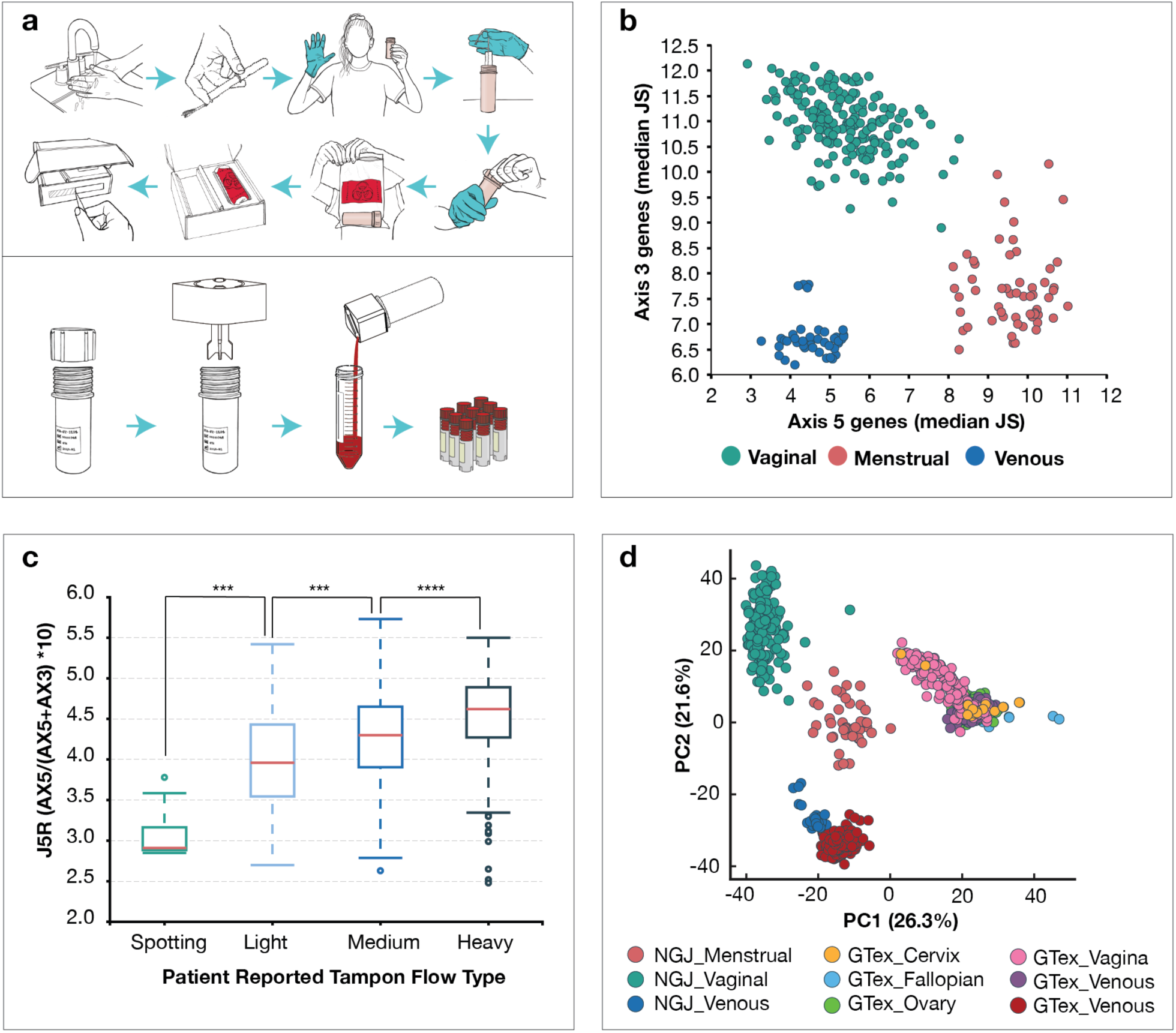
A standardized at-home menstrual collection system enables stable, tissue-aware transcriptomic profiling. **(a)** Schematic of the tampon-based, at-home collection workflow. Participants collect menstrual effluence using a standardized tampon and preservation system that releases stabilization buffer at the time of collection, enabling ambient shipment and centralized processing. Back in the laboratory, the sample is compressed, poured into a conical tube, centrifuged, and aliquoted for freezer storage. **(b)** 267 vaginal, menstrual, and whole blood specimens plotted by AX5 and AX3. AX5 and AX3 co-expression modules distinguish cervicovaginal, menstrual, and venous blood samples, defining a continuous transcriptional gradient from epithelial to uterine-enriched tissue. **(c)** Analysis of 1,135 libraries (601 tampons) stratified by J5R. The J5R ratio, derived from AX3 and AX5, scales with patient-reported tampon flow type (Global Kruskal Wallis H = 115.4; p < 1×10^−4^), supporting its use as a quantitative proxy for uterine contribution. *** = p <0.001; **** = p < 1×10^−4^). **(d)** The same 267 samples analyzed with whole blood and reproductive tissues from GTex. Analysis was performed on the tope 2,000 most variable genes. Principal component analysis situates menstrual effluence as a distinct transcriptional compartment intermediate between reproductive epithelia and blood, consistent across NGJ and GTEx reference tissues.

**Figure 2.**
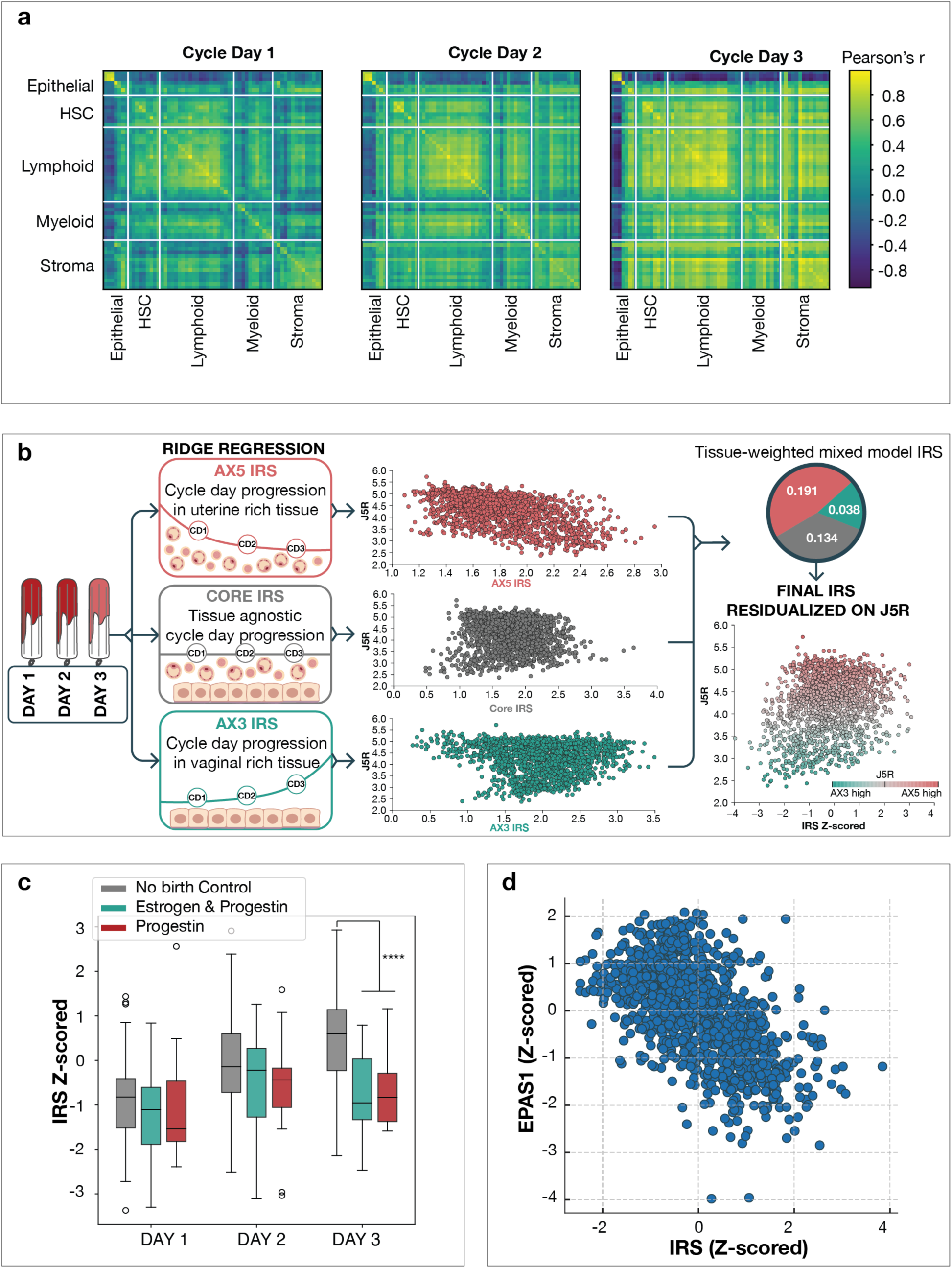
Construction and validation of a tissue-aware Inflammatory Resolution Score (IRS) **(a)** Pearson correlation between 489 cell-specific gene modules (switches) in cycle days 1-3 (10 tampons per cycle day) from 17 donors. Color scale represents correlation to all samples within a single cycle day **(b)** Conceptual framework for IRS construction. IRS development was performed on 1,116 libraries (562 tampons) from patients not on birth control. Tissue-specific ridge regression models are trained separately in uterine-enriched (AX5 IRS – 48 genes), cervicovaginal-enriched (AX3 IRS – 45 genes), and tissue-agnostic (Core IRS – 69 genes) regimes, then combined into a mixed model and residualized on J5R to remove composition effects. Tissue specific IRS models are plotted against J5R to visualize correlation with tissue axes. Weights for the combine IRS are given for AX5, AX3, and Core IRS models. Final residualized and z scored IRS is plotted against J5R. Weights for each IRS module for the overall score is given **(c)** Analysis of 158 tampons from patients with no birth control, Estrogen and Progestin, or Progestin only therapy. IRS increases monotonically from menstrual day 1 to day 3 across the full cohort of no birth control, consistent with progressive inflammatory resolution. Cycle day 3 tampons on any birth control show significant reduction in IRS compared to those not on birth control (****p < 1×10^−4^). **(d)** Data from 562 tampons from 255 patient donors (birth control samples excluded). IRS is inversely correlated with EPAS1 (HIF2α) expression r = -0.586; 95% CI -0.623 to -0.546; p < 1×10^−4^.

Menstrual effluence exhibited a reproducible, tissue-linked transcriptional structure consistent with its mixed uterine and cervicovaginal origin rather than technical degradation or noise. Two co-expression modules named AX3 and AX5 hereafter, captured the dominant sources of tissue composition across cervicovaginal, menstrual, and venous blood samples (Fig. 1b). To identify these modules, we profiled cervicovaginal, menstrual and paired menstrual phase venous blood samples from the same individuals to learn gene programs selectively enriched in each compartment, rather than relying on predefined tissue markers (Supp. Results Table 2). AX3 was enriched for epithelial programs characteristic of cervicovaginal tissue, whereas AX5 reflected uterine-derived stromal, immune, and vascular programs (Supplementary Fig. 4a. When plotted jointly, AX3 and AX5 defined a continuous transcriptional gradient rather than discrete tissue classes, enabling tissue origin to be classified with approximately 98% accuracy using these two axes alone (Fig. 1a). This structure indicates that menstrual effluence represents a coherent composite tissue state rather than nonspecific contamination.

Tissue composition could be represented quantitatively using a continuous uterine-enrichment metric, J5R, defined as AX5 divided by the sum of AX3 and AX5 and scaled by a factor of 10 to preserve log₂ interpretability. J5R scaled monotonically with patient-reported tampon flow type (Global Kruskal–Wallis H = 115.4; p < 1 × 10^−4^) Fig. 1c) and correlated strongly with established endometrial markers, including PAEP and GNLY (Supplementary Fig. 4b–c). These relationships support J5R as a quantitative proxy for uterine contribution and motivated its use downstream for sample gating, residualization, and biological interpretation rather than as a binary exclusion criterion. Thresholding behavior and sensitivity analyses for J5R are provided in Supplementary Fig. 4.

To place menstrual effluence within an external biological context, we compared its transcriptomic structure to reference tissues from the Genotype-Tissue Expression project (GTEx), including whole blood and reproductive tissue biopsies. Principal component analysis of the 2,000 most variable genes across NGJ and GTEx datasets positioned menstrual effluence as a distinct transcriptional compartment intermediate between reproductive epithelia and blood (Fig. 1d). Despite containing blood, menstrual effluence did not cluster with GTEx whole blood, instead occupying a separable space reflecting uterine, epithelial and whole blood contributions. Cervicovaginal samples aligned with GTEx cervix and vagina, and venous blood aligned with GTEx whole blood, demonstrating that menstrual effluence is transcriptionally distinct from systemic blood. This positioning was robust to normalization strategy and gene-set selection (Supplementary Fig. 3).

Together, these analyses demonstrate that tampon-collected menstrual effluence is a stable, structured, and quantifiable biospecimen whose tissue composition can be explicitly modeled rather than filtered away. This explicit modeling of tissue composition and biological context is required to distinguish temporal resolution biology from variation in sample admixture in subsequent analyses.

### 2. The Inflammatory Resolution Score (IRS) captures coordinated menstrual repair

Healthy menstruation is characterized by a monotonic, system-level cascade of coordinated cellular activity from cycle day 1 through cycle day 3, rather than by isolated changes in gene expression magnitude^18^. To define this baseline biology, we examined how cell-specific transcriptional programs co-vary across the early menstrual window in healthy individuals. Using xCell-derived gene sets^19^, expression was summarized into 489 cell-specific programs (“switches”) spanning stromal, epithelial, immune, endothelial, and hematopoietic compartments. Correlation analysis across cycle days 1–3 revealed a highly ordered increase in coordination across these programs, consistent with a progressive transition from inflammatory breakdown toward tissue repair (Fig. 2a). Cycle length per patient was not recorded.

This coordinated network structure increased monotonically across the menstrual window, defining a reproducible temporal trajectory of inflammatory resolution. Network structure was evaluated by comparing Pearson correlation-based analyses of cell specific programs (switches) between cycle days (Δr). Between cycle days 1 and 2, the median change in correlation across switch–switch correlations was Δr = 0.09, with 65% of edges showing increased coupling; this trend strengthened further between days 2 and 3 (median Δr = 0.14, 70.2% positive edges). When comparing cycle day 3 directly to cycle day 1, 79.7% of edges exhibited increased coordination, with both Wilcoxon signed-rank and sign tests rejecting the null hypothesis of no global shift (p < 1 × 10^−300^). Together, these results establish coordinated network reorganization as a defining feature of normal menstrual resolution.

However, this monotonic cascade unfolds within a biospecimen whose tissue composition differs between individuals and changes rapidly within individuals across time, exerting a strong influence on observed gene expression structure. As shown in Figure 2a and in normalization benchmarking analyses (Supp. Fig. 3d), correlations between canonical housekeeping gene expression and the AX3 and AX5 modules expression vary substantially depending on whether cervicovaginal or uterine programs dominate a given sample. Under conventional normalization approaches such as DESeq2^20^, these effects compress biologically meaningful secondary structure and introduce strong coupling between tissue composition and apparent transcriptional signals. In this context, any attempt to summarize menstrual resolution without explicitly accounting for tissue heterogeneity yields unstable and non-generalizable results.

To quantify position along the menstrual repair trajectory while addressing this heterogeneity, we developed a tissue-aware Inflammatory Resolution Score (IRS) using 562 tampons from patients not using hormonal birth control (Supp. Results Table 4). IRS construction was explicitly guided by the compositional framework established in Section 1, with separate ridge-regression models trained within uterine-enriched (AX5 IRS), cervicovaginal-enriched (AX3 IRS), and tissue-agnostic (Core IRS) regimes, followed by integration into a mixed model and residualization on J5R to remove tissue-composition effects (Fig. 2b; Supplementary Fig. 6). By constraining feature selection within biologically coherent tissue regimes and explicitly accounting for composition, the final IRS is defined by a reduced and interpretable gene set (162 genes) and exhibits improved stability and reproducibility across samples within and between individuals. This formulation yields a continuous, cycle-aware measure of inflammatory resolution that is comparable across heterogeneous menstrual effluent samples and suitable for downstream biological and clinical analyses.

Following residualization, IRS behaved as a temporal index of resolution rather than a proxy for tissue abundance. IRS showed minimal correlation with uterine-enrichment metrics (|r| ≈ 0.03 with J5R), while retaining strong cycle-day structure across the cohort. Among patients not on birth control, mean IRS increased from−0.59 on cycle day 1 to 0.15 on day 2 and 0.51 on day 3 (ANOVA F = 116.3; p ≈ 1.4 × 10^−46^; Fig. 2c), consistent with progressive inflammatory resolution. These properties indicate that IRS captures timing along a coordinated repair trajectory, not static inflammatory burden or sample composition.

IRS progression aligned with established hypoxia-regulated biology of menstrual repair, providing independent biological anchoring for the score. IRS correlated inversely with *EPAS1* (endothelial PAS domain protein 1; HIF2α), expression (r = −0.586; 95% CI −0.623 to −0.546; p < 1 × 10^−4^; Fig. 2d) and positively with *HIF1*α (hypoxia-inducible factor 1 alpha). Prior work has shown a transition from *EPAS1*-dominated states during menstrual breakdown to *HIF1*α-mediated programs during tissue repair^8^, and the observed IRS associations follow this same pattern. These results directly link IRS to known hypoxia-driven mechanisms of endometrial repair.

Hormonal suppression provided a physiologic perturbation that further validated IRS as a measure of deviation from the normal menstrual trajectory. In samples collected on cycle day 3, individuals using any form of hormonal birth control exhibited a marked reduction in IRS compared with individuals not using birth control (p < 1 × 10^−4^; Fig. 2c). Notably, this separation was largest on cycle day 3, the time point at which healthy samples show maximal progression along the resolution trajectory. These results demonstrate that IRS is sensitive to drug-induced disruption of menstrual resolution and highlight its potential use as a molecular endpoint for evaluating therapeutic effects on inflammatory repair.

Together, these analyses establish IRS as a cycle- and tissue-aware quantitative reference that captures coordinated menstrual inflammation resolution, is grounded in known hypoxia biology, and is sensitive to physiologic and pharmacologic perturbation. This framework enables rigorous comparison of resolved and unresolved states at biologically meaningful time points and provides the foundation for disease-specific analyses in subsequent sections.

### 3. Disease-specific failure modes of inflammatory resolution

Cycle day 3 marks a critical inflection point in menstruation at which coordinated inflammatory resolution should be underway in healthy individuals, providing a natural window to interrogate failures of resolution in disease. In healthy cycles, network-level coordination increases sharply between cycle days 2 and 3, whereas perturbations such as hormonal suppression blunt this transition most strongly at day 3 (Fig. 2c). We therefore hypothesized that persistent, unresolved states at this biological milestone encode disease-specific biology that interfere with subsequent resolution across the menstrual window.

To test this hypothesis at a shared biological time point, we compared transcriptional network structure between resolved and unresolved samples collected on cycle day 3 across clinical cohorts. We stratified samples using IRS to control for tissue composition and resolution timing, yielding 113 cycle day 3 tampons from 91 individuals, including 24 healthy controls, 55 endometriosis cases, and 34 autoimmune cases (Fig. 3a; Supp. Tables 4 and 6). We evaluated network structure using correlation-based analyses, in which nodes represent genes and edges represent pairwise gene–gene coexpression relationships (Fig. 3b). We quantified differences between resolved and persistent/unresolved states using ΔZ, defined as the absolute change in correlation strength for each edge to capture the magnitude of network rewiring, and Δρ, defined as the signed change in mean correlation, to capture the direction of rewiring toward unresolved or resolved states.

**Figure 3.**
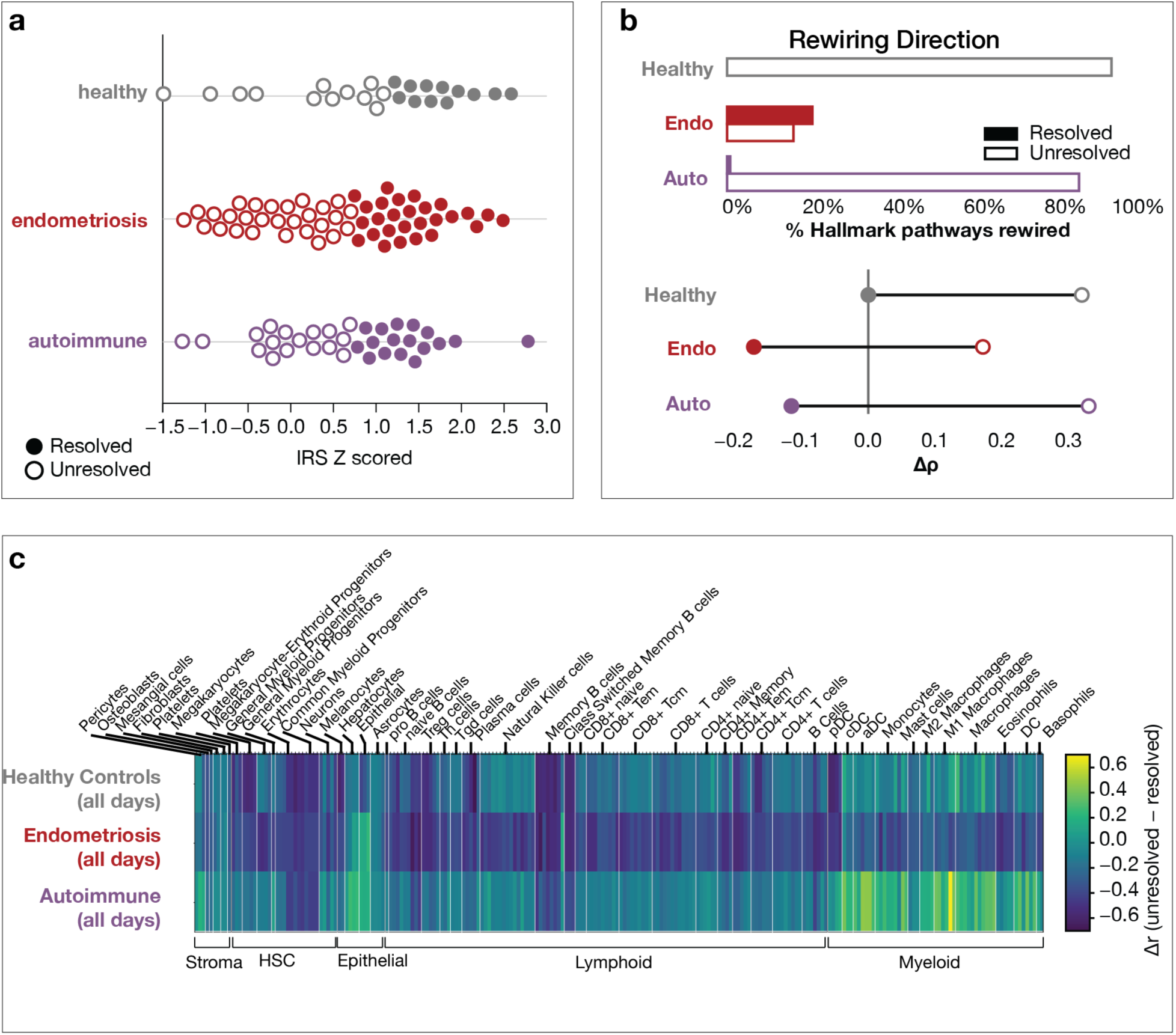
Unresolved inflammatory states exhibit disease-specific transcriptional rewiring architectures. **(a)** IRS distributions across 113 cycle day 3 tampons from 91 individuals: healthy controls (24 tampons), endometriosis (55 tampons), and autoimmune disease (34 tampons). Resolved vs. unresolved (median IRS for each cohort) tampons are highlighted by filled in or empty circles. **(b)** Cycle day 3 analysis of Hallmark network rewiring in unresolved samples, showing the extent of pathway rewiring and the direction toward resolved or unresolved phenotypes. **(e)** Analysis of 486 tampons from 208 patients (Healthy – 37; endometriosis – 114; autoimmune – 57). Cycle days 1-3 total IRS trajectory analysis of cell specific activity. Delta r correlation shifts from resolved to unresolved for endothelial cell programs compared to other cell specific activity by disease group. Positive delta R means the correlation is stronger in unresolved states. Negative delta r means correlations are stronger in resolved states.

At the level of global network organization, persistent/unresolved samples showed quantitative differences in the extent of transcriptional rewiring across disease cohorts. Autoimmune samples exhibited the greatest reorganization burden, with 81 rewired hub genes and ∼3,600 altered edges (∼18% of the inferred interaction network). Endometriosis samples showed a more constrained pattern with 20 hubs and ∼2,650 edges (∼13%), whereas healthy samples exhibited fewer perturbations, with 10 hubs and ∼2,000 edges (∼10%) (Fig. 3a,b). These node and edge counts were identified at FDR < 0.1 and are sensitive to cohort size; accordingly, we interpret these values as descriptive measures of network reorganization burden rather than definitive biological signatures. Full hub identities, edge distributions, and ΔZ summaries are provided in Supplementary Fig. 7. These edges can be found in supplemental data.

Biological coherence emerged when we examined network rewiring at the pathway level. Aggregating ΔZ and Δρ across 53 HALLMARK pathway sets revealed widespread pathway engagement in persistent/unresolved states, with distinct disease-dependent patterns (Supplementary Fig. 7). In autoimmune samples, most pathways showed broad reinforcement, with a mean of ∼92% of genes per pathway exceeding a moderate rewiring threshold and positive Δρ values, consistent with persistent immune, interferon, and antigen-presentation programs. In contrast, endometriosis samples showed more selective pathway involvement, with a lower mean fraction of rewired genes (∼80%) and reinforcement concentrated in extracellular matrix remodeling and angiogenic pathways. These differences indicate that disease specificity arises from which biological programs are reinforced and how they are reinforced, rather than from the mere presence of rewiring.

Analyses restricted to cycle day 3, however, cannot distinguish transient delay from persistent failure of resolution across menstruation. Because IRS provides a continuous, tissue-aware measure of progression independent of calendar cycle day, we next examined whether disease-specific coordination patterns persist across the full resolution trajectory. Using 486 tampons from 208 patients spanning cycle days 1–3 (only healthy, endometriosis, and autoimmune patient samples were included in this analysis), we analyzed how endothelial cell programs coupled to other cell-specific activity states as samples progressed from low to high IRS (Fig. 3e).

This analysis revealed distinct disease-specific patterns of endothelial remodeling. In autoimmune disease, resolution coincided with active uncoupling of endothelial programs from antigen-presenting cell activity . In low-IRS autoimmune samples, endothelial programs showed strong coupling to dendritic cell and related antigen-presenting states (Δr > 0.3), whereas these correlations collapsed toward zero at high IRS, indicating coordinated disengagement as resolution progressed (Fig. 3e). In contrast, endometriosis samples showed persistent endothelial coupling to epithelial and stromal programs across the IRS trajectory (Δr > 0.2–0.3), with positive correlations remaining even at high IRS. Healthy samples exhibited minimal endothelial coupling shifts across IRS strata, with correlations remaining near zero, consistent with stable coordination during successful resolution.

Together, these analyses demonstrate that inflammatory resolution fails through distinct, disease-specific network architectures that can be detected both at a fixed biological milestone and across the continuous resolution trajectory. By integrating cycle day 3 rewiring with IRS-based endothelial coupling across the menstrual window, this framework distinguishes immune-dominant failure in autoimmune disease from tissue-centric persistence in endometriosis while accounting for tissue heterogeneity, temporal confounding, and statistical uncertainty.

### 4. Translation of inflammation biology profiling into a non-invasive endometriosis diagnostic

The resolution-aware framework established above provides a foundation for translating menstrual biology into a practical, non-invasive diagnostic context. By explicitly modeling tissue composition, temporal progression, and disease-specific failure modes of inflammatory resolution, Sections 1–3 define biologically grounded constraints that can be evaluated for their ability to support disease discrimination for endometriosis in clinically relevant populations.

We therefore grounded feature discovery for diagnostic modeling in resolution biology and menstrual context rather than applying unconstrained machine learning. We selected candidate features based on association with endometriosis status and reduced correlation with known menstrual covariates, including cycle day, tissue composition axes, and bleeding-related metrics, to minimize confounding by physiologic variation. We restricted discovery analyses to cycle days 2–3, the interval showing the most stable separation between resolved and persistent/unresolved states, and to uterine-enriched samples defined by the J5R axis. The discovery cohort comprised 136 tampons from 46 patients, with multiple tampons per patient retained to increase power given a measured intraclass correlation coefficient (ICC) of 0.25 (Fig. 4a). The final feature set included 19 molecular features spanning host transcription, microbial activity, and tissue composition, together with one categorical bleeding phenotype, selected for biological relevance and stability.

**Figure 4.**
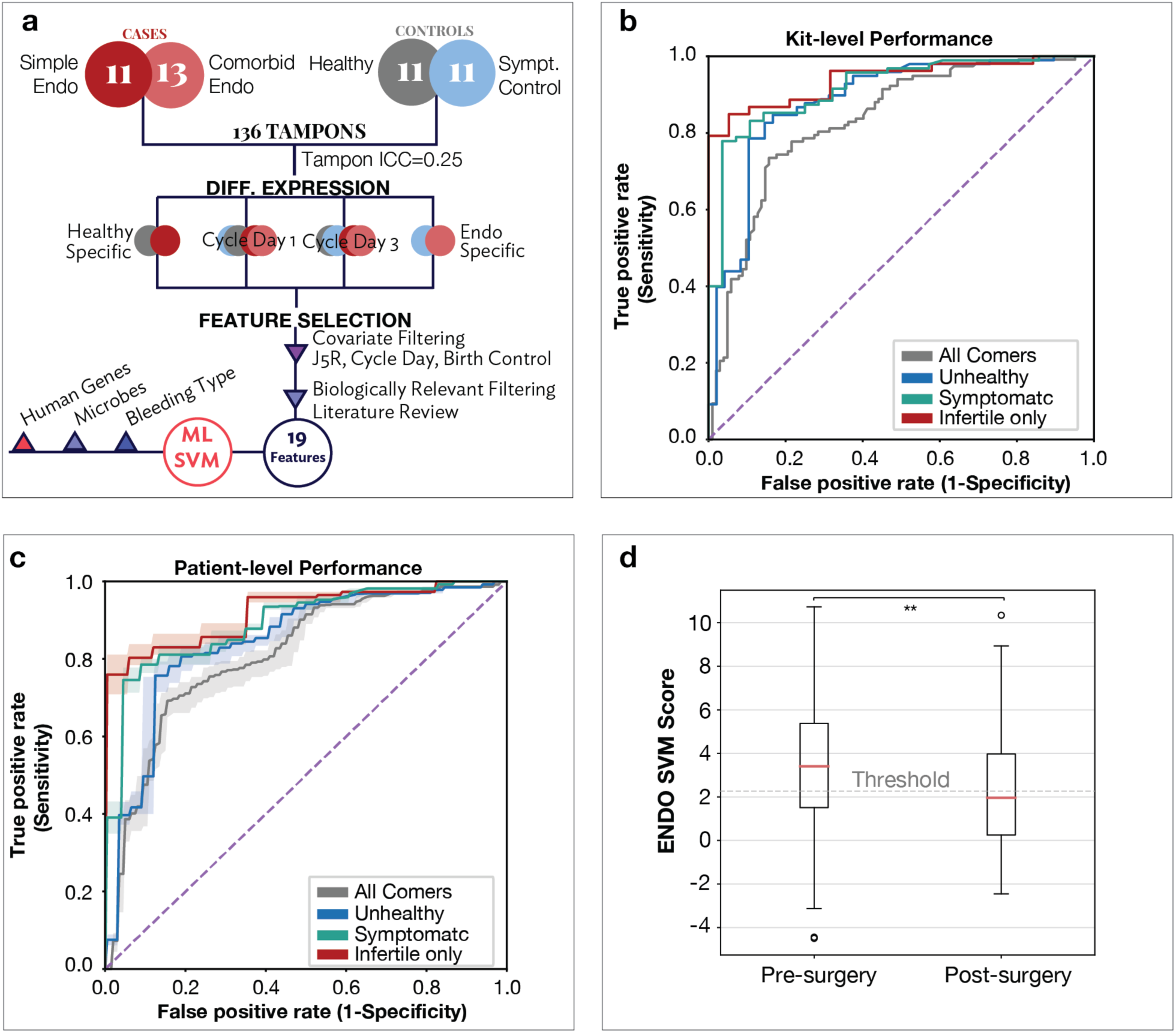
Translation of resolution-aware biology into a non-invasive endometriosis diagnostic. **(a)** Feature discovery and classifier development pipeline. Discovery and training analysis performed on 136 tampons from 46 individuals (Healthy – 11; Symptomatic controls – 11; endometriosis with no comorbidities – 11; and endo with 1 or more comorbidities – 13). Interclass coefficient between tampons collected on separate days is 0.25, demonstrated value in including multiple tampons from the same individual in discovery and training. Differential expression, biologically informed filtering, and covariate control yield a 19-feature molecular panel (host transcription, microbial activity, tissue composition), together with a categorical bleeding phenotype variable incorporated as a clinical covariate, trained using a linear support vector machine. **(b)** Kit level validation of endometriosis classifier on independent cohort of patients (1 tampon per patient validation). Receiver operater characteristic curves demonstrate robust diagnostic performance in independent validation cohorts, including all-comers (gray, AUC 0.83), unhealthy patients (blue, AUC 0.88), symptomatic patients (teal, AUC 0.92) and infertility-only patients (red, AUC 0.94). **(c)** Patient level validation of endometriosis classifier on independent cohort of patients (1 tampon per patient validation). Receiver operater characteristic curves demonstrate robust diagnostic performance in independent validation cohorts, including all-comers (gray, 139 patients, AUC 0.81), unhealthy patients (blue, 97 patients, AUC 0.84), symptomatic patients (teal, 78 patients, AUC 0.89) and infertility-only patients (red, 54 patients, AUC 0.92). 95% CI present for each as shaded area. Breakdown of cases and controls, patient metadata and comorbidities can be found in Supplemental Table 6 **(d)** Analysis of ENDO SVM classifier in Pre- and Post-Surgery cohorts. Median score for Pre (3.406) vs. Post (1.962), shows a significant (MWU p = 0.006) reduction in ENDO score post-surgery.

We designed model training and evaluation to prioritize generalizability over apparent training performance. We trained a linear support vector machine (SVM) classifier with strict patient-level separation to prevent information leakage, and we derived all reported performance metrics from independent validation cohorts set aside for testing and not utilized in discovery or model fitting. We performed validation at the kit level and the patient level to assess the analytical and clinical validity of the test. The primary validation cohort included 139 patients (all-comers), with prespecified subgroups of 97 unhealthy patients (at least one disease known to cause menstrual disruption), 78 symptomatic patients (infertility and/or chronic pelvic pain), and 54 infertility patients, enabling evaluation in clinically ambiguous and high-risk populations rather than healthy screening settings (Fig. 4b; Supplementary Fig. 8f). The definition and composition of these cohorts can be found Supp. Table 6B.

Across cohorts set aside for validation, the classifier demonstrated consistent and biologically meaningful gains in discriminative performance. In the all-comers validation cohort of 139 patients, the model achieved an AUC of 0.83 at the kit level, with improved discrimination in unhealthy patients (AUC 0.88) and the strongest performance in symptomatic (AUC 0.92) and infertile patients (AUC 0.94) (Fig. 4b). Importantly, these increases in AUC were accompanied by corresponding increases in Matthew’s correlation coefficient (MCC), ranging from approximately 0.65 to 0.73, indicating that improved performance reflects genuine gains in balanced classification rather than threshold-dependent effects. Operating points were selected to emphasize specificity (sensitivity 84.9% and specificity 94.7%; Supp. Fig. 8e), consistent with the intended use of the test as a diagnostic aid in symptomatic and infertility populations rather than as a population-level screening assay.

To assess expected clinical performance with a single specimen per individual, we evaluated classifier accuracy at the patient level by randomly selecting one tampon per patient and averaging performance across ten iterations (Fig. 4c). Across all comers, patient-level discrimination remained robust, with a mean AUC of 0.812 (95% CI 0.79–0.83) and an MCC of 0.554 (95% CI 0.51–0.58), representing only modest attenuation relative to kit-level performance (AUC 0.83). Performance improved with clinical enrichment: patient-level AUC reached 0.844 (95% CI 0.83–0.86) in the unhealthy cohort, 0.893 (95% CI 0.88–0.90) among symptomatic patients, and 0.922 (95% CI 0.91–0.94) in the infertility subgroup (54 patients), with corresponding MCCs of 0.604, 0.648, and 0.712, respectively. These results demonstrate that the classifier maintains strong discriminative power when applied at the patient level using a single menstrual sample, supporting its clinical applicability.

The diagnostic markers retained in the final classifier align with the resolution biology described in Sections 2 and 3 rather than representing arbitrary correlates of disease status. Host transcriptional features included genes involved in immune signaling, cellular stress responses, and tissue remodeling, processes that are dynamically regulated during menstrual breakdown and repair. For example, *BCL6* (B cell lymphoma 6), a transcriptional regulator of immune activation and progesterone resistance, reflects persistence of inflammatory and immune programs that fail to disengage during resolution in endometriosis. Similarly, *RGS16* (regulator of G protein signaling 16), which modulates G protein–coupled receptor signaling and immune cell trafficking, maps onto the immune coordination and disengagement patterns identified in the network analyses. Together with additional metabolic and mitochondrial markers, these features capture coordinated resolution dynamics rather than static inflammatory burden.

Microbial features included in the classifier should be interpreted as contextual components of the menstrual ecosystem rather than direct diagnostic drivers of endometriosis. Menstrual microbial activity fluctuates with tissue shedding, immune exposure, and repair, and microbial transcripts contribute information about the local inflammatory milieu without implying causality. Their modest but consistent contribution to classification reflects this contextual role and reinforces the importance of modeling menstrual biology as an integrated host–microbial system rather than filtering microbial signal entirely.

To assess robustness beyond performance metrics, we examined classifier behavior under orthogonal perturbations. In an unpaired, population-level comparison including 127 pre-surgery and 46 post-surgery samples (Median 5.5 months post-excision; IQR 2-24 months), ENDO SVM scores were significantly lower following surgery, with median scores decreasing from 3.406 pre-surgery to 1.962 post-surgery (Mann–Whitney U p = 0.006) (Fig. 4e). Although this analysis does not establish within-patient causality, the direction and magnitude of the shift provide independent support that the classifier reflects disease-associated molecular states rather than static patient characteristics.

Additional analyses assessed performance under increased biological heterogeneity and generalizability in clinically realistic populations. Performance decreased in cycle day 1 samples, consistent with greater heterogeneity during early inflammatory breakdown (Supplementary Fig. 8a–b). Post hoc nested cross-validation across the full dataset showed stable performance with minimal variance across folds, providing secondary evidence against overfitting (Supplementary Fig. 8c). The validation cohort included comorbidity structure, including adenomyosis, fibroids, PCOS, and autoimmune disease, as shown in Supplementary Table 6. Inclusion of symptomatic controls during discovery and training reinforces that the classifier aims to resolve endometriosis within a clinically realistic population rather than to distinguish disease from health.

Together, these results demonstrate that resolution-aware menstrual biology can support non-invasive disease discrimination across heterogeneous, symptomatic patient populations. By grounding feature selection in menstrual context, demonstrating concordant gains in AUC and MCC, validating behavior under surgical perturbation, and testing performance under conditions of increased biological noise, this framework supports translational relevance while avoiding claims of population screening utility. Broader implications for clinical integration are considered in the Discussion.

## DISCUSSION

This work reframes menstruation as a quantitative, tissue- and cycle-aware human model of inflammatory resolution and repair. Rather than treating menstrual effluence as a confounded biospecimen, we show that its heterogeneity can be explicitly modeled to reveal coordinated biological programs that are otherwise difficult to observe in humans. By integrating tissue composition, temporal progression, and network-level coordination, this framework shifts the focus from static inflammatory markers toward resolution as a measurable, dynamic process.

A central conceptual advance of this study is the distinction between inflammatory burden and inflammatory resolution. Many molecular approaches implicitly equate inflammation with magnitude, emphasizing elevated markers or differential expression at isolated time points^21^. In contrast, our results indicate that healthy menstruation is defined by a monotonic increase in coordinated cellular signaling across the early menstrual window, reflecting orderly progression through breakdown, clearance, and repair. The IRS metric was designed to capture this progression as a temporal index of coordinated repair rather than an intensity metric. Its alignment with hypoxia-regulated transitions, including the opposing behavior of *EPAS1* and *HIF1*α, situates this score within established endometrial repair biology^8^ while enabling quantitative assessment of resolution timing at scale.

Applying this resolution-aware framework to disease reveals that failure of resolution is not uniform but instead reflects distinct biological architectures. Endometriosis and autoimmune disease diverge not simply in the extent of network rewiring, but in the logic of the programs that remain engaged. Endometriosis is characterized by persistence of epithelial–stromal and angiogenic coordination, consistent with tissue-centric failure to disengage repair processes. Autoimmune disease, by contrast, shows immune-dominant persistence, with broad engagement of interferon and antigen-presentation pathways. These differences underscore that resolution failure encodes disease-specific states rather than generic inflammatory delay.

The endothelial compartment emerges as a particularly important integrator of resolution biology. Trajectory analysis shows that successful resolution in autoimmune disease involves active uncoupling of endothelial programs from antigen-presenting cell activity, suggesting that disengagement of immune–vascular crosstalk is a necessary step in repair. This uncoupling is absent in endometriosis, where endothelial coordination with epithelial and stromal programs persists, consistent with ongoing remodeling rather than immune disengagement. These findings highlight repair biology as a process of selective disengagement, not simply attenuation of inflammation, and point to vascular–immune interactions as a key axis of failure in immune-mediated disease^22,23^.

An important nuance that cannot be fully explored in the Results is the role of microbial signal within menstrual effluence. Bacterial transcripts are included in this framework not as diagnostic features of endometriosis per se, but as contextual components of the menstrual ecosystem^24^. Menstruation is not a sterile process, and microbial activity fluctuates alongside tissue breakdown and repair^25^ (Supp. Fig. 5). By modeling microbial signal in concert with host transcription rather than filtering it out, we preserve biologically meaningful context while avoiding spurious associations driven by contamination or low-biomass artifacts. In this framework, microbial features contribute modestly and contextually, reinforcing that they serve as ecological markers rather than disease drivers.

The translational analyses illustrate how resolution-aware menstrual biology can be applied without decoupling diagnostics from underlying biology. By constraining feature selection to markers with reduced correlation to menstrual covariates and grounding model development in tissue- and cycle-aware principles, we demonstrate that a non-invasive diagnostic signal can generalize across heterogeneous, symptomatic populations. Concordant increases in area under the curve and Matthew’s correlation coefficient across validation cohorts indicate genuine gains in balanced classification rather than threshold effects. Alignment of diagnostic scores with resolution biology, despite independent training, supports shared biological underpinnings rather than circularity, while reduced performance on cycle day 1 reinforces the importance of biological context and intended use.

Several limitations warrant consideration and point toward important future directions. Menstrual effluence is inherently heterogeneous, and although explicit modeling of tissue composition and temporal progression mitigates this challenge, residual variability remains, particularly at early cycle days when inflammatory breakdown dominates. Larger disease-specific cohorts will be required to resolve finer-grained subtypes and to extend this framework beyond endometriosis and autoimmune disease. Longitudinal within-patient sampling across multiple cycles will be critical for distinguishing stable failure modes from state-dependent fluctuations. Hormonal contraceptive users were underrepresented, and the observed suppression of IRS under hormonal exposure suggests that tailored models may be required for these populations. Endometriosis endophenotypes (endometrioma, deep infiltrating, or peritoneal) were not assessed in this study. While some operative reports assessed overall stage and subtype of disease, many operative reports did not, making endophenotypic beyond presence or absence difficult. While a single, low-absorbency junior tampon was well tolerated within our cohort, other collection modalities that may better suit patients with vulvodynia or other conditions that may make tampon use difficult, should be considered in future studies. Prospective interventional studies will also be necessary to determine whether modulation of resolution biology, as measured by resolution-aware metrics, predicts clinical outcomes.

In summary, this work introduces a menstrual framework that makes inflammatory resolution measurable in humans and reveals how repair fails across disease contexts. By treating menstruation as a structured biological process rather than a confounder, this approach unlocks new opportunities to study human repair biology, develop resolution-aware diagnostics, and inform therapeutic strategies aimed not merely at suppressing inflammation, but at restoring coordinated repair.

## Supporting information

Supplemental Results

AX3 and AX5 reactome analysis

Autoimmune CD3 top edges

Autoimmune CD3 top nodes

Endometriosis CD3 top edges

Endometriosis CD3 top nodes

Healthy CD3 top edges0

Healthy CD3 top nodes

## Acknowledgements

We would like to thank Elucidata Inc., Xitong Li, and Dylan Kotliar for bioinformatic support. Margaret Eisen, Julia Carr and Aparna Kola coordinated all clinical collection efforts for this study. Kit IFU artwork was done by Sarah Bush. The analyses in this paper were partly funded through two SBIR grants and one R01 subaward grant through the National Institute of Child Health and Human Development 5R44HD103159 and R44HD118899, and 1R01HD099341, respectively.

## Author contributions

Stephen Gire: Conceptualization, Methodology, Validation, Formal Analysis, Investigation, Resources, Writing, Editing, Visualization, Supervision, Funding Acquisition. Ridhi Tariyal: Conceptualization, Methodology, Resources, Data Curation, Writing, Editing, Supervision, Project administration, Funding acquisition. Stephen Palmer: Conceptualization, Resources, Editing, Funding Acquisition, Abhishek Ja: Methodology, Validation, Formal Analysis, Resources, Supervision.

## Data availability

Data summaries of patient and kit level metadata and sequencing data are available in the supplemental. The sequencing datasets generated and analyzed during the current study are not publicly available due to their proprietary nature and potential for intellectual property development. These data comprise a novel resource that may enable multiple future patent filings. As such, access to the raw sequencing data will only be considered in the context of a formal collaboration agreement. Interested researchers may contact the corresponding author to discuss potential collaborative opportunities and access procedures.

## METHODS

A detailed methods section is available in the supplemental. A breakdown of patient samples, comorbidities, tampons and cycle days are provided in supplemental Tables. In total, 1,718 libraries from 1,004 tampons collected from 342 patients were included in this study, spanning various analyses. Supplemental Tables outline the samples, kits, and patients used for each main analysis in this paper.

Analyses draw on multiple, partially overlapping datasets assembled to address distinct biological and translational questions. Because menstrual effluence is a heterogeneous biospecimen and individuals may contribute multiple tampons across cycle days or clinical contexts, inclusion criteria and units of analysis were prespecified and vary by result. Tissue composition gene modules (AX3, AX5) and the uterine enrichment metric (J5R) were derived from an internal calibration dataset with matched cervicovaginal, menstrual, and venous blood samples, enabling compartment-specific modeling and alignment to GTEx.

Downstream analyses applied biologically motivated filters, including sequencing quality thresholds, restriction to cycle days 1–3, and clinically defined disease status. Endometriosis diagnostic development used strict case–control definitions, excluded post-surgical samples from discovery, training, and validation, and prioritized independent validation cohort size to support generalizability. Diagnostic performance is reported at both the kit and patient levels.

### Study design and participant enrollment

Participants were enrolled under WIRB-approved protocols (20192619, 20191947, 20233438) and provided informed consent for tampon-based menstrual effluence collection and clinical metadata reporting. Enrollment occurred through decentralized at-home and clinic-based workflows, enabling longitudinal sampling across menstrual cycles. Metadata were collected using a hierarchical structure spanning patient, cycle, kit, and sample levels, including age, geography, menstrual cycle day, hormonal birth control use, bleeding phenotype, tampon wear time, and sequencing quality metrics. This structure enabled explicit modeling of temporal progression and tissue composition while accounting for repeated measures contributed by individual participants.

### Standardized tampon-based collection and ambient stabilization

All menstrual samples were collected using a standardized tampon-based kit manufactured under an ISO 13485 quality system. Participants were instructed to wear a low-absorbency organic cotton tampon for four hours on menstrual cycle days 1 through 5, and for 30 minutes if collected outside of menstruation. Following collection, the tampon was sealed within a collection jar that automatically released a nucleic-acid stabilization buffer, preserving RNA and microbial transcripts at ambient temperature during mail transit. Upon laboratory receipt, samples were extruded, centrifuged to remove debris, and aliquoted for downstream molecular processing. RNA integrity was assessed using the DegNorm-derived DI25 metric to quantify transcript degradation^26^.

### RNA sequencing and preprocessing

Total RNA was extracted using column-based (Norgen Biotek) or magnetic bead-based protocols (ThermoFisher miRvana total RNA) incorporating Turbo DNase treatment and bead-based cleanup. Libraries were prepared using Zymo Research ribofree RNA library preparation kit. This kit utilizes a double stranded DNA nuclease to reduce overabundant transcripts. This includes ribosomal RNA, hemoglobin, or other abundant species that are sample specific. Samples were standardardized to 500ng in 9ul aliquots for input into kit. Libraries were amplified for 12 cycles, cleaned, quantified, and run on an Illumina NextSeq Kit, generating 100bp paired-end reads. Sequencing quality thresholds were enforced uniformly, including minimum strandedness, ribosomal RNA content, uniquely mapped read counts Y chromosome transcript amount. Reads were adapter-trimmed, aligned to the human reference genome using STAR aligner^27^. Reads were then normalized for degradation using three reference menstrual samples representing high quality material through Degradation Normalization^26^. Raw counts were RKPM transformed and put into JaneScore (Supp. Materials).

### Axes and Cell-specific switch construction

Pathway specific gene modules (Axes) and cell-specific transcriptional programs (Switches) were calculated by taking the median of the gene set per program. Switches were defined using xCell gene sets^19^, which digitally portray tissue cellular heterogeneity across immune, stromal, epithelial, endothelial, and hematopoietic compartments. Axes were defined by coexpression and pathway analyses exercises. This yielded 489 cell-specific switches used for coordination and coupling analyses. 10 axes are also calculated. AX3 and AX5 are presented here.

### Cell-specific coordination analysis across cycle days

Healthy reference samples were used to characterize normative changes in coordinated cell-specific signaling across cycle days 1-3. For each cycle day analyzed, ten tampon samples were selected to construct a reference correlation network from patients with clear cycle day shifts in hypoxic programs. Pairwise Pearson correlations were computed across all 489 switches, producing cycle day–specific correlation matrices. Differences in correlation strength between cycle days were quantified as Δr values across all unique switch–switch edges. Global shifts were summarized using median Δr, interquartile range, and the fraction of edges with positive Δr. Statistical significance was assessed using Wilcoxon signed-rank and sign tests applied to edge-wise Δr distributions.

### Inflammatory Resolution Score construction

The Inflammatory Resolution Score (IRS) was designed to quantify progression along the menstrual repair trajectory independent of tissue composition. Resolution-associated gene and switch features were identified based on cycle-day dependence and minimal correlation with tissue composition axes. Separate ridge regression models were trained within uterine-enriched, cervicovaginal-enriched, and tissue-agnostic regimes to capture complementary resolution programs. These components were integrated into a composite score and residualized on uterine enrichment (J5R) to remove composition effects. The final IRS was standardized to enable comparison across samples and cohorts.

### IRS stratification and hormonal birth control analysis

IRS values were examined as a function of cycle day and hormonal birth control exposure. Analyses were conducted at the kit level to avoid pseudo replication. Birth control status was derived from participant-reported metadata and categorized as no birth control, estrogen–progesterone, or progestin-only formulations. Group comparisons were performed using nonparametric tests, and regression analyses incorporated patient-clustered robust standard errors.

### Disease cohort definition and resolved–unresolved stratification

Disease cohorts were defined based on clinical metadata and included healthy controls, endometriosis cases, autoimmune cases, and comorbid presentations. For cycle day–specific analyses, resolved and unresolved states were defined using IRS thresholds within each cohort. For trajectory-based analyses, IRS was treated as a continuous variable across all cycle days.

### Endothelial coupling analysis across the resolution trajectory

Endothelial activity was summarized at the kit level by averaging z-scored endothelial switch activities. Coupling between endothelial activity and other cell-specific switches was quantified using Pearson correlation, stratified by low-IRS and high-IRS states within each disease cohort. Differences in correlation coefficients were assessed using Fisher’s z transformation, and multiple testing correction was performed using the Benjamini–Hochberg procedure. Switches meeting a false discovery rate threshold were considered differentially coupled.

### Diagnostic classifier development and evaluation

A linear support vector machine classifier was trained to detect endometriosis using resolution-aware molecular features, including selected gene expression markers, microbial taxa, tissue composition metrics, and bleeding phenotype. Model training employed strict patient-level segregation to prevent information leakage. Performance was evaluated in independent validation cohorts, and classifier scores were examined in pre- and post-surgical samples using unpaired statistical comparisons. All feature selection and model specification decisions were finalized prior to evaluation in independent validation cohorts; post hoc nested cross-validation was performed solely to estimate generalizability and did not inform model training or feature selection.

### Statistical analysis

All statistical analyses were performed using Python. Nonparametric tests were used where distributional assumptions were violated. Multiple hypothesis testing was controlled using false discovery rate correction as appropriate. Effect sizes and confidence intervals were reported alongside p values to support quantitative interpretation. Gene-level expression differences between various cohorts were assessed using two-sided Welch t-tests on log2-transformed expression values. Resulting p-values were adjusted for multiple hypothesis testing using the Benjamini–Hochberg false discovery rate procedure and reported as continuous measures of statistical support.

---

**Supplemental Figure 1.**
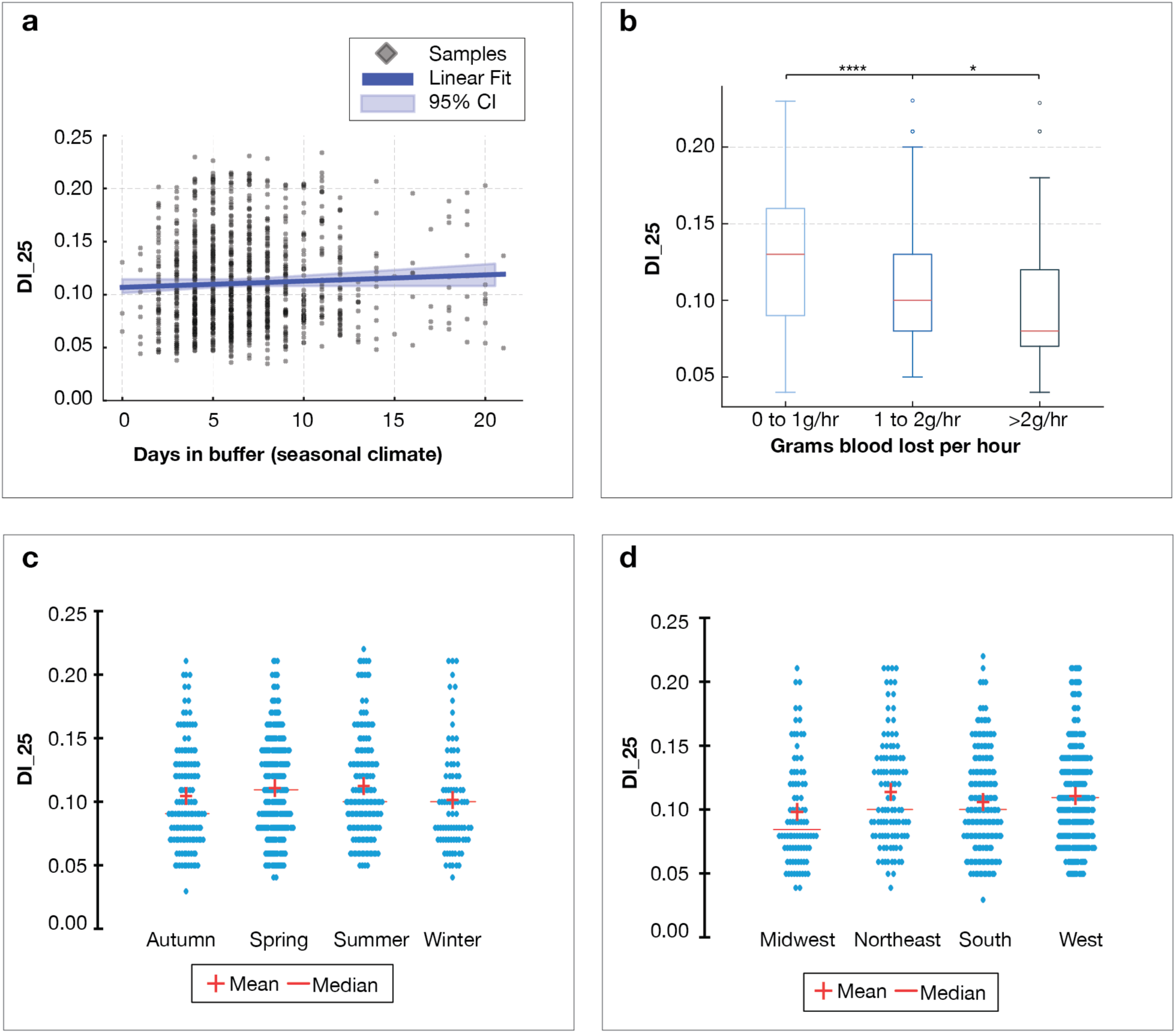
Analysis of technical and biological contributions to degradation. **(a)** Analysis of 1,135 sequencing libraries. RNA integrity, measured by the DegNorm-derived DI_25 metric, remains stable with no significant difference across days spent in buffer (ANOVA p = 0.107) under real-world seasonal conditions while varying systematically with menstrual flow rate. **(b)** Analysis of 241 tampons with recorded blood volume. DI_25 by grams of menstrual blood loss per hour. Samples with higher flow rates show lower degradation, indicating that biological shedding dynamics, rather than shipping time, dominate RNA quality. **(c) & (d)** Analysis of 1,135 sequencing libraries. DI_25 plotted by season (U.S) and by region (U.S.) the collection occurred. Midwest collected samples showed a lower mean DI_25 compared to other regions (p = 0.023; cohen’s f = 0.127).

**Supplemental Figure 2.**
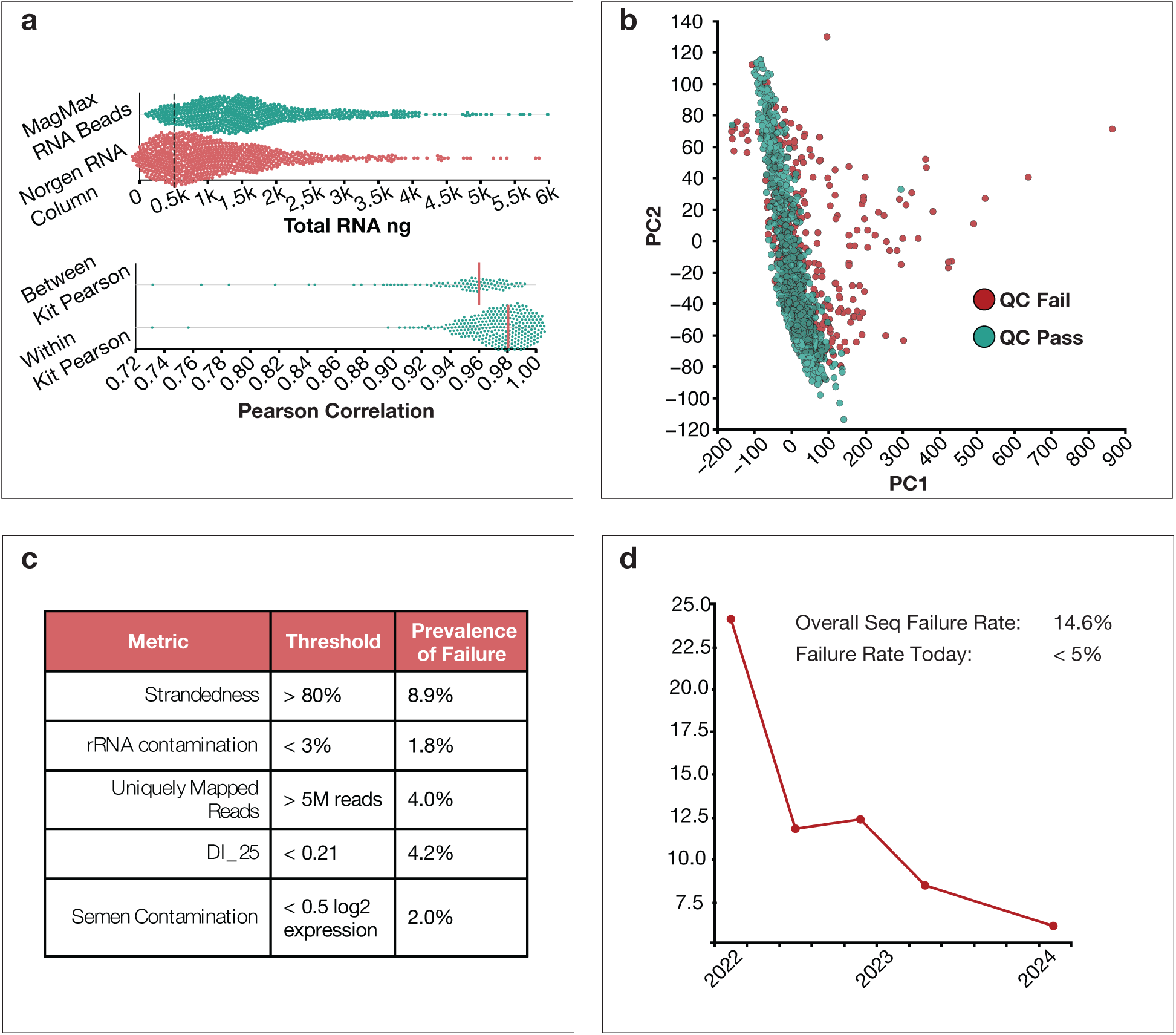
Platform reproducibility metrics and failure rates. **(a)** Analysis of 1,135 sequencing libraries passing filter. Top panel represents total RNA extracted from a single aliquot of lysate (mixture of menstrual blood to preservation buffer). Magnet beads outperform RNA columns, especially in recovery of fragmented RNA common in exfoliated sample types. Dotted line represents the 500ng threshold, the high end required amount for zymo ribofree library preparation. Kits. The bottom panel shows both within kit and between kit person correlations, demonstrating high replicate correlation and high between sample correlation for samples collected from the same individual on the same day. **(b)** PC1 and PC2 analysis of 2,229 seq libraries (representing samples outside of menstruation or not in the 1-3 day window, and all failed samples. Dots in red represent samples that fail QC thresholds presented in panel c. PC1 carries the majority of technical variability when failed samples are included in the PCA analysis. **(c)** The QC metrics used to determine pass/fail of samples, and the percent of 2,229 samples that fail on each criteria. **(d)** Failure rate of sequencing over time. Dramatic improvement of failure rate over time of 2,229 sequence libraries. Dramatic improvement are punctuated by improvement of DNase procedures, movement of extractions to magnetic beads, and replicate based sequencing.

**Supplemental Figure 3.**
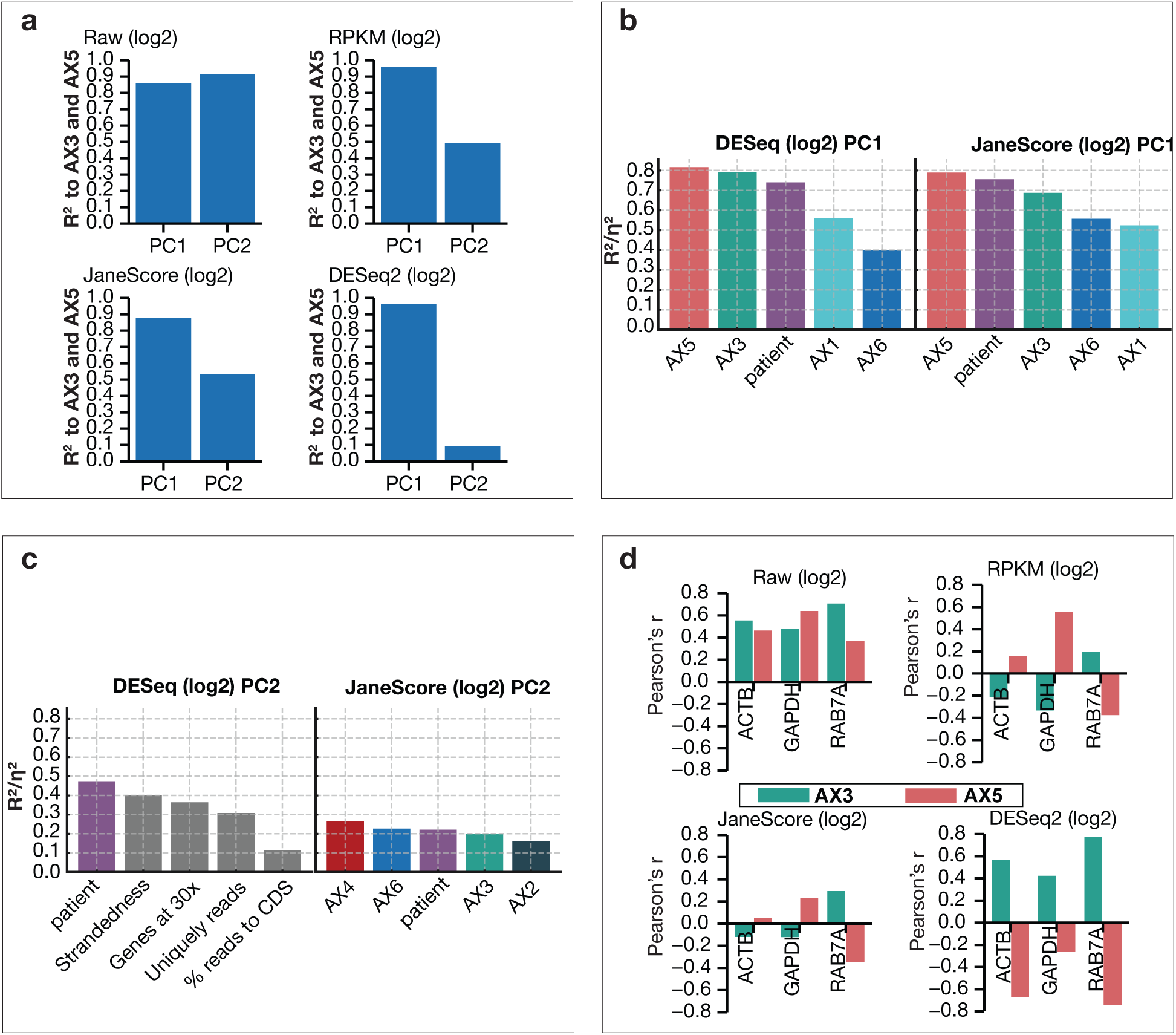
Gene normalization benchmarking. Analysis of 48 tampon samples comparing raw counts, RPKM, DESeq2 and JaneScore normalization procedures. **(a)** PCA analysis of 48 tampon sequences and the PC1 and PC2 R-squared value to AX3 and AX5 tissue modules. RPKM and JaneScore perform similarly, where DESeq2 retains most tissue relevant information to PC1 only. **(b).** Top 5 contributors to PC1 between DESeq2 and JaneScore, show similar makeup and magnitude of contributors. AX1 and AX6 are additional gene modules that represent hemoglobin genes, and estrogen receptor and signaling genes. **(c)** Top 5 contributors to PC2 between DESeq2 and JaneScore, show similar makeup and magnitude of contributors. AX2 and AX4 are additional gene modules that represent mitochondrial genes, and extracellular matrix remodling genes. (d) Correlation of housekeeping genes post normalization. JaneScore shows the least correlation to AX3 and AX5 modules post normalization. DESeq2 shows a change in correlation of housekeeping genes post normalization, demonstrating that DESeq2 is preferentially normalizing to a single homogenous tissue source instead of tissue admixture.

**Supplemental Figure 4.**
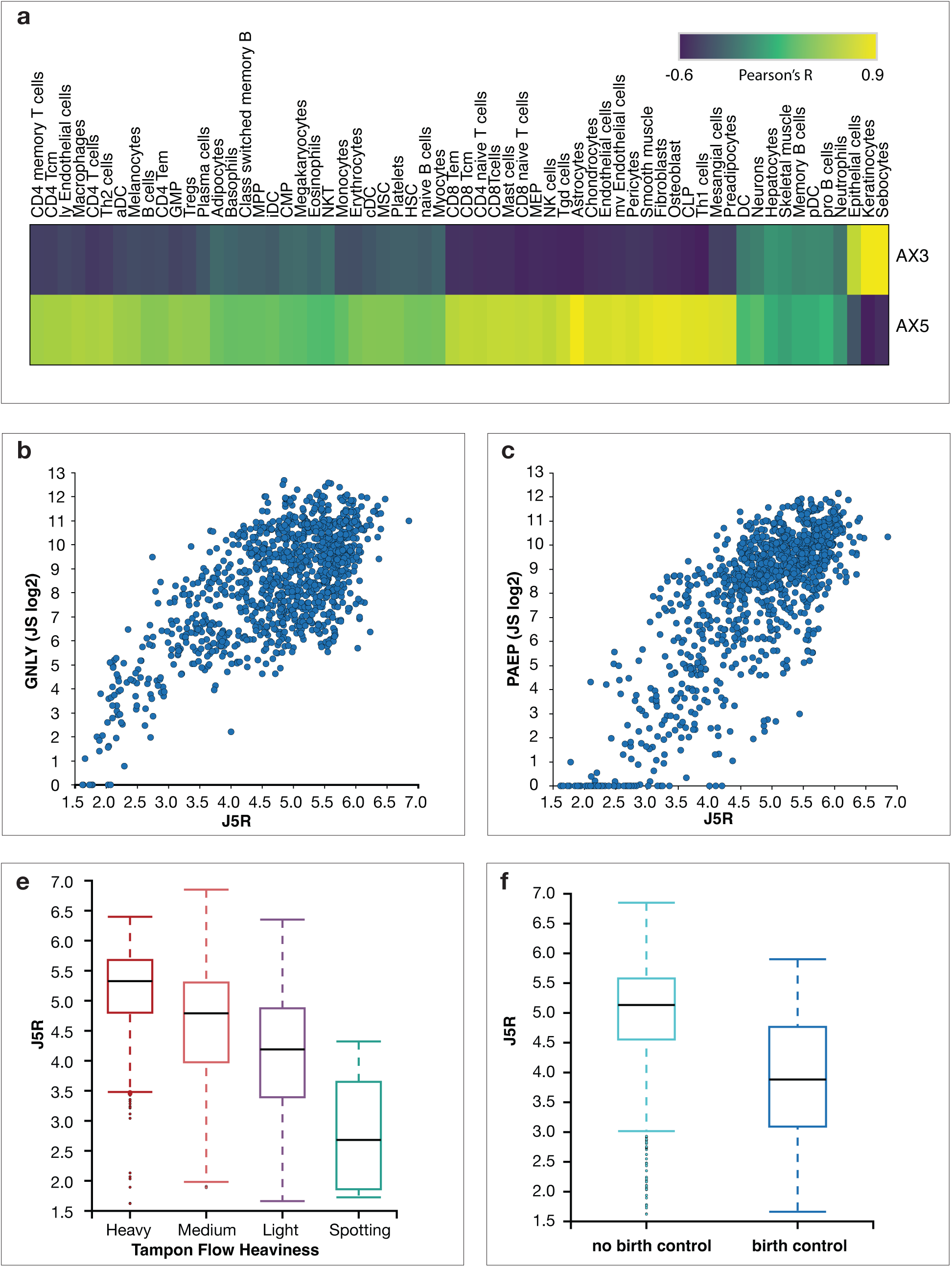
AX5 and AX3 biological structure. Analysis of 1,135 sequence libraries (601 tampons). **(a)** Pearson correlation of 64 cell type activity derived from gene sets included in UCSF’s cell type enrichment analysis pipeline against AX5 and AX3 gene modules. AX3 is highly enriched for epithelial derived cell types. AX5 is enriched broadly for stromal, myeloid, and lymphoid cell types. **(b) & (c)** Scatter plots of J5R to canonical uterine enriched genes GNLY and PAEP show strong positive correlation. **(d)** J5R boxplot by patient reported flow description. **(f)** J5R plotted by patient not on birth control and on any form of birth control.

**Supplemental Figure 5.**
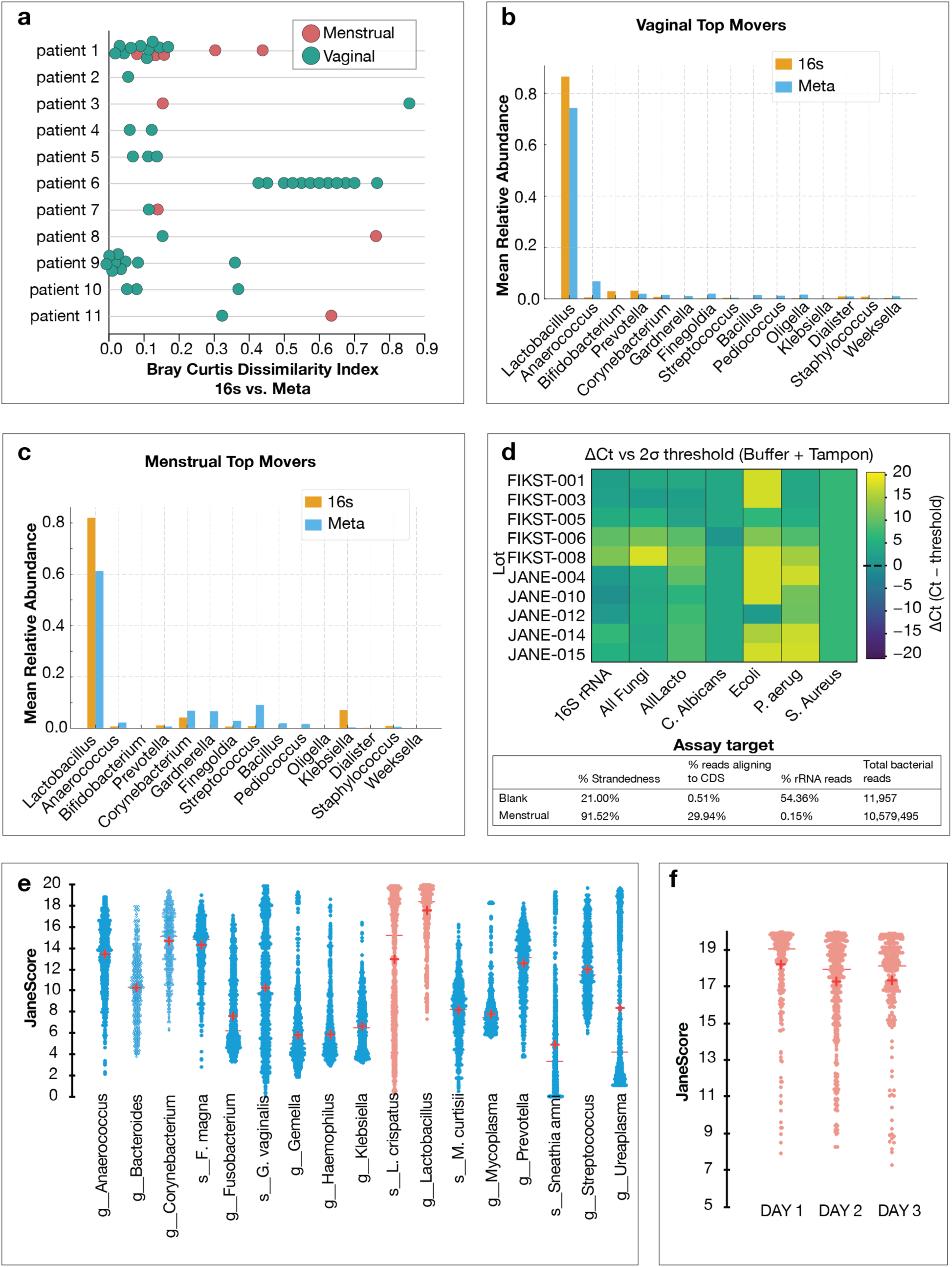
Bacterial analysis of tampon sampling and controlling for contamination. Analysis of 53 tampon samples from 11 patients including vaginal and menstrual tampon collections. **(a)** Bray Curtis dissimilary analysis between 16s DNA relative abundance and metatranstromic relative abundance for vaginal and menstrual samples at the patient level. Stronger dissimilarity exists between patient than within patient. Menstrual samples display larger dissimilarity scores than vaginal, but is highly patient specific. **(b) & (c)** Plots of the mean relative abundance of the top 15 movers between vaginal and menstrual samples and the relative abundance of these bacteria for DNA and RNA. **(d).** Top panel – analysis of qPCR lot testing of blank tampons in buffer compared to established 2 sigma threshold and high and low concentration, pathogen specific controls. These thresholds were established on 100 tested blank samples. For lot testing, a representative sampling of devices was chosen at random for testing. Positive values represent lot testing showed lower overall detection of pathogens compared to the 2 sigma threshold. No lots showed a negative delta Ct for any tampons tested. Lower panel shows the average QC metrics for menstrual tampons and blank tampons sequenced, to establish the overall contamination or bioburden of blank tampons. Blank tampons show low biomass with less than 15,000 reads detected. **(e)** Genus and species level activity across dataset for lactobacillus and bacterial vaginosis associated bacteria. **(f)** Genus level lactobacillus activity by cycle day, showing decrease in overall activity between days 1 and 2.

**Supplemental Figure 6.**
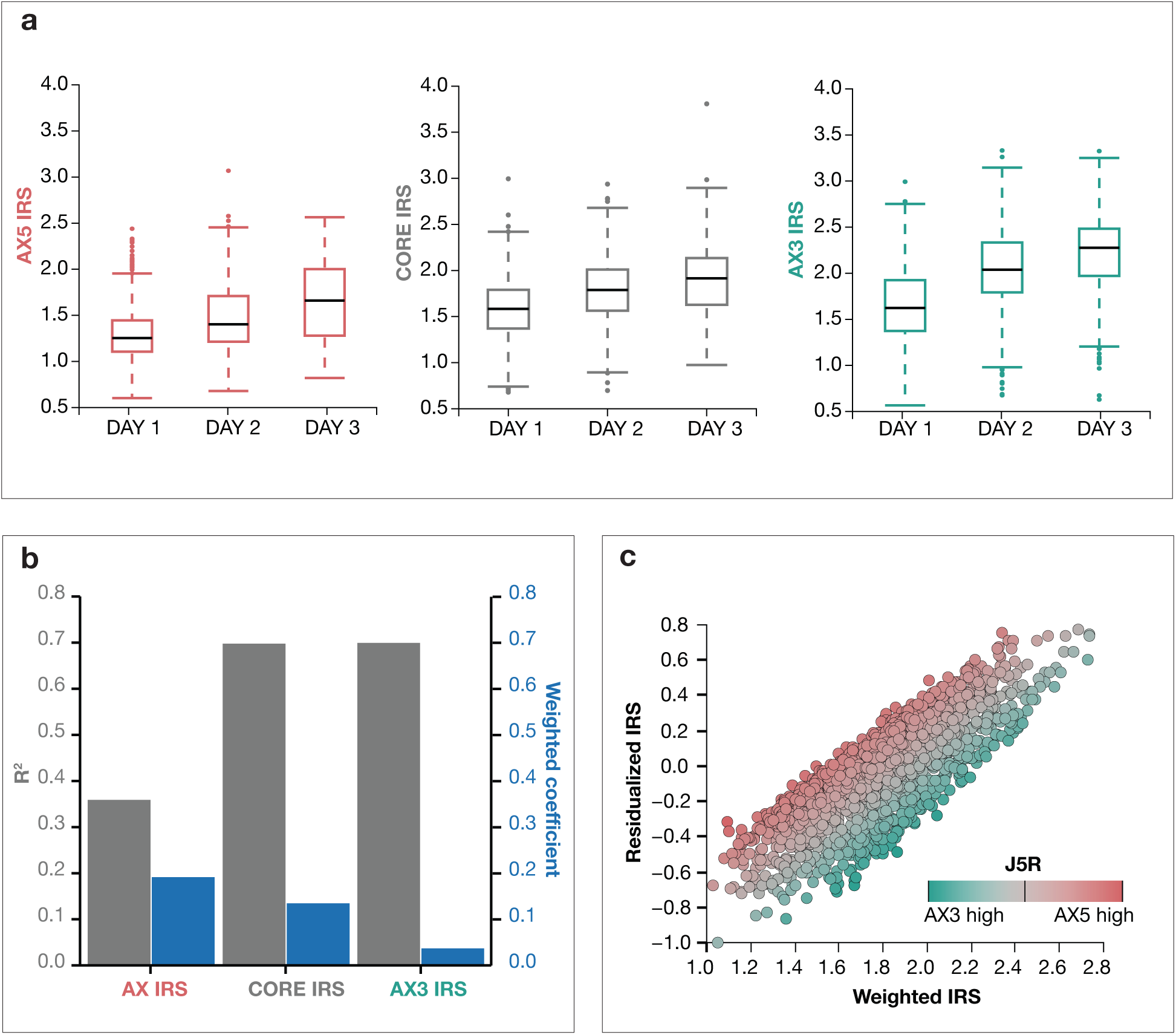
IRS development. IRS development from 1,116 sequences (562 cycle day 1-3 tampons). **(a)** Boxplots of tissue specific IRS models (ridge regression) by cycle day. **(b)** Primary axis: R-squared values of each IRS model to cycle day. Core and AX3 IRS models show highest R-squared values. Secondary axis: Weights given to each IRS model in the combined “mixed IRS”. AX5 has the highest contribution because most samples in our database are highly uterine enriched. **(c)** Scatter plot of “mixed IRS” before residualization on J5R and after. A strong correlation still exists between residualized and non-residualized IRS, but shows less dependence on AX5 and AX3 (main figure 2b).

**Supplemental Figure 7.**
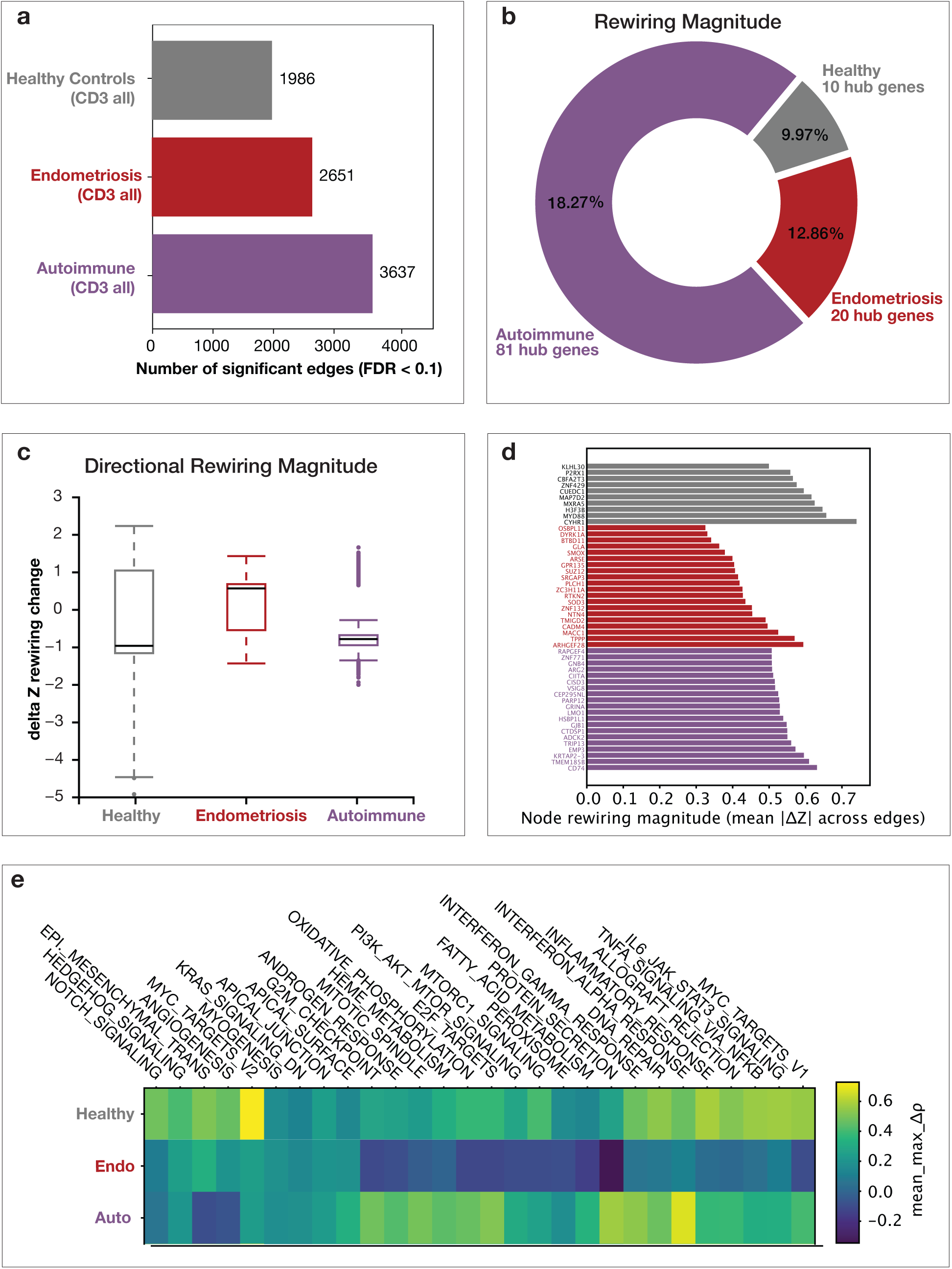
Cycle day 3 HALLMARK network rewiring analysis between resolved and unresolved tampons. Supplemental results from main Figure 3a and b. Analysis of 113 cycle day 3 tampons from 91 individuals: healthy controls (24 tampons), endometriosis (55 tampons), and autoimmune disease (34 tampons). Resolved vs. unresolved (median IRS for each cohort) tampons are highlighted by filled in or empty circles. **(a**) **& (b)** Analysis of top gene nodes and edges between resolved and unresolved by disease cohorts. Healthy – 10 gene hubs at FDR < 0.1 show 1,986 significant edges (gene to gen interactions), representing 9.97% of the interaction network based on 19,900 interactions. Endometriosis – 20 gene hubs show 2,651 significant edges, representing 12.8% of the interaction network. Autoimmune – 81 gene hubs show 3,637 significant edges representing 18.27% of the interaction network. **(c)** ΔZ analysis, defined as the change in correlation of networks from unresolved minus resolved that are then Z-scored to better represent magnitude of interaction change. Positive shifts are more rewired in unresolved. Negative shifts are more rewired in resolved. **(d)** Top 20 significant gene nodes by disease cohort (FDR <0.1) and their absolute magnitude |ΔZ|. **(e)** Δ rho (change in mean correlation of genes in a pathway relative to a hub), quantifies the directional bias of rewiring coordination at the pathway level. Δ rho here is rho unresolved minus rho resolved. Positive values denote stronger overall rewiring in unresolved networks, where negative values represent rewiring in resolved networks.

**Supplemental Figure 8.**
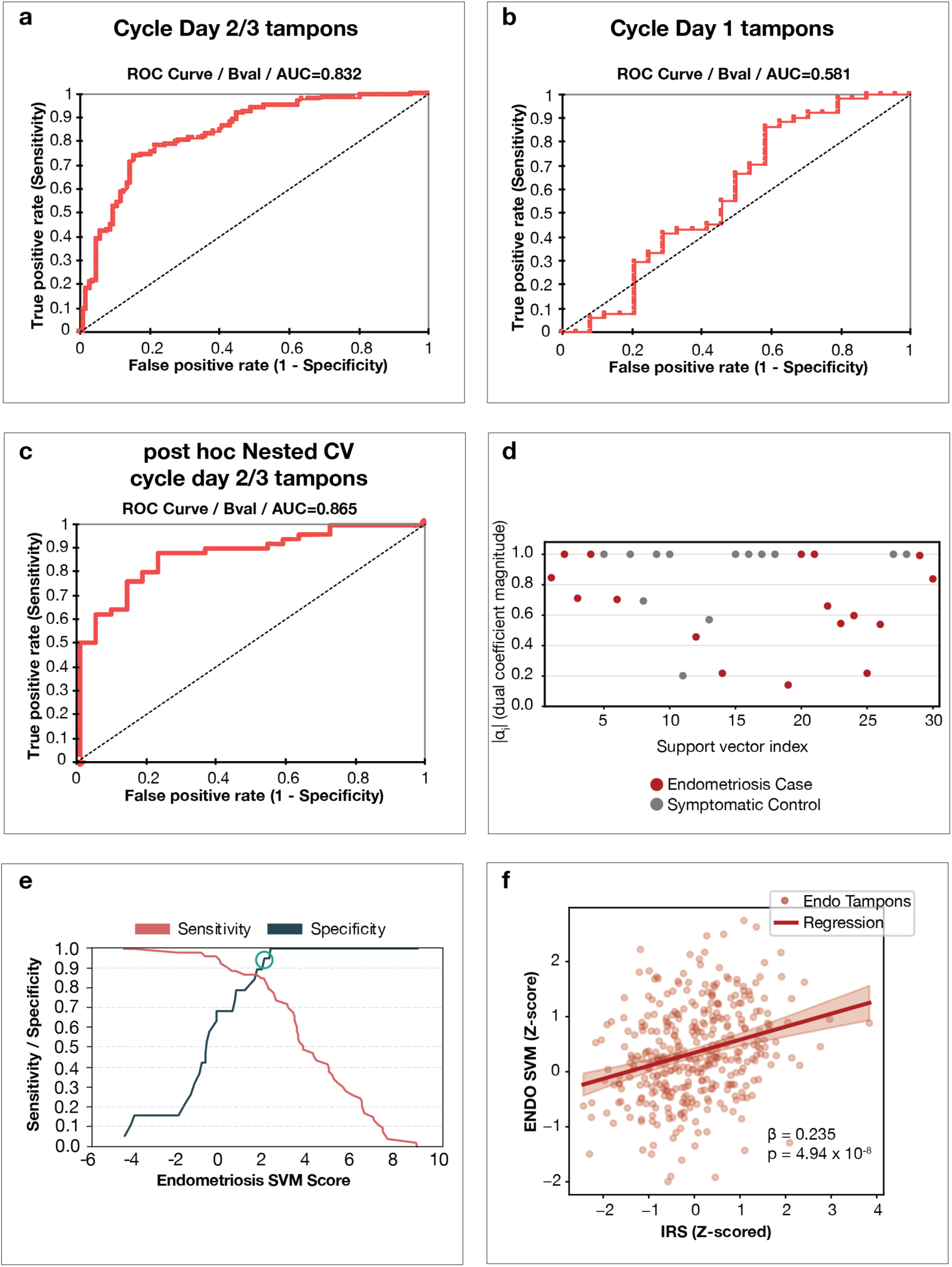
Endometriosis SVM classifier characteristics. **(a)** & **(b)** Analysis 167 patients ( 93 cases and 74 controls) in holdout validation and their performance on the Endo SVM classifier, broken out by performance on cycle days 2 and 3 (139 all comers validation cohort), and cycle day 1. 73 cases and 66 controls had a cycle day 2 or 3 sample for analysis of performance in panel a. Cycle day 1 performance was analyzed from 46 cases and 18 controls where a cycle day 1 tampon was available. **(c)** Post-hoc nested cross validation average AUC results. This analysis was performed on all 185 patients from training and validation (46 training patients and 139 validation patients), to estimate generalizability and potential overfitting of our classifier developed on 47 patients. **(d)** SVM dual coefficient for the 30 support vectors that define the Endo SVM algorithm, colored by endometriosis case (red) or control (gray). **(e)** Sensitivity analysis to determine optimal sensitivity and specificity parameters. Teal circle represents the optimization of specificity (0.947) without significant loss of sensitivity (0.849). Breakdown of patient characteristics can be found in Supplementary Results Tables S5 and S6. **(f)** Scatter plot of IRS and Endo SVM classifier from 747 seq libraries (cycle days 1-3). Within endometriosis cases, ENDO SVM scores are positively associated with IRS Slope ß= 0.235; pvalue = 1×10^−4^, indicating that higher diagnostic scores correspond to increased IRS states. The fitted regression line and confidence band illustrate the stability of this relationship across the observed IRS range.

