## Supplemental Methods for "Quantifying inflammatory resolution in human menstruation reveals disease-specific failure modes and enables a non-invasive diagnostic for endometriosis"

**Supplementary Methods**

**S1. Sample collection, stabilization, RNA extraction, and sequencing quality control**

S1.1 Study population and sample eligibility

Menstrual effluence samples were collected from participants enrolled under IRB-approved protocols (20192619, 20191947, 20233438). Participants provided informed consent for tampon-based specimen collection and associated clinical metadata.

Samples were eligible for inclusion if they met all of the following criteria:

1. Collected on menstrual cycle days 1–5, with downstream analyses further restricting cycle days as specified in later sections.
2. For samples collected outside of menstruation, data was recorded for ovulation or non-ovulation samples
3. Accompanied by complete metadata for cycle day, kit identifier, and sequencing QC metrics
4. Successfully sequenced and passed all RNA sequencing quality control thresholds defined below

Samples collected outside menstruation (e.g., off-cycle cervicovaginal tampons) were excluded from all analyses unless explicitly stated otherwise.

S1.2 At-home tampon collection and ambient stabilization

Participants collected menstrual effluence using a standardized, low-absorbency organic cotton tampon provided as part of an at-home collection kit assembled under an ISO 13485 quality system. Participants were instructed to wear the tampon for four hours during menstruation and 30 minutes outside of menstruation and to record collection start and end times.

Immediately following removal, the tampon was placed into a proprietary collection jar. Upon sealing the jar, a mechanical mechanism released approximately 20 mL of a nucleic-acid stabilization buffer (Norgen Biotek) directly onto the tampon. This buffer lyses cells and preserves host and microbial nucleic acids at ambient temperature. Patient records cycle day and flow characteristics, including any pain medication during current collection on the lid of the box before returning through mail.

Samples were returned to the laboratory by standard mail without cold chain.

S1.3 Lot **release contamination testing and nucleic acid preparation**
Lot release testing was performed on randomly selected NAPS devices from each manufacturing lot in accordance with established quality management procedures. Devices were assessed under two conditions: NAPS buffer alone and NAPS buffer following exposure to a TOTM Light tampon and harvesting step. For each device, aliquots of recovered buffer were subjected to particulate analysis, nucleic acid extraction, and downstream molecular testing. Genomic DNA and RNA were extracted from 150 µL of each sample using the Norgen DNA/RNA Purification Micro Kit, with a non-template control processed in parallel to monitor background contamination. Extracted nucleic acids were quantified using Qubit high-sensitivity assays, and integrity was evaluated by Fragment Analyzer analysis using predefined DNA and RNA quality criteria. Samples exceeding established thresholds for particulate contamination, nucleic acid yield, or fragment profiles were classified as failures based on lot-specific acceptance criteria defined during initial validation studies.
To improve detection sensitivity for low-abundance contaminants, extracted DNA samples and controls were subjected to multiplex PCR preamplification prior to quantitative PCR. Preamplification reactions were assembled using Takara Bio preamplification master mix and a pooled primer/probe set targeting bacterial, fungal, and human sequences, including 16S rRNA, Lactobacillus, Candida albicans, Staphylococcus aureus, Pseudomonas aeruginosa, and Escherichia coli High and low positive controls were generated by serial dilution and included alongside experimental samples and non-template controls. Preamplification was performed for 14 cycles on a Bio-Rad CFX96 thermal cycler using a standardized protocol, after which products were diluted and analyzed by TaqMan-based qPCR on an Applied Biosystems 7900HT system. All samples and controls were run in technical triplicate using a fixed 384-well plate layout to control for assay and positional effects. Average cycle threshold (Ct) values were compared against assay- and condition-specific thresholds defined as two sigma below the mean of validated control distributions, with samples falling below threshold classified as failures for lot release determination

S1.4 Laboratory processing and aliquoting

Upon receipt, collection date and laboratory accession date were recorded for each sample to enable calculation of time in buffer. Weight of collection (before - during manufacturing, and after collection are taken to assess total weight of blood collected). A semi-quantitative color value is assigned to every sample, and an alkaline hematin equivalent value (535nm) is taken upon a 2-fold serial dilution of sample.

Samples were processed in a biosafety cabinet. Tampons were extruded from the collection jar using a custom plastic harvester designed to minimize fiber shedding. The recovered lysate was transferred to a 50 mL conical tube and centrifuged at 3,400 × g for 10 minutes to pellet debris.k Approximately 17mLs of lysate and sample are recovered per tampon collection.

Lysate was then aliquoted into 2 mL and 5 mL cryovials and stored at −80 °C prior to nucleic acid extraction or sequencing.

S1.5 RNA extraction protocols

Total RNA was extracted using one of two validated protocols:

1. Column-based extraction
   Norgen Deep 96-well Total RNA extraction plates were used according to the manufacturer’s protocol, with the following modifications:
   - 1.2 mL of lysate per sample was split across two wells
   - Eluates were combined
   - In-solution DNase treatment was performed using Turbo DNase (37 °C, 30 min)
   - RNA was cleaned and concentrated using Beckman RNA XP beads
2. Magnetic bead-based extraction
   ThermoFisher MagMAX mirVana Total RNA Isolation was performed on 600 µL of lysate with protocol modifications optimized for menstrual effluence, including a reduced isopropanol ratio to balance recovery of large and small RNA fragments.

Extraction method choice did not differ systematically by cohort or outcome and was validated for yield and reproducibility as described in Supplementary Results I.

RNA quantity was measured using Qubit fluorometry. RNA quality was measured on an Agilent Fragment Analyzer. Extracted RNA was used immediately for library preparation.

S1.6 RNA-seq library preparation and sequencing

RNA libraries were prepared using the Zymo-Seq RiboFree Total RNA Library Kit with 500 ng of total RNA input per sample. Ribosomal RNA was depleted enzymatically. Libraries were amplified for 12 PCR cycles, purified using AMPure XP beads, and assessed for fragment size distribution using an Agilent Fragment Analyzer.

Sequencing was performed on Illumina NextSeq platforms using paired-end 100 bp reads (200-cycle P3 and P4 kits). Samples were pooled to target approximately 60 million reads per library.

S1.7 RNA-seq preprocessing

Raw FASTQ files were processed as follows:

1. Quality assessment using FastQC
2. Adapter trimming using CutAdapt
3. Alignment to the human reference genome (hg38) using STAR
4. Gene-level read counting using FeatureCounts

Reads not aligning to the human genome were retained for downstream microbial analyses described in S9.

S1.8 Correction for transcript degradation

To account for RNA degradation inherent to menstrual effluence, gene-level counts were normalized using DegNorm. DegNorm estimates transcript-specific degradation by modeling 5′–3′ coverage bias and corrects expression values relative to high-quality reference samples.

Three pooled, high-quality menstrual RNA samples were used as DegNorm references. All downstream expression analyses were conducted on DegNorm-corrected counts.

RNA integrity was quantified using the DegNorm-derived DI₍25₎ metric, representing the 25th percentile of transcript degradation across genes.

S1.9 Sequencing quality control thresholds

Samples were excluded from all downstream analyses if any of the following criteria were not met:

- Strandedness < 80%
- Ribosomal RNA content > 3%
- Uniquely mapped reads < 8 million
- Excess Y-chromosome transcript signal, indicating likely semen contamination

QC failures were identified based on coordinated deviation across multiple metrics rather than marginal threshold effects. Samples that initially failed QC but passed after resequencing were retained; samples that failed irrecoverably were excluded.

All QC filtering was applied prior to normalization, tissue modeling, IRS construction, and any statistical analysis.

S1.10 Units of analysis and aggregation rules

Unless otherwise specified:

- Sample-level analyses were used for:
  - RNA integrity (DI₍25₎)
  - Normalization benchmarking
  - Microbial abundance and activity comparisons
- Kit-level analyses were used for:
  - Inflammatory Resolution Score (IRS)
  - Endothelial coupling analyses
  - Network rewiring analyses
  - Diagnostic classifier development

For kit-level analyses, multiple samples associated with the same kit_id were aggregated by taking the mean of gene expression values, switch scores, and derived metrics. This aggregation was used to avoid pseudoreplication while preserving biological signal. Unless explicitly stated otherwise, all downstream biological and modeling analyses were performed at the kit level. Where multiple RNA-seq libraries were generated from the same kit, expression values, switch scores, and derived metrics were aggregated by taking the mean across libraries associated with that kit prior to analysis. Sample-level analyses were restricted to sequencing quality control, RNA integrity assessment, normalization benchmarking, and microbial activity comparisons.

**S2. RNA-seq normalization, expression matrices, and benchmarking of normalization strategies**

S2.1 Purpose and scope

This section defines the normalized gene expression values used in all downstream analyses and specifies the normalization strategy selected for this study. Because menstrual effluence is a compositionally heterogeneous biospecimen with large, biologically meaningful variation in tissue admixture, normalization choices materially affect interpretability. All analyses reported in the main manuscript and Supplementary Results were therefore conducted in a reference-anchored expression space (JaneScore), defined explicitly below.

S2.2 Input expression data

Normalization was applied to RNA-seq gene-level counts that satisfied all preprocessing and QC criteria described in Supplementary Methods S1.

Input format:

- Rows: gene symbols (hg38 annotation)
- Columns: sample_id
- Values: DegNorm-corrected RPKM counts

Genes with zero variance across all retained samples were excluded prior to normalization. No additional filtering by mean expression or prevalence was applied at this stage.

S2.3 RPKM calculation

For each gene *g* in sample *s*, reads per kilobase per million mapped reads (RPKM) were computed using DegNorm-corrected counts:

$$\text{RPKM}_{g,s}=\frac{\text{counts}_{g,s}}{\text{gene length}_{g}/1000}\div\frac{\text{total mapped reads}_{s}}{{10}^{6}}$$

Gene lengths were derived from the hg38 reference annotation used by FeatureCounts. Only uniquely mapped reads were included in the library size denominator.

S2.4 JaneScore normalization (exact definition)

All downstream expression analyses were conducted using JaneScore, a reference-anchored normalization designed to stabilize expression values across heterogeneous tissue admixture while preserving secondary biological structure.

For each gene *g* in sample *s*, JaneScore was defined as:

$$\text{JaneScore}_{g,s}={log}_{2}(\text{RPKM}_{GENEXg,s})-[{log}_{2}(\text{geomean (RPKM}_{\text{ACTB},s},\text{RPKM}_{\text{GAPDH},s},\text{RPKM}_{\text{RAB7A},s})]+10$$

where ACTB, GAPDH, and RAB7A serve as internal reference genes selected for:

- high expression across samples,
- low variance across cervicovaginal- and uterine-enriched specimens,
- minimal correlation with tissue composition axes.

The additive constant (+10) ensures all JaneScore values are positive and preserves log₂ interpretability. No further centering, scaling, or variance stabilization was applied. The resulting JaneScore matrix (genes × samples) constitutes the canonical expression matrix used in all subsequent analyses unless otherwise specified.

S2.5 Construction of switch-level expression matrices

Cell-specific switch scores (defined in S4) were computed using median JaneScore-normalized gene expression values. For the gene set. Switch-level matrices were therefore intrinsically normalized and did not undergo additional transformation. Switch scores were stored as log₂-scaled values and aligned to the same sample_id index as the gene-level JaneScore matrix.

S2.6 Alternative normalization strategies evaluated

For benchmarking purposes only, three additional normalization strategies were evaluated on a restricted subset of samples spanning cervicovaginal-dominant, uterine-dominant, and mixed tissue profiles:

1. Raw counts, log₂(1 + counts)
2. RPKM, log₂(RPKM)
3. DESeq2 size-factor normalization, log₂(1 + normalized counts)

These alternative normalizations were used exclusively for comparative evaluation and were not used for primary analyses, model training, or reported results. DESeq2-normalized values were used exclusively for normalization benchmarking and were not used for feature selection, modeling, or any reported results.

S2.7 Benchmarking criteria for normalization choice

Normalization strategies were evaluated based on their ability to:

1. Preserve the dominant tissue-composition gradient
2. Preserve secondary, biologically interpretable structure beyond the dominant axis
3. Minimize coupling of housekeeping genes to tissue composition
4. Reduce subject-specific and technical imprinting in secondary principal components

Principal component analysis (PCA) was performed on each normalized matrix after gene-wise centering.

For each normalization method, PC1 and PC2 were modeled as linear functions of AX3 and AX5 (defined in S3). Model fit was quantified using R², F-statistics, and associated *p*-values.

S2.8 Preservation of biological structure under JaneScore

Under all normalization methods, PC1 was dominated by tissue composition, with AX3 and AX5 jointly explaining the majority of PC1 variance. However, divergence emerged at PC2.

Under JaneScore normalization:

- AX3 and AX5 jointly explained a substantial fraction of PC2 variance
- PC2 correlated more strongly with biological covariates than with technical metrics
- Housekeeping genes showed minimal correlation with tissue axes

Under DESeq2 normalization:

- PC2 exhibited weak alignment with AX3 and AX5
- PC2 was dominated by subject identity and technical sequencing metrics
- Housekeeping genes remained strongly correlated with tissue composition

These results are reported quantitatively in Supplementary Results and motivated selection of JaneScore as the primary expression space.

S2.9 Final expression matrices used downstream

All downstream analyses (S3–S10) used the following matrices:

1. Gene-level JaneScore matrix
   - Rows: genes
   - Columns: sample_id
   - Values: JaneScore (log₂ scale)
2. Switch-level activity matrix
   - Rows: switches
   - Columns: sample_id
   - Values: log₂ switch activity scores derived from JaneScore

Principle component analysis PC1 and PC2 were calculated on initial two sequencing runs to capture technical and biological variation that could be used in subsequent runs to monitor drift and batch effects. Batches that fell outside of this pre-established range were excluded from analysis. This was to ensure that batch correction was not utilized an any analysis, but instead used as a QC filter that would initiate re-sequencing. This process insures high fidelity data output needed for clinical applications.

No additional normalization or scaling was applied after this stage unless explicitly stated in the relevant section.

**S3. Tissue composition modeling using AX3, AX5, and the uterine enrichment metric J5R**

S3.1 Purpose and scope

Menstrual effluence is a compositionally heterogeneous biospecimen containing variable contributions from uterine-derived tissue, cervicovaginal epithelium, immune cells, blood, and microbial material. To explicitly model this heterogeneity rather than filtering it away, we derived two transcriptional axes—AX3 and AX5—that capture the dominant cervicovaginal-to-uterine continuum in menstrual samples. These axes were subsequently combined into a continuous uterine enrichment metric (J5R) used for stratification, residualization, and sensitivity analyses throughout the manuscript. This section specifies the derivation, scoring, and downstream use of AX3, AX5, and J5R.

S3.2 Discovery dataset for axis derivation

AX3 and AX5 were derived using an internal reference dataset comprising menstrual effluence samples, cervicovaginal tampon samples, and matched venous blood samples collected using the same at-home platform and processed through the a different RNA-seq pipeline. These earlier samples were processed using RNA Trizol extractions and prepped using Illumina’s exome plus, globin minus RNA preparation protocol and reagents, a hybrid-probe based enrichment procedure. All samples were sequenced on an Illumina HiSeq. Because of this, these samples were not folded into future samples and instead used as an independent development cohort. This dataset was used exclusively for axis discovery and validation, and for GTex comparisons. Once defined, AX3 and AX5 gene sets were treated as fixed and were not modified, re-trained, or re-optimized in any downstream analyses.

S3.3 Derivation of AX3 and AX5 tissue composition axes

AX3 and AX5 were derived through a single, integrated pipeline designed to identify stable, high-confidence co-expression programs that distinguish cervicovaginal-dominant from uterine-dominant transcriptional states.

First, candidate tissue-associated genes were identified by differential expression analysis between cervicovaginal tampon samples and menstrual effluence samples using RPKM-normalized expression values. Differential expression was assessed using two-sided Welch *t*-tests with Benjamini–Hochberg false discovery rate correction. Genes were retained as candidates if they met all of the following criteria:

- Statistically significant differential expression after FDR correction
- Consistent directionality across replicate samples

These candidate genes were then used as input to co-expression network analysis rather than treated as final tissue markers.

Next, a gene–gene correlation network was constructed by computing pairwise Spearman correlation coefficients across all candidate genes. To identify robust transcriptional programs rather than diffuse correlated sets, we applied an iterative clique-trimming procedure. In this procedure, maximal cliques of size 2–8 were identified, and genes were retained if they participated in multiple higher-order cliques, reflecting dense and reproducible co-expression structure. Genes with low connectivity or isolated correlations were pruned.

This combined differential-expression–seeded, clique-trimmed co-expression analysis yielded multiple candidate modules. Two modules were selected based on their biological interpretability, stability across samples, and consistent separation of tissue types:

- AX3, a module enriched in cervicovaginal samples and dominated by epithelial programs
- AX5, a module enriched in menstrual effluence samples and capturing uterine-associated biology spanning stromal, immune, and vascular programs

Once selected, the gene membership of AX3 and AX5 was fixed. No genes were added, removed, or re-weighted in downstream analyses.

S3.4 Scoring of AX3 and AX5 in individual samples

For each sample *s*, AX3 and AX5 scores were computed using JaneScore-normalized expression values as:

$$AX3_{s}=\text{median}(\text{JaneScore}_{g,s}\text{ }\mid\text{ }g\in AX3)$$

$$AX5_{s}=\text{median}(\text{JaneScore}_{g,s}\text{ }\mid\text{ }g\in AX5)$$

Median aggregation was chosen to reduce sensitivity to outlier genes and residual technical variation while preserving the central tendency of each transcriptional program. Scores were computed at the sample level and subsequently aggregated to the kit level by taking the mean across all samples associated with a given kit for analyses requiring kit-level resolution.

S3.5 Validation of tissue specificity and stability

AX3 and AX5 were validated by examining their behavior across known tissue types and biological correlates. Specifically, we:

1. Compared AX3 and AX5 score distributions across cervicovaginal samples, menstrual effluence samples, and venous blood samples
2. Assessed correlations with canonical tissue-associated genes, with AX3 aligning with epithelial markers and AX5 aligning with uterine, immune, and vascular markers
3. Evaluated tissue classification accuracy using a k-nearest neighbors classifier in the two-dimensional AX3–AX5 space

These analyses demonstrated that AX3 and AX5 jointly define a continuous transcriptional gradient rather than discrete tissue classes, supporting their use as quantitative composition axes.

S3.6 Definition of the uterine enrichment metric J5R

To summarize tissue composition as a single continuous value, AX3 and AX5 scores were combined into the uterine enrichment metric J5R:

$$J5R=10\times\frac{AX5}{AX3+AX5}$$

The scaling factor of 10 preserves interpretability on a log₂-like scale while maintaining numerical stability. J5R increases monotonically with uterine contribution and decreases with increasing cervicovaginal contribution.

J5R was used as a continuous variable for stratification, residualization, and sensitivity analyses rather than as a hard inclusion filter.

S3.7 Downstream use and fixed definitions

AX3, AX5, and J5R were used downstream for:

- Stratification of uterine-enriched and cervicovaginal-enriched samples
- Residualization of tissue composition effects during IRS construction
- Sensitivity analyses assessing robustness to tissue admixture
- Explicit covariates in diagnostic modeling

To ensure reproducibility:

- AX3, AX5, and J5R are deterministic functions of JaneScore-normalized expression
- No re-training or re-derivation of tissue axes was performed in downstream analyses

**S4. Cell-specific switch construction and annotation**

S4.1 Purpose and scope

To quantify coordinated changes in cellular activity within menstrual effluence, we summarized gene-level expression into a fixed set of cell-specific transcriptional programs (“switches”). These switches represent biologically interpretable cellular states spanning immune, stromal, epithelial, endothelial, and hematopoietic compartments and serve as the fundamental units for all coordination, coupling, and network analyses reported in the manuscript. This section defines the source, construction, scaling, annotation, and downstream use of these cell-specific switches.

S4.2 Source of cell-specific gene sets

Cell-specific gene sets were obtained from xCell, a curated compendium of transcriptional signatures designed to digitally portray tissue cellular heterogeneity across diverse immune and stromal cell types. xCell gene sets were used as provided by the authors and were not modified, re-weighted, or re-trained within this study.

Each xCell gene set corresponds to a defined CellType (e.g., macrophage, dendritic cell, endothelial cell, fibroblast) and is associated with a higher-level Subgroup annotation (e.g., Myeloid, Lymphoid, Endothelial, Stroma, Epithelial, Hematopoietic Stem Cell).

S4.3 Input expression data

Switch construction was performed using the JaneScore-normalized gene expression matrix defined in Supplementary Methods S2.

Input requirements:

- Rows: gene symbols
- Columns: sample_id
- Values: JaneScore (log₂ scale)

Only genes present in both the JaneScore matrix and the xCell gene sets were used for switch construction. No imputation was performed for missing genes.

S4.4 Switch activity score calculation

For each sample *s* and each xCell gene set *k*, a switch activity score was computed as the median JaneScore across all genes belonging to the xCell gene set:

$$\text{SwitchScore}_{k,s}=\text{median}(\text{JaneScore}_{g,s}\text{ }\mid\text{ }g\in\text{xCell}_{k})$$

Median aggregation was chosen to. No gene-level weighting was applied within gene sets.

- reduce sensitivity to outlier genes,
- minimize amplification of residual technical variation,
- preserve relative ordering of cellular activity across samples.

S4.5 Scaling and transformation

Switch scores were retained on the log₂ JaneScore scale and were not z-scored, centered, or otherwise transformed at this stage. All downstream analyses explicitly state when additional scaling (e.g., z-scoring across kits) was applied for a specific purpose. A total of 489 switches were constructed and retained for downstream analysis. The resulting switch matrix had the following structure:

- Rows: switch identifiers (xCell gene sets)
- Columns: sample_id
- Values: log₂ switch activity scores

S4.6 Aggregation to kit-level resolution

For analyses conducted at the kit level (IRS construction, endothelial coupling, network rewiring, diagnostic modeling), switch scores were averaged across all samples that passed QC and associated with that kit:

Mean aggregation was used at the kit level to preserve quantitative differences in activity magnitude while avoiding pseudoreplication from multiple samples per kit. Sample-level switch scores were retained unchanged for analyses explicitly performed at the sample level.

S4.7 Switch annotation and ontology

Each switch was annotated with:

- CellType (as defined by xCell)
- Subgroup, one of:
  - Epithelial
  - Stroma
  - Myeloid
  - Lymphoid
  - Endothelial
  - Hematopoietic Stem Cell

These annotations were used for:

- subgroup-level decomposition of global rewiring,
- organization and labeling of heatmaps,
- interpretation of endothelial coupling analyses,
- stratification of cell-type pair contributions.

The switch ontology table was treated as fixed and is provided in the Supplementary Data.

S4.8 Downstream use of switch scores

Switch scores served as inputs for the following analyses:

- Cycle-day coordination analysis (S5), where switch–switch correlation matrices were computed
- Inflammatory Resolution Score (IRS) construction (S6), where switches contributed as candidate features
- Endothelial coupling analysis (S7), where switches were correlated with endothelial activity scores

At no point were switch gene sets redefined, filtered, or re-weighted based on disease status, cycle day, or outcome.

S4.9 What is fixed versus learned

For clarity and reproducibility:

- xCell gene sets are external and fixed
- Switch construction is a deterministic transformation of JaneScore expression
- The full set of 489 switches is retained across all analyses
- No feature selection is applied at the switch-construction stage

All selection, weighting, or modeling decisions involving switches are described explicitly in later sections.

**S5. Cycle-day coordination and global rewiring of cell-specific programs**

S5.1 Purpose and scope

This section defines how coordinated changes in cell-specific transcriptional programs were quantified across the early menstrual window in healthy samples. The goal of this analysis was to establish a normative, system-level baseline for menstrual inflammatory resolution by measuring how correlations between cell-specific switches evolve from cycle day 1 through cycle day 3.

All analyses in this section were performed exclusively in healthy reference samples and define the biological framework that motivates subsequent construction of the Inflammatory Resolution Score (IRS).

S5.2 Input data and unit of analysis

Inputs to this analysis were:

- The cell-specific switch matrix defined in Supplementary Methods S4
- Associated metadata including sample_id, kit_id, patient_id, and cycle_day

Analyses in this section were conducted at the sample level rather than the kit level to preserve within-day biological variability. No aggregation across kits or patients was performed. Only samples meeting all QC criteria defined in Supplementary Methods S1 were included.

S5.3 Sample selection and stratification by cycle day

Analyses were restricted to samples collected on menstrual cycle days 1, 2, or 3, from patients who were not on birth control

For each cycle day independently:

- Exactly ten tampon samples were selected from healthy individuals
- Samples were selected from healthy samples meeting QC and cycle-day criteria, with no additional outcome-based filtering

These three day-specific sample sets (CD1, CD2, CD3) were treated as independent reference cohorts.

S5.4 Construction of cycle-day–specific coordination matrices

For each cycle day *d* ∈ {1, 2, 3}, a switch–switch correlation matrix was constructed as follows:

1. For the 489 cell-specific switches, pairwise Pearson correlation coefficients were computed across the ten samples collected on day *d*
2. This produced a symmetric 489 × 489 correlation matrix *R<sub>d</sub>*
3. Diagonal entries (self-correlations) were excluded from all downstream analyses

Correlation coefficients were computed using raw switch activity scores without additional scaling or transformation.

S5.5 Quantification of coordination changes across cycle days (Δr)

To quantify changes in coordination between cycle days, edge-wise differences in correlation strength were computed. For two cycle days *d₁* and *d₂*, the change in correlation for each unordered switch pair *(i, j)* was defined as:

$$\Delta r_{ij}^{(d_{2}-d_{1})}=r_{ij}^{(d_{2})}-r_{ij}^{(d_{1})}$$

This procedure yielded a distribution of Δr values across all unique switch–switch edges:

$$\frac{489\times488}{2}=119,316\text{ edges}$$

Δr distributions were computed for:

- CD2 − CD1
- CD3 − CD2
- CD3 − CD1

S5.6 Summary statistics for global coordination shifts

For each Δr distribution, global coordination changes were summarized using:

- Median Δr
- Interquartile range (IQR)
- Fraction of edges with Δr > 0

These statistics quantify the magnitude, spread, and directionality of system-level coordination changes across the menstrual window.

S5.7 Statistical testing of global shifts

Two complementary non-parametric tests were applied to each Δr distribution:

1. Wilcoxon signed-rank test
   - Tests whether the median Δr differs from zero
2. Sign test
   - Tests whether the fraction of edges with Δr > 0 differs from 0.5

Both tests were applied to the full set of edge-wise Δr values. Because of the large number of edges, p-values frequently fell below numerical precision and are reported as *p* < 1 × 10⁻³⁰⁰. No multiple-testing correction was applied at this stage because each test was applied to a single global distribution rather than to individual edges.

S5.8 Decomposition of global rewiring by cell subgroup

To identify which biological compartments contributed most strongly to global coordination changes, total rewiring energy was quantified as the sum of squared correlation changes:

$$\text{RewiringEnergy}=\sum_{i<j} (\Delta r_{ij})^{2}$$

Each edge’s Δr² contribution was divided equally between its two endpoint switches. Contributions were then aggregated by Subgroup annotation (defined in S4), yielding the fraction of total rewiring energy attributable to:

- Epithelial
- Stroma
- Myeloid
- Lymphoid
- Endothelial
- Hematopoietic Stem Cell

Subgroup contributions were normalized to sum to one and interpreted as relative contributions to global network reorganization.

S5.9 Identification of dominant cell-type pair contributions

In parallel, Δr² contributions were aggregated across unordered CellType pairs to identify specific intercellular relationships driving global coordination changes. For each unordered pair of CellTypes *(A, B)*, total rewiring energy was computed as the sum of Δr² across all switch pairs where one switch belonged to CellType *A* and the other to CellType *B*. Pairs were ranked by their cumulative contribution.

No statistical testing was applied at the cell-type pair level; these results are reported descriptively to highlight dominant biological interactions.

**S6. Construction of the Inflammatory Resolution Score (IRS)**

**S6.1 Purpose and design principles**

The Inflammatory Resolution Score (IRS) was designed to quantify a sample’s position along the coordinated menstrual repair trajectory defined in Supplementary Methods S5, while remaining independent of tissue composition variability inherent to menstrual effluent. Unlike static inflammatory burden metrics, IRS was constructed to capture **system-level coordination and timing** of inflammatory resolution rather than absolute expression magnitude.

Key design constraints were therefore:

1. Sensitivity to monotonic coordination changes across cycle days 1–3
2. Robustness to cervicovaginal versus uterine tissue admixture
3. Stability across heterogeneous real-world samples
4. Interpretability as a continuous temporal axis rather than a binary state

**S6.2 Input data and unit of analysis**

IRS construction used the following inputs:

- **JaneScore-normalized gene expression matrix** (Supplementary Methods S2)
- **Cell-specific switch matrix** (Supplementary Methods S4)
- **Tissue composition metrics** AX3, AX5, and J5R (Supplementary Methods S3)
- **Cycle day annotations** (days 1–3)

IRS was computed at the **sample level**. For downstream analyses of IRS, all samples associated with a kit IRS averaged across replicates. Only samples meeting all QC criteria defined in Supplementary Methods S1 were eligible.

**S6.3 Candidate feature pool**

Candidate IRS features consisted of a combined pool of:

- **Gene-level features** (JaneScore expression)
- **Switch-level features** (cell-specific switch scores)

Features were eligible for consideration if they met all of the following criteria:

1. Mean JaneScore (or mean switch score) ≥ 2 across eligible samples
2. Demonstrated monotonic or near-monotonic dependence on cycle day in healthy samples
3. Did not trivially encode tissue composition (see correlation constraints below)

No disease labels were used at any stage of IRS feature selection.

**S6.4 Tissue-aware feature partitioning**

Because the visibility of resolution biology differs by tissue context, features were explicitly partitioned into three biologically motivated regimes prior to modeling:

1. **Uterine-enriched features (AX5-associated)**
   - Features positively correlated with AX5
   - Represent stromal, immune, and vascular repair programs most visible in uterine-rich samples
2. **Cervicovaginal-enriched features (AX3-associated)**
   - Features positively correlated with AX3
   - Capture epithelial-dominant programs present in cervicovaginal-heavy samples
3. **Core (tissue-agnostic) features**
   - Features showing minimal correlation with AX3, AX5, or J5R
   - Represent conserved resolution biology visible across tissue admixture states

Correlation thresholds used for assignment were fixed and applied uniformly across features. Features not meeting criteria for any group were excluded from IRS construction.

This partitioning step prevents a single tissue regime from dominating the score and enables resolution biology to be learned where it is most visible.

**S6.5 Training subsets for component models**

Three independent ridge regression models were trained, each restricted to the tissue regime in which its features were most interpretable:

- **AX5 IRS model**
  - Training data: kits with J5R > 4.5
  - Represents uterine-enriched resolution biology
  - Samples used: 523
- **AX3 IRS model**
  - Training data: kits with J5R < 3.8
  - Represents cervicovaginal-enriched resolution biology
  - Samples used: 184
- **Core IRS model**
  - Training data: kits spanning complete cycle day 1–3 trajectories
  - Represents tissue-agnostic resolution biology
  - Samples used: 331

These thresholds were fixed a priori and used only for model training, not for downstream filtering.

**S6.6 Ridge regression model specification**

Each component IRS model was fit using ridge regression with cycle-day progression as the response variable.

For each component model:

- Input features were standardized (mean 0, variance 1) within the training subset
- Ridge regression was fit with a fixed regularization parameter (α), selected via internal cross-validation and held constant thereafter
- Model fitting was performed using scikit-learn

The fitted models output a continuous predicted value representing progression along the menstrual resolution trajectory within the relevant tissue regime.

**S6.7 Scoring of component IRS values**

Each fitted component model (AX5 IRS, AX3 IRS, Core IRS) was applied to **all eligible kits**, regardless of tissue composition, producing three component scores per kit. Component scores were then standardized across all kits to place them on a comparable scale prior to integration. No clipping or thresholding was applied.

**S6.8 Integration into a mixed IRS**

To integrate information across tissue regimes, standardized component scores were combined using a second ridge regression model:

$$\text{IRS}_{\text{mixed}}=\beta_{0}+\beta_{1}(\text{AX5 IRS})+\beta_{2}(\text{Core IRS})+\beta_{3}(\text{AX3 IRS})$$

This integration model was trained to approximate the original inflammatory index previously developed in this system, using all eligible kits and patient-level cross-validation to prevent leakage. The resulting mixed IRS captures resolution biology visible across tissue contexts while down-weighting regime-specific noise.

**S6.9 Residualization on tissue composition**

Because uterine enrichment strongly influences transcriptional signal magnitude, the mixed IRS retained residual dependence on tissue composition. To remove this effect, mixed IRS values were residualized with respect to J5R (as defined in S3.6):

1. A linear regression was fit:

$$\text{IRS}_{\text{mixed}}=\gamma_{0}+\gamma_{1}\cdot J5R+\epsilon$$

2. Residuals (ε) were extracted
3. Residuals were z-scored across all kits

The resulting value constitutes the **canonical Inflammatory Resolution Score (IRS)** used throughout the manuscript. After residualization, IRS exhibited minimal correlation with J5R while retaining strong cycle-day structure.

**S6.10 Interpretation of IRS values**

IRS is a **relative, continuous measure** of resolution timing:

- Lower IRS values indicate delayed or unresolved inflammatory states
- Higher IRS values indicate more advanced progression along the repair trajectory

IRS does not represent inflammatory burden, tissue abundance, or disease probability, and was never thresholded during construction. Thresholds used to define “resolved” versus “unresolved” states in downstream analyses were applied *post hoc* and are described explicitly in the relevant sections.

**S6.11 Validation and robustness checks**

IRS behavior was evaluated for:

- Monotonic increase across cycle days in healthy samples
- Stability across repeated kits from the same individual
- Minimal dependence on tissue composition after residualization
- Sensitivity to hormonal suppression

These checks are reported in Supplementary Results and were not used to tune the score.

**S6.12 What is fixed versus learned**

For reproducibility:

- Feature eligibility rules are fixed
- Tissue partition thresholds (J5R > 4.5, J5R < 3.8) are fixed
- Model regularization parameters are fixed after selection
- AX3, AX5, and J5R definitions are fixed
- IRS computation is a deterministic function of input expression values

No disease labels were used in IRS construction.

**S7. Endothelial coupling analysis across inflammatory resolution states**

S7.1 Purpose and scope

This section defines how coordination between endothelial programs and other cell-specific transcriptional states was quantified as a function of inflammatory resolution. The objective of this analysis was to determine whether endothelial coordination behaves differently across resolved and unresolved states in healthy, endometriosis, and autoimmune cohorts, after controlling for tissue composition and menstrual timing via the Inflammatory Resolution Score (IRS).

All analyses in this section were conducted at the kit level and use the canonical IRS defined in Supplementary Methods S6.

S7.2 Input data and unit of analysis

Inputs to this analysis were:

- Kit-level cell-specific switch matrix, constructed as described in Supplementary Methods S4
- Kit-level IRS values, defined in Supplementary Methods S6
- Cohort labels, including Healthy, Endometriosis, and Autoimmune

Only kits meeting all QC criteria defined in Supplementary Methods S1 were included.

Analyses were performed independently within each cohort to preserve disease-specific structure.

S7.3 Definition of endothelial activity score

Endothelial activity was summarized using a single composite score per kit, referred to as the ENDO score.

Construction of the ENDO score proceeded as follows:

1. All switches annotated as *Endothelial* in the switch ontology table were identified
2. For each endothelial switch, kit-level switch scores were z-scored across all kits within the cohort
3. The ENDO score for a given kit was defined as the mean z-score across all endothelial switches

Formally, for kit *k*:

$$\text{ENDO}_{k}=\frac{1}{N_{\text{endo}}}\sum_{i=1}^{N_{\text{endo}}} z(\text{SwitchScore}_{i,k})$$

This aggregation reduces noise from individual endothelial programs while preserving coordinated endothelial behavior.

S7.4 Stratification into resolved and unresolved states

Resolution status was defined using IRS thresholds applied *post hoc* and independently of endothelial data.

For each cohort, kits were classified as:

- Unresolved: IRS below the cohort-specific threshold defined as the median
- Resolved: IRS above the cohort-specific threshold defined as the median

Threshold values and cohort-specific distributions are reported in the corresponding Results sections. No thresholds were optimized or tuned within this analysis.

S7.5 Quantification of endothelial coupling

For each cohort and each resolution state separately, endothelial coupling to all other cell-specific switches was quantified using Pearson correlation.

For a given switch *j*:

- $r_{j}^{\text{unres}}$: Pearson correlation between ENDO score and switch *j* across unresolved kits
- $r_{j}^{\text{res}}$: Pearson correlation between ENDO score and switch *j* across resolved kits

Correlations were computed using raw kit-level switch scores without additional scaling beyond the z-scoring applied to endothelial switches during ENDO score construction.

Pearson correlation was used in endothelial coupling analyses to preserve linear effect size interpretation, whereas Spearman correlation was used in network rewiring analyses to ensure robustness to non-normal expression distributions.

S7.6 Differential coupling statistic (Δr)

For each switch *j*, differential endothelial coupling was defined as:

$$\Delta r_{j}=r_{j}^{\text{unres}}-r_{j}^{\text{res}}$$

Positive Δr values indicate stronger endothelial coupling in unresolved states, while negative Δr values indicate stronger coupling in resolved states.

S7.7 Statistical testing of differential coupling

Differences between correlation coefficients were assessed using Fisher’s z-transformation.

For each switch *j*, the null hypothesis tested was:

$$H_{0}:r_{j}^{\text{unres}}=r_{j}^{\text{res}}$$

Two-sided p-values were computed accounting for the number of kits contributing to each correlation estimate.

Multiple testing correction was performed using the Benjamini–Hochberg false discovery rate (FDR) across all 489 switches within each cohort. Switches with FDR < 0.1 were considered significantly differentially coupled.

S7.8 Organization and annotation of significant switches

Significant switches were annotated using the fixed switch ontology defined in Supplementary Methods S4, including:

- Subgroup (Epithelial, Stroma, Myeloid, Lymphoid, Endothelial, Hematopoietic Stem Cell)
- CellType (xCell-defined)

No aggregation or collapsing of switches was performed prior to statistical testing. Aggregation by Subgroup was used only for visualization and interpretation.

S7.9 Interpretation and downstream use

This analysis identifies resolution-dependent changes in endothelial coordination, distinguishing:

- Stable endothelial behavior in healthy cycles
- Epithelial- and stromal-biased endothelial engagement in endometriosis
- Immune- and antigen-presenting–biased endothelial hyper-coupling in autoimmune disease

Results from this section are reported in the main manuscript and Supplementary Results and are used to interpret disease-specific failure modes of inflammatory resolution.

S7.10 What is fixed versus learned

For reproducibility:

- Endothelial switch annotations are fixed
- ENDO score construction is deterministic
- IRS thresholds are applied post hoc and not optimized here
- Correlation method (Pearson) and testing framework (Fisher z + BH-FDR) are fixed

No disease labels were used to define ENDO score or switch membership.

### **S8. Network rewiring analyses of unresolved inflammatory states**

#### S8.1 Purpose and scope

This section defines the network-based analyses used to characterize transcriptional reorganization in unresolved inflammatory states across disease cohorts. Two complementary rewiring frameworks were employed:

1. **ΔZ-based network rewiring**, which quantifies changes in gene–gene correlation structure between resolved and unresolved states and identifies rewired hubs.
2. **Δρ-based pathway-level rewiring**, which quantifies directional changes in correlation strength between key hubs and pathway genes.

Together, these analyses distinguish disease-specific failure modes of inflammatory resolution beyond gene-level differential expression. All analyses in this section were conducted at the **kit level** and restricted to **menstrual cycle day 3**, unless otherwise specified, to minimize temporal heterogeneity.

#### S8.2 Input data and preprocessing

Inputs to all network analyses were:

- **Kit-level JaneScore-normalized gene expression matrix** (Supplementary Methods S2)
- **Kit-level IRS values** (Supplementary Methods S6)
- **Cohort labels** (Healthy, Endometriosis, Autoimmune)

For kits with multiple samples, gene expression values were aggregated by taking the mean across samples per kit prior to analysis. Only kits meeting all QC criteria defined in Supplementary Methods S1 were included.

#### S8.3 Definition of resolved and unresolved states

Resolution status was defined using IRS thresholds applied post hoc and independently of network structure.

Within each cohort, kits were classified as:

- **Unresolved**: IRS below the cohort-specific threshold
- **Resolved**: IRS above the cohort-specific threshold

Threshold values were fixed based on IRS distributions reported in the Results and were not optimized within network analyses.

### Part I. ΔZ-based network rewiring and hub identification

#### S8.4 Construction of correlation matrices

For each cohort independently, gene–gene correlation matrices were constructed separately for resolved and unresolved kits.

- Gene expression values were JaneScore-normalized and log₂-scaled
- Spearman correlation coefficients were computed pairwise across genes
- Analyses were restricted to the **top 200 genes** selected based on differential expression signal magnitude to focus on biologically responsive features

This yielded two correlation matrices per cohort:

- $R^{\text{res}}$(resolved)
- $R^{\text{unres}}$(unresolved)

#### S8.5 Fisher z-transformation and ΔZ computation

To stabilize variance across correlation estimates, all correlation coefficients were transformed using Fisher’s z transformation:

$$Z=\frac{1}{2}\ln(\frac{1+r}{1-r})$$

For each unordered gene pair (i, j), network rewiring was quantified as:

$$\Delta Z_{ij}=Z_{ij}^{\text{unres}}-Z_{ij}^{\text{res}}$$

Absolute ΔZ values quantify the magnitude of correlation restructuring independent of direction.

#### S8.6 Permutation testing and FDR control

To assess statistical significance of ΔZ values, a permutation-based null distribution was constructed:

1. Resolution labels (resolved vs unresolved) were randomly permuted within cohort
2. Correlation matrices and ΔZ values were recomputed
3. This process was repeated **500 times** to generate a null distribution for each gene pair

Empirical p-values were computed by comparing observed |ΔZ| values to the permutation null. Multiple testing correction was performed using **Benjamini–Hochberg FDR** across all tested edges within each cohort.

Edges with FDR ≤ 0.1 were considered significantly rewired.

#### S8.7 Identification of rewired hubs

To summarize rewiring at the gene level, node-wise rewiring scores were computed.

For each gene g, mean absolute ΔZ across all incident significant edges was calculated:

$$\text{HubScore}_{g}=\frac{1}{N_{g}}\sum_{j} \mid\Delta Z_{g,j}\mid$$

Genes were ranked by HubScore, and the **top 10 genes per cohort** were designated as rewired hubs.

Hub identities were not constrained to overlap across cohorts.

#### S8.8 Pathway gene sets (HALLMARK)

To interpret network rewiring at the level of biologically coherent processes, we used curated gene sets from the **Molecular Signatures Database (MSigDB) HALLMARK collection**. The HALLMARK gene sets distill overlapping pathways into compact, non-redundant representations of core biological programs and are therefore well suited for correlation-based network analyses. All pathway analyses in this section used **HALLMARK gene sets exclusively**. No GO, Reactome, or custom gene sets were used.

The following HALLMARK pathways were evaluated based on their relevance to inflammatory resolution, tissue remodeling, immune activation, and vascular biology:

- HALLMARK_EPITHELIAL_MESENCHYMAL_TRANSITION
- HALLMARK_ANGIOGENESIS
- HALLMARK_INTERFERON_ALPHA_RESPONSE
- HALLMARK_INTERFERON_GAMMA_RESPONSE
- HALLMARK_INFLAMMATORY_RESPONSE
- HALLMARK_TNFA_SIGNALING_VIA_NFKB
- HALLMARK_IL6_JAK_STAT3_SIGNALING
- HALLMARK_COMPLEMENT
- HALLMARK_APOPTOSIS
- HALLMARK_HYPOXIA
- HALLMARK_OXIDATIVE_PHOSPHORYLATION
- HALLMARK_G2M_CHECKPOINT

Each HALLMARK gene set was intersected with the kit-level JaneScore-normalized expression matrix prior to analysis to ensure all pathway members reflected measured transcriptome features.

#### S8.9 Directional correlation rewiring (Δρ)

For each cohort and each resolved–unresolved contrast, **directional network rewiring** was quantified between rewired hub genes (identified in S8.7) and genes belonging to each HALLMARK pathway.

For a given hub gene h and pathway gene g, Spearman correlations were computed separately in resolved and unresolved kits:

- $\rho_{h,g}^{\text{res}}$: correlation in resolved kits
- $\rho_{h,g}^{\text{unres}}$: correlation in unresolved kits

Directional rewiring was defined as:

$$\Delta\rho_{h,g}=\rho_{h,g}^{\text{unres}}-\rho_{h,g}^{\text{res}}$$

Positive Δρ values indicate stronger coordination in unresolved states, whereas negative Δρ values indicate stronger coordination in resolved states.

Pearson correlation was used in endothelial coupling analyses to preserve linear effect size interpretation, whereas Spearman correlation was used in network rewiring analyses to ensure robustness to non-normal expression distributions.

#### S8.10 Aggregation of Δρ to pathway-level metrics

For each HALLMARK pathway and cohort, Δρ values were summarized using the following predefined metrics:

1. **Engagement fraction (|Δρ| ≥ 0.5)**
   Fraction of pathway genes exhibiting moderate or greater rewiring.
2. **Strong engagement fraction (|Δρ| ≥ 0.7)**
   Fraction of pathway genes exhibiting high-amplitude rewiring.
3. **Mean and maximum rewiring amplitude**
   For each pathway gene, the maximum |Δρ| across all rewired hubs was computed. Pathway-level summaries were calculated as the mean and maximum of these values.

Genes with maximum |Δρ| < 0.3 across all hubs were classified as **pathway dropouts**, reflecting functional disengagement in that cohort.

These thresholds were fixed a priori and applied uniformly across all cohorts and pathways.

#### S8.11 Interpretation and downstream use

ΔZ-based analyses characterize **global network restructuring and hub architecture**, while Δρ-based analyses reveal **directionality and pathway-specific engagement**.

Together, these frameworks distinguish:

- Global, high-amplitude immune–endothelial rewiring in autoimmune disease
- Selective stromal and epithelial rewiring in endometriosis
- Transient, high-amplitude but sparse rewiring in healthy unresolved states

These results are reported in the main manuscript and Supplementary Results and provide mechanistic context for disease-specific failure modes of inflammatory resolution.

#### S8.12 What is fixed versus learned

For reproducibility:

- Gene universe (top 200 genes) is fixed per cohort
- Permutation count (500) and FDR threshold (0.1) are fixed
- Hub count (top 10) is fixed
- Δρ thresholds (0.3, 0.5) are fixed and biologically motivated
- Pathway gene sets are fixed

No tuning was performed to optimize disease separation.

### **S9. Microbial metatranscriptomic profiling and RNA–DNA comparison**

### S9.1 Purpose and scope

This section defines how microbial transcriptional activity was quantified from menstrual effluent RNA-seq data and how these profiles were compared to microbial presence measured by 16S rRNA gene sequencing in paired samples. The goal of these analyses was to distinguish microbial **activity** from microbial **abundance** and to assess whether observed microbial signals were structured, reproducible, and distinct from technical contamination.

All analyses in this section were descriptive and comparative. Microbial features were not used to construct IRS and were incorporated into diagnostic modeling only as explicitly described in Supplementary Methods S10.

#### S9.2 Input data and unit of analysis

Inputs to microbial analyses included:

- **Unmapped RNA-seq reads** from STAR alignments (Supplementary Methods S1–S2)
- **Paired 16S rRNA gene sequencing data**, available for a subset of samples
- **Negative controls**, including blank tampons and buffer-only controls processed alongside biological samples

Unless otherwise specified, analyses were performed at the **sample level** to preserve paired structure between RNA and DNA measurements. Only samples passing all RNA-seq QC thresholds defined in Supplementary Methods S1 were included.

#### S9.3 Extraction of microbial reads from RNA-seq data

Following alignment of RNA-seq reads to the human genome (hg38) using STAR, reads that did not align to the human reference were extracted from the resulting BAM files. These unmapped reads were treated as candidate microbial transcripts and were used as input for taxonomic classification.

#### S9.4 Taxonomic classification using Kraken2

Unmapped reads were classified taxonomically using **Kraken2**, a k-mer–based taxonomic classification system.

- Classification was performed against a comprehensive microbial reference database
- Reads were assigned to the lowest possible taxonomic rank supported by the database
- Only bacterial taxa were retained for downstream analysis

Taxonomic assignments were aggregated to the **genus level** for all analyses to maximize robustness and comparability across platforms.

#### S9.5 Normalization and transformation of microbial RNA counts

For each sample, genus-level microbial RNA counts were normalized as **counts per million (CPM)** using the total number of microbial reads detected in that sample as the library size denominator.

Normalized values were then log-transformed as:

$$\log_{2}(\text{CPM}+1)$$

This transformation stabilizes variance while retaining interpretability for low-abundance taxa.

#### S9.6 Processing of 16S rRNA gene sequencing data

Paired 16S rRNA gene sequencing data were processed separately using standard amplicon pipelines (described in Supplementary Results). Taxonomic assignments were aggregated to the **genus level**.

For direct comparison with metatranscriptomic data:

- Genus-level 16S counts and metatranscriptomic counts were converted to **relative abundance** within each sample
- Only genera detected in both 16S and metatranscriptomic datasets were retained

No attempt was made to equate absolute abundance between platforms; comparisons were performed on a compositional basis.

#### S9.7 Identification of paired RNA–DNA samples

Paired analyses were restricted to samples for which both the following were available from the same specimen.

1. Metatranscriptomic RNA-seq data, and
2. 16S rRNA gene sequencing data

Sample identifiers were harmonized across datasets using patient ID, kit ID, and collection metadata. Only unambiguous one-to-one matches were retained.

#### S9.8 Comparison of microbial presence and activity

##### S9.8.1 Bray–Curtis dissimilarity

To quantify global differences between microbial presence (16S) and microbial activity (RNA), **Bray–Curtis dissimilarity** was computed between paired genus-level profiles for each sample.

Bray–Curtis distances were used because they:

- are sensitive to compositional differences,
- do not assume normality,
- emphasize relative rather than absolute abundance.

Within-patient and between-patient distances were compared descriptively to assess individuality and stability of microbial profiles.

##### S9.8.2 Genus-level activity–abundance difference (Δ)

To identify taxa exhibiting disproportionate transcriptional activity relative to DNA abundance, a genus-level difference statistic was computed:

$$\Delta_{g}=\log_{2}(\text{RNA}_{g}+1)-\log_{2}(\text{DNA}_{g}+1)$$

where RNA${}_{g}$ denotes log₂(CPM+1) from metatranscriptomic data and DNA${}_{g}$ denotes relative abundance from 16S data.

- Positive Δ indicates transcriptional enrichment relative to abundance
- Negative Δ indicates structural abundance with comparatively low transcriptional activity

Δ values were summarized across samples and tissues to identify consistent directional patterns.

#### S9.9 Statistical testing and multiple-testing correction

For genus-level Δ analyses:

- One-sample tests were used to assess whether mean Δ differed from zero within a tissue compartment
- Tests were performed independently for menstrual effluent and cervicovaginal samples where applicable
- P-values were adjusted using **Benjamini–Hochberg false discovery rate (FDR)** correction within each tissue compartment

Because paired sample sizes were modest, non-parametric tests were favored where distributional assumptions were violated.

#### S9.10 Contamination controls and filtering

To assess potential reagent or device contamination:

- Blank tampons and buffer-only controls were processed through RNA extraction, library preparation, sequencing, and Kraken2 classification
- Taxa detected consistently in negative controls or at extremely low abundance across biological samples were flagged as likely contaminants

For downstream analyses:

- Low-prevalence taxa were excluded as described in Supplementary Results
- Remaining taxa showed structured, sample-specific, and tissue-specific patterns inconsistent with random contamination

No taxa were removed based on disease status or outcome.

#### S9.11 Interpretation and limitations

These analyses distinguish microbial **transcriptional activity** from microbial **presence** and demonstrate that RNA-based microbial profiles are structured, individualized, and partially decoupled from DNA-based abundance.

Because metatranscriptomic profiling reflects activity at the time of sampling, results should not be interpreted as absolute microbial load or infection status. Analyses were not powered to infer causality or temporal dynamics beyond the sampling window.

### **S10. Endometriosis diagnostic classifier development and evaluation**

#### S10.1 Purpose and intended use

This section defines the development, training, and evaluation of a non-invasive endometriosis diagnostic classifier derived from menstrual effluent molecular profiles. The classifier was designed for **cycle-aware, tissue-aware discrimination** between endometriosis cases and symptomatic or infertile controls within a restricted biological window. It was not intended as a population screening tool and was not trained or evaluated outside its stated intended use.

All analyses in this section were conducted independently of IRS construction and network analyses, except where explicitly stated.

#### S10.2 Input data and unit of analysis

Classifier discovery and training occurred at the sample level, where some kits contained multiple replicates. This aided in modeling technical variation in the SVM model. Classifier validation used **kit-level data**, with one kit per patient used for model testing and evaluation.

Inputs included:

- **Kit-level JaneScore-normalized gene expression values** (Supplementary Methods S2)
- **Kit-level microbial features** derived from metatranscriptomic profiling (Supplementary Methods S9)
- **Tissue composition metrics** AX3 and AX5 (Supplementary Methods S3)
- **Bleeding phenotype**, derived from participant-reported metadata

Only kits meeting all QC criteria defined in Supplementary Methods S1 were eligible.

#### S10.3 Cohort construction and segregation

##### S10.3.1 Training cohort

The training cohort consisted of **46 patients** contributing **136 kits** collected on menstrual cycle days 2–3. These included laparoscopically confirmed endometriosis cases and symptomatic or infertile controls.

Multiple kits from the same patient were permitted in the training set to improve feature discovery; however, **all model evaluation steps were performed at the patient level**, and no patient appeared in both training and validation cohorts.

##### S10.3.2 Independent validation cohort

Independent validation was performed on **139 patients**, with **one kit per patient**, collected on cycle days 2–3. None of these patients contributed data to the training cohort.

Subgroup analyses were conducted within this validation cohort for:

- symptomatic patients (fibroids, PCOS, adenomyosis, infertility, Chronic pelvic pain and/or autoimmune disease)
- infertility patients,

#### S10.4 Feature selection and constraints

Candidate features were drawn from four predefined categories:

1. **Host gene expression features**
2. **Microbial transcriptional features**
3. **Tissue composition metrics** (AX3, AX5)
4. **Bleeding phenotype** (categorical)

Feature selection was guided by the following constraints:

- Features showing strong dependence on cycle day were excluded
- Features strongly correlated with tissue composition (AX3, AX5, or J5R) were excluded
- Only features demonstrating consistent signal across training samples were retained
- Low expressor genes were excluded

The final classifier used:

- **9 host gene features**,
- **8 microbial features**,
- AX3 and AX5, and
- bleeding phenotype.

The feature list was fixed prior to validation and was not modified thereafter.

#### S10.5 Model specification

A **linear support vector machine (SVM)** classifier was used.

- Kernel: linear
- Regularization parameter: fixed (C = 1)
- Implementation: scikit-learn

No nonlinear kernels or feature interactions were used, prioritizing interpretability and stability.

#### S10.6 Training procedure and leakage prevention

Model training was performed using the training cohort only.

To prevent information leakage:

- Patient identifiers were used to ensure that no patient contributed samples to both training and validation
- Feature selection was completed prior to validation
- No tuning or refitting was performed using validation data

Classifier decision thresholds were derived from training data only.

#### S10.7 Evaluation metrics

Classifier performance was evaluated using:

- **Area under the ROC curve (AUC)**
- **Matthews correlation coefficient (MCC)**
- **Sensitivity and specificity**

All metrics were computed at the **patient level** in the validation cohort.

#### S10.8 Nested cross-validation

To assess generalizability and overfitting risk, **nested cross-validation** was performed post hoc on the combined dataset of **186 patients**.

- The outer loop evaluated performance
- The inner loop selected hyperparameters (held fixed thereafter)

Nested cross-validation was performed strictly for performance estimation and **did not inform model training or feature selection**.

#### S10.9 Interpretation of classifier outputs

The classifier outputs a continuous decision score reflecting alignment with the endometriosis molecular signature.

- Higher scores indicate greater consistency with the endometriosis profile
- Scores do not represent probability of disease
- Thresholds were applied only for evaluation and reporting

Classifier outputs were interpreted in the context of cycle timing, tissue composition, and intended use.

#### S10.10 Limitations and scope

The classifier was trained and validated exclusively on samples collected on cycle days 2–3 and is not expected to generalize outside this window. The classifier performance shows similar performance on day 4 menstrual samples and reduced performance on day 1. Performance in individuals on hormonal suppression or outside the intended symptomatic/infertility context were assessed but not included in the final performance characteristics.

The classifier is not intended for population screening and should be interpreted in conjunction with clinical evaluation.

#### S10.11 What is fixed versus learned

For reproducibility:

- Feature sets are fixed
- Model type and parameters are fixed
- Training and validation cohorts are strictly segregated
- No post-validation tuning was performed

All classifier behavior is a deterministic function of the specified inputs.
