## Supplemental Results for "Quantifying inflammatory resolution in human menstruation reveals disease-specific failure modes and enables a non-invasive diagnostic for endometriosis"

**SUPPLEMENTAL RESULTS I**

**Platform performance, RNA stability, and technical robustness**

To evaluate the stability and reproducibility of tampon-collected menstrual effluence under decentralized, real-world conditions, we quantified RNA integrity, yield, sequencing quality, and technical reproducibility across a large, heterogeneous dataset. Analyses in this section are intended to establish that observed biological structure in the main manuscript is not driven by pre-analytical degradation, batch effects, or contamination.

**RNA integrity is stable across ambient transit and dominated by biological factors**

RNA integrity was assessed in **1,135 RNA-seq libraries derived from 601 tampon collections** using the DegNorm-derived **DI₍25₎** metric, which quantifies 5′–3′ transcript degradation. DI₍25₎ showed no significant association with time in stabilization buffer under real-world shipping conditions (ANOVA **p = 0.107**), indicating that ambient transit time does not measurably impact RNA integrity (Supplementary Figure 1a). In contrast, DI₍25₎ varied systematically with menstrual flow rate, with higher flow associated with lower degradation values (Kruskal–Wallis **p < 1 × 10⁻⁴**), consistent with biological shedding dynamics rather than technical handling driving integrity differences (Supplementary Figure 1b).

Seasonal and geographic effects on RNA integrity were modest. DI₍25₎ varied slightly by collection season and U.S. region, with Midwest samples exhibiting a lower mean DI₍25₎ than other regions (**p = 0.023**, Cohen’s *f* = 0.127), but these effects were small relative to flow-associated variation and did not approach thresholds that would compromise downstream analyses (Supplementary Figure 1c–d).

**RNA yield and extraction reproducibility support scalable library preparation**

Total RNA yield was sufficient and consistent across the cohort to support ribosomal RNA–depleted library preparation without additional concentration steps. From a **600 µL aliquot**, **93%** of samples extracted using magnetic bead–based protocols yielded at least **500 ng** of total RNA, corresponding to the upper input requirement for the Zymo ribofree library preparation kit (Supplementary Figure 2a). Median RNA yield was **1.5 µg**, providing substantial margin for repeat library preparation or orthogonal assays.

Technical reproducibility was high at both the gene and module levels. Within-kit technical replicates showed a median gene-level Pearson correlation of **0.979**, while between-kit replicates from the same individual collected on the same day retained a median correlation of **0.957** (Supplementary Figure 2a). Higher-order biological features used in downstream analyses, including co-expression modules and cell-specific activity scores, exhibited within-kit coefficients of variation below **3%** (Supplementary Figure 2b), indicating that derived features are at least as stable as gene-level measurements.

**Technician-level reproducibility across independent library preparations**

To quantify operator-level variability in RNA integrity measurements, we assessed technician reproducibility using kits processed across independent sequencing preparations. The technician comparison dataset contained **49 libraries from 10 kits**, processed by **two technicians** with multiple libraries per kit (median **4** libraries per kit; range **4–9**).

Across these 10 kits, **DI₍25₎ reproducibility was high**, with a **median within-kit coefficient of variation (CV) of 0.149** (mean CV 0.180). Within-kit DI₍25₎ ranges were modest relative to between-kit differences; for example, kit-level DI₍25₎ spanned **0.065 to 0.175** across the dataset when summarized at the kit mean. When DI₍25₎ was summarized by technician within each kit, the **mean absolute difference in technician means (Technician 2 minus Technician 1)** was **0.0357 DI₍25₎ units** (median absolute difference **0.0300**; range **−0.0700 to +0.0533** across kits).

Together, these analyses show that technician-to-technician differences contribute a bounded, quantifiable component of DI₍25₎ variability, with a typical technician effect size on the order of **~0.03–0.04 DI₍25₎ units** at the kit level. This magnitude is small relative to the biologically structured DI₍25₎ shifts observed across flow and cohort strata in the full dataset, supporting the interpretation that the integrity trends reported in the main manuscript and supplement are not driven by operator effects.

**Sequencing quality metrics and failure rates remain low across time**

Sequencing quality metrics were uniformly high across libraries. More than **95%** of samples passed all predefined quality thresholds, including strandedness greater than **80%**, ribosomal RNA content below **3%**, male XY transcript contamination below **1 log₂ RPKM**, and at least **8 million** uniquely mapped reads (Supplementary Figure 2c). Failure rates declined over time as protocols were optimized, with improvements corresponding to adoption of DNase treatment, magnetic bead–based extraction, and replicate-aware sequencing workflows (Supplementary Figure 2d).

**Correlation of qPCR measurements between day 1 and day 14 buffer incubation**

To directly assess concordance of qPCR measurements across buffer incubation timepoints, we evaluated the correlation between cycle threshold–normalized values measured at **day 1** and **day 14** across all assayed targets. This analysis leverages paired measurements for each reaction rather than relying on per-target summary statistics.

Across **44 paired qPCR measurements spanning 11 assays**, day 1 and day 14 values were **strongly correlated**. Pearson correlation analysis yielded **r = 0.960** with **p = 9.26 × 10⁻²⁵**, corresponding to **R² = 0.921**. This indicates that more than **92% of the variance** in day 14 measurements is explained by day 1 measurements.

The tight linear relationship demonstrates that relative qPCR signal is preserved over two weeks of ambient buffer incubation, despite modest target-specific shifts in absolute Ct values. Importantly, this correlation-based assessment shows that samples maintain their **rank order and relative abundance structure** across timepoints, supporting the conclusion that buffer incubation does not introduce stochastic degradation or assay instability.

Together with RNA-seq–based DI₍25₎ stability metrics, these results provide orthogonal, quantitative evidence that nucleic acid integrity and relative abundance are maintained during extended ambient storage following buffer release.

**Effect of delayed stabilization in menstrual cup collections on RNA integrity**

To directly test whether post-collection delay prior to stabilization alters RNA integrity, we analyzed DI₍25₎ in menstrual cup samples incubated for **0 minutes versus 30 minutes** prior to stabilization. The incubation dataset contained **22 measurements at 0 minutes** and **8 measurements at 30 minutes**, and included **5 patients** with paired measurements across both timepoints.

Across all samples, DI₍25₎ increased at the 30-minute incubation timepoint relative to immediate stabilization. Median DI₍25₎ was **0.099** at 0 minutes versus **0.137** at 30 minutes (means **0.096** vs **0.135**). An unpaired comparison yielded **Mann–Whitney U = 140, p = 0.013**. In the paired patient subset (n = 5), the mean within-patient increase (30 minus 0) was **+0.0456 DI₍25₎ units** (median **+0.0431**, range **+0.0147 to +0.0792**). A paired t-test across the five paired patients yielded **t = 4.09, df = 4, p = 0.0149** (paired Wilcoxon p = 0.0625).

These results show that even a short post-collection delay prior to stabilization is associated with a measurable upward shift in DI₍25₎, providing quantitative support that **immediate stabilization is a key determinant of RNA integrity** in menstrual effluent and offering a mechanistic contrast to the stability observed after buffer release in the decentralized tampon workflow.

**Blank device testing and low-biomass controls confirm absence of contamination**

To assess potential contamination from collection materials or reagents, we performed quantitative PCR testing on blank tampons incubated in stabilization buffer and on buffer-only controls. Across **100 tested blank samples**, pathogen-specific assays yielded cycle threshold values of **34 or higher** following pre-amplification, with no lots showing signal exceeding established two-sigma thresholds (Supplementary Figure 5d). Sequencing of blank tampons confirmed low biomass, with fewer than **15,000 total bacterial reads** detected per sample and minimal host signal (Supplementary Figure 5e).

Together, these analyses demonstrate that tampon-collected menstrual effluence supports stable RNA integrity, sufficient yield, and high technical reproducibility under decentralized collection conditions. Biological factors, particularly menstrual flow dynamics, dominate residual variability, while technical effects and contamination remain low and well controlled. These findings establish a robust foundation for tissue-aware, cycle-resolved molecular analyses presented in the main manuscript.

**SUPPLEMENTAL RESULTS II**

**Normalization strategy and tissue-composition modeling preserve biological structure in heterogeneous menstrual effluence**

Accurate interpretation of menstrual effluence requires normalization strategies that stabilize expression values while preserving biologically meaningful variation arising from tissue composition and temporal progression. Because menstrual samples contain variable mixtures of uterine and cervicovaginal tissue that change across individuals and cycle days, global scaling approaches risk collapsing secondary biological structure or amplifying technical effects. This section evaluates normalization choices and demonstrates that tissue-aware modeling preserves interpretable biological gradients required for downstream analyses.

**Housekeeping gene–anchored normalization preserves secondary biological axes**

To stabilize expression values across compositionally heterogeneous samples, we implemented **JaneScore**, a reference-gene–anchored normalization strategy that scales each gene’s log-transformed expression relative to the mean expression of three housekeeping genes (**ACTB, GAPDH, and RAB7A**). These genes were selected for low variance across cervicovaginal and menstrual samples and minimal correlation with tissue composition axes.

We benchmarked JaneScore against raw counts, RPKM, and DESeq2 normalization using a **48-sample subset** spanning cervicovaginal, menstrual, and mixed tissue profiles. Principal component analysis (PCA) revealed that under all normalization methods, the first principal component (PC1) captured the dominant tissue composition axis. However, the behavior of the second principal component (PC2) differed markedly across methods.

Under JaneScore normalization, **AX3 and AX5 together explained 88% of PC1 variance (R² = 0.88; p < 1 × 10⁻¹⁶)** and **53% of PC2 variance (R² = 0.53; p = 3.3 × 10⁻⁸)**, indicating that both principal components retained interpretable biological structure (Supplementary Figure 3a). In contrast, under DESeq2 normalization, AX3 and AX5 explained **97% of PC1 variance** but only **10% of PC2 variance (R² = 0.10; p = 0.105)**, indicating that secondary structure was dominated by technical or participant-specific effects rather than tissue biology (Supplementary Figure 3a).

**Housekeeping gene behavior differs across normalization methods**

We further examined the behavior of housekeeping genes across normalization strategies. Under JaneScore normalization, housekeeping gene expression showed minimal correlation with AX3 and AX5, indicating effective decoupling from tissue composition effects. In contrast, under DESeq2 normalization, housekeeping genes exhibited altered correlations with tissue axes, reflecting preferential normalization toward a single dominant tissue source rather than preservation of mixed-tissue structure (Supplementary Figure 3d). These findings indicate that global normalization methods can inadvertently distort internal reference behavior in heterogeneous biospecimens.

**Comparison of menstrual effluent collected by tampon versus menstrual cup**

Because collection device can modulate the relative contribution of vaginal epithelium to menstrual effluent, we compared RNA-seq profiles obtained from **menstrual cup (n = 70)** versus **tampon (n = 68)** collections that passed sequencing quality filters. Comparisons were performed at the sequencing level using the same normalization and QC framework applied throughout the manuscript.

Device type was associated with large differences in keratinized epithelial signal. Expression of **KRT13** differed strongly between collection methods, with a one-way ANOVA yielding **F = 583.75** and **p = 4.69 × 10⁻⁵¹** across the 138 libraries. The corresponding effect size was extremely large (**η² = 0.811**, **Cohen’s f = 2.072**), indicating that device choice strongly perturbs keratinized epithelial contribution. In contrast, uterine-associated markers (including PAEP and ESR1, shown in the same dataset) remained detectable across both devices, supporting that both collection methods capture uterine-derived transcriptional signal, while differing most sharply in epithelial admixture.

These data demonstrate that tampon versus cup collection is not a neutral substitution at the level of epithelial content, and directly motivate explicit tissue-composition modeling as used in downstream analyses.

**AX3 and AX5 capture stable, biologically interpretable tissue axes**

Using JaneScore-normalized expression values, we derived co-expression modules representing dominant tissue axes from a **267-sample dataset** comprising cervicovaginal samples, menstrual effluence, and matched whole blood. Two modules, **AX3** and **AX5**, captured the primary axis of tissue variation and formed the basis for composition-aware modeling.

AX3 expression was highest in cervicovaginal samples, with a median value of **6.31**, approximately **2.3-fold higher** than in menstrual effluence (median **2.71**) and **2.2-fold higher** than in whole blood (median **2.89**). In contrast, AX5 expression was highest in menstrual effluence, with a median value of **5.39**, corresponding to **4.6-fold higher** expression than in cervicovaginal samples (median **1.18**) and **8.7-fold higher** expression than in whole blood (median **0.62**) (Supplementary Figure 4a).

Correlation with cell-type–specific gene signatures confirmed biological specificity. AX3 correlated strongly with epithelial programs, with median Spearman correlations exceeding **0.60**, while AX5 correlated with stromal, endothelial, dendritic cell, and myeloid signatures, with median correlations greater than **0.50** (Supplementary Figure 4b).

**AX3 and AX5 enable accurate tissue classification and continuous composition modeling**

Using only AX3 and AX5 scores, a k-nearest neighbors classifier (**k = 5**) achieved **97.99% accuracy** and a macro-averaged F1 score of **0.97** in classifying tissue origin in a held-out test set of **149 samples** (Supplementary Figure 4c). This performance demonstrates that the two modules capture dominant and discriminative tissue structure without reliance on extensive feature sets.

To summarize tissue contribution as a continuous variable, AX3 and AX5 were combined into **J5R**, a uterine enrichment metric derived from JaneScore-normalized values. Across the full cohort, J5R showed strong positive association with patient-reported flow descriptions and tampon flow rate (Global Kruskal–Wallis **p < 1 × 10⁻⁴**) and correlated with canonical uterine markers such as **PAEP** (Supplementary Figure 4d–f). These relationships validate J5R as a quantitative indicator of uterine contribution suitable for gating and regression-based adjustment.

**Robustness of tissue modeling across external reference datasets**

To assess whether tissue positioning was robust beyond the internal cohort, we compared JaneScore-normalized menstrual profiles to **GTEx reference tissues** using Spearman correlation on the **2,000 most variable shared genes**. Menstrual effluence exhibited intermediate similarity to reproductive tissues and blood, with correlations of **0.56–0.63** to cervix, vagina, uterus, and fallopian tube, and **0.70** to whole blood (Supplementary Figure 6). In contrast, cervicovaginal samples aligned more closely with GTEx cervix and vagina, and venous blood samples aligned strongly with GTEx whole blood (**r = 0.89**).

Principal component analysis confirmed that menstrual effluence occupies a distinct transcriptional space between reproductive epithelia and blood rather than collapsing onto either compartment (Supplementary Figure 6). This structure was preserved under JaneScore normalization, indicating that observed positioning reflects biological composition rather than normalization artifacts.

Together, these analyses demonstrate that housekeeping gene–anchored normalization preserves biologically interpretable structure in compositionally heterogeneous menstrual effluence. AX3, AX5, and J5R provide stable and quantitative representations of tissue composition, enabling explicit modeling of uterine and non-uterine contributions. These normalization and tissue-modeling choices form the foundation for inflammatory resolution analyses and diagnostic translation presented in the main manuscript.

**SUPPLEMENTAL RESULTS III**

**Construction, sensitivity, and robustness of the Inflammatory Resolution Score (IRS)**

The Inflammatory Resolution Score (IRS) was designed to quantify progression along the menstrual repair trajectory while explicitly accounting for tissue heterogeneity inherent to menstrual effluence. This section provides quantitative detail on IRS component construction, feature selection, residualization, and sensitivity analyses that support the main manuscript while remaining secondary to the biological interpretation presented there.

**Tissue-stratified IRS component construction**

To preserve biological specificity while enabling integration across heterogeneous samples, IRS was constructed from three tissue-aware components trained within biologically appropriate regimes: **AX5 IRS** (uterine-enriched), **AX3 IRS** (cervicovaginal-enriched), and **Core IRS** (tissue-agnostic). Samples were stratified using the J5R uterine enrichment metric, with uterine-enriched samples (J5R > 4.5) used for AX5 IRS training, cervicovaginal-enriched samples (J5R < 3.8) used for AX3 IRS training, and samples spanning the full composition range used for Core IRS training.

Feature assignment to IRS components was based on correlation with tissue axes. Genes and cell-specific switches with Spearman correlation ≥ 0.7 with AX5 were assigned to AX5 IRS, those with Spearman correlation ≥ 0.7 with AX3 were assigned to AX3 IRS, and features with absolute correlation < 0.3 to both axes were assigned to Core IRS. A minimum mean JaneScore-normalized expression threshold of 2 was required to exclude low-abundance features. Applying these criteria yielded **48 features for AX5 IRS**, **45 features for AX3 IRS**, and **69 features for Core IRS**.

Each component was trained using ridge regression to capture structured temporal variation while penalizing instability. AX5 IRS was trained on **523 uterine-enriched samples**, AX3 IRS on **184 cervicovaginal-enriched samples**, and Core IRS on **331 samples** spanning cycle days 1–3. Nested cross-validation was used to select regularization parameters, yielding penalty values of **46.42** for AX5 IRS and AX3 IRS, and **10.0** for Core IRS. In held-out samples, AX5 IRS achieved **R² = 0.36**, AX3 IRS achieved **R² = 0.70**, and Core IRS achieved **R² = 0.70**, confirming that each component captured reproducible temporal structure within its intended tissue context (Supplementary Figure 6a).

**Integration of tissue-specific components into Mixed IRS**

To enable resolution tracking across samples with heterogeneous tissue composition, AX5 IRS, AX3 IRS, and Core IRS were combined into a single composite score (Mixed IRS). Prior to integration, each component score was standardized. Integration was performed using ridge regression trained on JaneScore-normalized data, yielding weights of **0.191 for AX5 IRS**, **0.135 for Core IRS**, and **0.038 for AX3 IRS**, with an intercept of **1.789** (Supplementary Figure 6b).

The weighting structure reflects biological generalizability rather than variance dominance. AX5 IRS contributed the largest weight, consistent with the central role of uterine stromal remodeling in menstrual resolution. Core IRS contributed a substantial secondary weight, capturing shared temporal programs conserved across tissues. AX3 IRS contributed a smaller weight, reflecting its importance within epithelial-enriched samples but reduced generalizability across the full cohort. No single component dominated the integrated score, and all three contributed to the final Mixed IRS.

**Residualization removes tissue composition effects**

Because uterine-enriched samples constitute a large fraction of the dataset, Mixed IRS initially retained dependence on tissue composition. To ensure that the final score reflected resolution timing rather than uterine abundance, Mixed IRS was residualized with respect to J5R and standardized to produce the final IRS.

Prior to residualization, Mixed IRS showed a strong association with uterine enrichment (**Spearman r = −0.53; p ≈ 5.0 × 10⁻⁸⁰**). After residualization, this association was effectively eliminated (**r = −0.03; p = 0.28**), demonstrating successful removal of composition effects (Supplementary Figure 6c). Correlations with AX3 and J5R were minimal following correction, while correlation with AX5 remained modest (**r = −0.26**), consistent with uterine repair biology contributing to resolution timing without dominating the score.

**Sensitivity to feature selection and model complexity**

To assess whether IRS behavior depended on specific feature thresholds or modeling choices, we performed sensitivity analyses varying correlation cutoffs, expression thresholds, and regularization parameters. Relaxing tissue-correlation thresholds increased feature counts substantially and resulted in IRS variants composed of **more than 1,000 features**, but these variants exhibited increased variance, reduced reproducibility across cohorts, and diminished alignment with cycle-day progression (Supplementary Figure 6d). Conversely, overly stringent thresholds reduced feature counts but attenuated temporal signal.

The selected feature criteria balanced biological specificity and generalizability, yielding a compact feature set that preserved monotonic behavior across cycle days while minimizing noise. Across sensitivity analyses, the direction and ordering of IRS values by cycle day remained stable, indicating that IRS behavior reflects robust biological signal rather than tuning artifacts.

**IRS behavior across tissue regimes and cohorts**

IRS increased reproducibly from cycle day 1 through cycle day 3 across the full cohort, independent of tissue composition. When analyses were restricted to uterine-enriched samples only, IRS retained monotonic progression with similar effect sizes, indicating that integration across tissue regimes did not introduce spurious structure. Likewise, when analyses were restricted to cervicovaginal-enriched samples, IRS behavior remained directionally consistent but with increased variance, reflecting reduced uterine contribution in those samples.

Across disease cohorts, IRS distributions showed broader variance and altered central tendency relative to healthy controls, consistent with delayed or failed resolution rather than misclassification of tissue composition. These patterns were stable across repeated subsampling and were not driven by individual participants contributing multiple samples.

These analyses demonstrate that IRS construction is biologically grounded, quantitatively robust, and resistant to overfitting. By separating tissue-specific resolution programs, integrating them conservatively, and explicitly removing composition effects, IRS captures temporal progression through menstrual repair rather than static inflammatory burden. Sensitivity analyses confirm that IRS behavior is stable across reasonable modeling choices, supporting its use as a generalizable resolution metric in the main manuscript.

**SUPPLEMENTAL RESULTS IV**

**Network rewiring architecture and microbial activity in menstrual effluence**

This section provides quantitative depth for transcriptional network rewiring and microbial activity analyses summarized in the main manuscript.

**Cycle day 3 network rewiring: hub and edge distributions**

To characterize the extent of transcriptional reorganization underlying resolved and unresolved states at cycle day 3, we quantified changes in gene–gene correlation structure within each clinical cohort. Resolution status was defined post hoc for each analysis using IRS thresholds chosen to facilitate within-cohort contrasts; IRS itself is a continuous, tissue-normalized measure and was not thresholded during construction. For each cohort, Fisher z–transformed correlation matrices were constructed using the top 200 differentially expressed genes, and differences between unresolved and resolved states were summarized using ΔZ, defined as the absolute change in correlation strength for each gene–gene edge.

At a false discovery rate threshold of **FDR < 0.1**, unresolved autoimmune samples exhibited the most extensive network reorganization, with **81 significantly rewired hub genes** and **3,637 altered edges** out of **19,900 possible gene–gene relationships**, corresponding to **18.3%** of the inferred interaction network (Supplementary Figure 7a–b). The mean absolute ΔZ in this cohort was **0.88**, indicating broad and high-magnitude reorganization of correlation structure.

Unresolved endometriosis samples displayed a more constrained rewiring profile, with **20 rewired hub genes** and **2,651 altered edges** (**13.3%** of the network) and a mean absolute ΔZ of **0.66** (Supplementary Figure 7a–b). Rewired hubs spanned epithelial, stromal, and oxidative stress–related programs, consistent with selective disruption of tissue-centric coordination rather than global immune amplification.

Healthy unresolved samples showed the smallest fraction of altered network structure, with **10 rewired hubs** and **1,986 altered edges** (**9.9%** of the network), but exhibited the largest mean absolute ΔZ (**1.18**), indicating high-amplitude but spatially restricted reorganization (Supplementary Figure 7a–b). These patterns support the interpretation that unresolved states in healthy individuals reflect transient, self-limited deviations rather than persistent failure modes.

Complete lists of rewired hubs, edge-level ΔZ values, and corresponding false discovery rates are provided in Supplementary Data Tables.

**Directional pathway rewiring (Δρ) across clinical cohorts**

To assess whether transcriptional rewiring favored unresolved or resolved states within biological pathways, we quantified directional rewiring using Δρ, defined as the difference in hub–gene correlation between unresolved and resolved samples (Δρ = ρ_unresolved − ρ_resolved). Positive Δρ values indicate stronger coordination in unresolved states, whereas negative values indicate stronger coordination in resolved states.

Directional rewiring was summarized across **53 HALLMARK pathway sets**. In autoimmune unresolved samples, immune and interferon-related pathways exhibited consistently positive Δρ values, with interferon alpha response (mean Δρ = **0.67**) and interferon gamma response (mean Δρ = **0.53**) showing unresolved-dominant coordination in more than **95%** of pathway genes (Supplementary Figure 7c–d). Additional unresolved-dominant pathways included IL6–STAT3 signaling, MYC targets, and DNA repair, indicating broad immune and proliferative persistence.

In endometriosis unresolved samples, directional rewiring was more selective. Epithelial–mesenchymal transition and angiogenesis pathways showed positive Δρ values (mean Δρ = **0.31** and **0.18**, respectively), while multiple metabolic and proliferative pathways, including oxidative phosphorylation and the G2M checkpoint, showed negative Δρ values, indicating stronger organization in resolved states (Supplementary Figure 7c–d). This bidirectional pattern reflects partial disengagement of repair programs rather than global persistence.

Healthy unresolved samples exhibited high-amplitude Δρ values across several pathways but maintained balanced directionality, with both unresolved- and resolved-dominant coordination observed depending on pathway. This pattern is consistent with transient delay in otherwise intact repair dynamics rather than sustained failure.

**Endothelial coupling statistics across the resolution trajectory**

To extend cycle day–specific findings across the full menstrual window, we quantified endothelial coupling to other cell-specific transcriptional programs using IRS-based stratification. Because IRS captures continuous progression along the menstrual repair trajectory, endothelial coupling analyses should be interpreted as differences across resolution states rather than discrete biological classes. Endothelial activity was correlated with each of **489 cell-specific switches**, and differences between low-IRS and high-IRS states were summarized as Δr values.

In autoimmune samples, **58 switches** exhibited significant differential endothelial coupling at **FDR < 0.1**, with a strong bias toward positive Δr values. Median Δr across significant switches was approximately **+0.45**, indicating stronger endothelial coupling in low-IRS states (Supplementary Figure 7e). These effects were most pronounced in antigen-presenting cell and myeloid programs, where endothelial correlations frequently exceeded **0.6** in low-IRS samples and collapsed toward **0.1** in high-IRS samples.

Endometriosis samples showed **179 switches** with significant differential endothelial coupling, predominantly with negative Δr values. Median Δr values were approximately **−0.27**, indicating increased endothelial coupling in higher IRS states. These effects were concentrated in epithelial and stromal programs, consistent with progressive tissue remodeling during attempted resolution.

Healthy samples exhibited minimal differential endothelial coupling across IRS strata. No switches met significance thresholds after multiple testing correction, and Δr distributions were tightly centered near zero, with median absolute Δr values below **0.05**, indicating stable endothelial coordination across the resolution trajectory.

**Microbial activity profiling and contamination controls**

To assess microbial contributions to menstrual effluence, we performed paired **16S rRNA gene sequencing** and **metatranscriptomic profiling** on **53 samples from 11 participants**, enabling direct comparison of microbial presence and transcriptional activity within the same specimens. Across samples, genus-level RNA and DNA profiles differed systematically, with a median Bray–Curtis dissimilarity of **0.135** between RNA- and DNA-derived profiles (paired Wilcoxon signed-rank test **p = 1.19 × 10⁻¹⁰**), indicating structured divergence between microbial abundance and activity (Supplementary Figure 9b).

Within-participant Bray–Curtis distances were consistently lower than between-participant distances for both 16S and metatranscriptomic profiles, demonstrating strong participant-specific microbial signatures across compartments and time points (Supplementary Figure 9a). RNA–DNA divergence patterns were consistent across participants, with *Lactobacillus* showing negative activity relative to abundance and bacterial vaginosis–associated genera such as *Gardnerella*, *Anaerococcus*, and *Finegoldia* exhibiting positive activity–abundance differences.

To exclude contamination as a source of microbial signal, we performed lot-release quantitative PCR testing on **100 blank tampons** and buffer-only controls. All pathogen-specific assays yielded cycle threshold values of **34 or greater**, and no lots exceeded established two-sigma thresholds (Supplementary Table S8). Sequencing of blank tampons yielded fewer than **15,000 bacterial reads per sample**, confirming low biomass and absence of systematic contamination (Supplementary Figure 5).

Menstrual effluence contains transcriptionally active microbial signatures that vary with menstrual day and may include clinically relevant pathogens. Menstrual samples show a time dependent dysbiosis in microbial transcription with an increase in BV causing bacteria on days 2 and 3 of menstruation. RNA transcriptional activity is distinct from the presence (relative abundance) of microbial DNA. We performed genus-level metatranscriptomic analysis using Kraken2 on RNA reads. We identified high transcriptional activity from several bacterial genera commonly associated with vaginal and uterine microbiota, including Prevotella, Gardnerella, Corynebacterium, Finegoldia, and Anaerococcus (Supplemental Figure 5e). These genera are often implicated in microbial dysbiosis and reproductive tract infections. Lactobacillus species – typically associated with a healthy reproductive tract – were found to inversely correlate with the abundance of other bacterial taxa. In samples where Lactobacillus activity was high, the activity of other bacteria was generally suppressed. However, many samples exhibited low Lactobacillus levels (sum of all Lactobacillus species below 50% in 746 out of 1,135 sequences). alongside elevated expression of potentially pathogenic or opportunistic species. Interestingly, total Lactobacillus transcriptional activity decreased significantly on menstrual days 2 and 3 (Supplemental Figure 5f), suggesting a temporal shift in microbial composition during menstruation. These findings present an opportunity to explore and understand typical microbial transitions during a single menstrual cycle and map how vaginal microbiome returns to equilibrium after a menstrual disruption.

These supplemental analyses provide quantitative detail for network rewiring and microbial activity patterns described in the main manuscript. Full hub and edge distributions, directional pathway rewiring statistics, endothelial coupling values, and microbial presence–activity comparisons support the interpretation that unresolved inflammatory states exhibit structured, disease-specific architectures. Microbial signals are reproducible, participant-specific, and well controlled for contamination, supporting their contextual inclusion in resolution-aware analyses without introducing spurious diagnostic drivers.

**Endometriosis classifier and IRS**

Although the classifier was trained independently of IRS, diagnostic scores aligned with resolution biology in endometriosis cases. Across samples from those with endometriosis, ENDO SVM scores increased with IRS, with a positive association (β = 0.235; p ≈ 1 × 10⁻⁴), whereas this relationship was attenuated in healthy and unhealthy control samples (Supplementary Figure 8f). This concordance indicates that the diagnostic model captures molecular features associated with delayed or failed resolution rather than generic inflammation or tissue abundance, and that alignment with IRS arises from shared underlying biology rather than circular construction.

**Post hoc nested cross-validation of the endometriosis classifier**

To further quantify generalizability and evaluate overfitting risk under a strict leakage-control framework, we performed a post hoc nested cross-validation analysis of the endometriosis classifier across the full cycle day 2–3 dataset. This nested analysis was performed **after** independent validation and did not inform feature selection or model specification.

Across all included samples (combined training and validation; **n = 185**), nested cross-validation yielded a **median AUC of 0.865** with an interquartile range of **0.849–0.883**. The narrow spread of AUC values across folds indicates that diagnostic performance is stable under repeated re-partitioning when hyperparameter selection is confined to inner folds and evaluation is restricted to held-out outer folds.

Together, these nested cross-validation results quantitatively support that observed discrimination is not an artifact of a single train/test split and is consistent with generalizable signal under fold-separated model tuning.
