## Supplemental Tables for "Quantifying inflammatory resolution in human menstruation reveals disease-specific failure modes and enables a non-invasive diagnostic for endometriosis"

**Patient summary tables by analysis**

#### In total, 1,718 libraries from 1,004 tampons collected from 342 patients were included in this study, spanning various analyses. Below are summary lists of samples, kits, and patients used for each main analysis in this paper. Patient and cycle level characteristics, including comorbidities are included where appropriate. A full list of all samples and metadata included in this paper can be found in the supplemental data package. Patients inclusion were determined by reproductive age window, and ability to wear a tampon

#### **Supplementary Table 1. 16S–metatranscriptomic paired analysis cohort**

This cohort was selected to enable direct comparison between microbial presence (16S rRNA gene sequencing) and microbial transcriptional activity (metatranscriptomics) within the same biological specimens. Inclusion was restricted to vaginal and menstrual samples with paired 16S and RNA-seq data passing quality control. Venous blood samples were excluded from this analysis. The cohort is intentionally small and focused, reflecting the requirement for paired assays rather than population-level representativeness. These samples are used exclusively for benchmarking microbial activity–presence relationships and are not included in resolution modeling or diagnostic analyses.

| **Metric** | **16s meta** |
| --- | --- |
| Total number of seq libraries | 53 |
| Number of unique patients | 11 |
| Number of unique kits | 53 |
| Number and percent of menstrual samples | 9 (17.0%) |
| number and percent of vaginal samples | 44 (83.0%) |

#### **Supplementary Table 2. AX3/AX5 tissue-composition validation cohort**

This dataset was assembled specifically for tissue-composition modeling and calibration. It includes cervicovaginal samples collected outside of menstruation, menstrual samples, and matched venous blood draws from the same individuals. This structure was required to learn cervicovaginal- and uterine-enriched transcriptional programs (AX3 and AX5), to define a continuous uterine enrichment metric (J5R), and to anchor menstrual effluence relative to external reference tissues. Whole-blood samples from this cohort served as the primary comparator for alignment with GTEx whole-blood data. These samples are not used for IRS construction or diagnostic modeling.

| **Metric** | **AX3.AX5** |
| --- | --- |
| Total number of seq libraries | 267 |
| Number of unique patients | 32 |
| Number of unique kits | 267 |
| Number and percent of menstrual samples | 48 (18.0%) |
| number and percent of vaginal samples | 188 (70.4%) |
| number and percent of whole blood samples | 31 (11.6%) |

#### **Supplementary Table 3. Platform validation cohort**

This cohort was used to assess technical robustness, RNA integrity, and biological reproducibility of tampon-collected menstrual effluence under decentralized, real-world collection conditions. Inclusion was restricted to menstrual samples collected on cycle days 1–3 that passed sequencing quality control, with bleeding phenotype, flow type, race, and ethnicity recorded. Hormonal contraceptive users were excluded from downstream resolution analyses to avoid confounding of inflammatory timing. This cohort establishes platform stability and biological variance bounds and underlies reproducibility analyses but is not used directly for IRS training or diagnostic modeling.

| **Metric** | **Platform Validation** |
| --- | --- |
| Total number of seq libraries | 1135 |
| Number of unique patients | 276 |
| Number of unique kits | 601 |
| Number and percent of menstrual samples | 601 (100.0%) |
| number and percent of vaginal samples | 0 (0.0%) |
| Number and percent of samples with J5R below 3.8 | 170 (28.3%) of 601 kits with J5R |
| Number and percent of samples with J5R above 4.5 | 225 (37.4%) of 601 kits with J5R |
| ***Cycle Day (kits)*** | |
| Cycle Day 1 | 172 (28.62%) |
| Cycle Day 2 | 284 (47.25%) |
| Cycle Day 3 | 145 (24.13%) |
| ***Bleeding phenotype (kits)*** | |
| Heavy | 264 (43.9%) |
| Medium | 264 (43.9%) |
| Light | 73 (12.1%) |
| Unknown | 0 (0.0%) |
| Other | 0 (0.0%) |
| ***Age group (patients)*** | |
| 18–24 | 29 (10.5%) |
| 25–34 | 137 (49.6%) |
| 35–44 | 97 (35.1%) |
| 45+ | 13 (4.7%) |
| Unknown | 0 (0.0%) |
| ***Birth control (patients)*** | |
| No birth control | 221 (80.1%) |
| Estrogen+Progestin | 32 (11.6%) |
| Progestin-only | 18 (6.5%) |
| Non-hormonal IUD | 4 (1.4%) |
| Hormonal (unspecified) | 1 (0.4%) |
| Other/unspecified | 0 (0.0%) |
| Unknown | 0 (0.0%) |

#### **Supplementary Table 4. IRS analysis cohort**

This cohort was used for construction and validation of the Inflammatory Resolution Score (IRS). Inclusion was restricted to menstrual samples collected on cycle days 1–3 from patients not using hormonal birth control and passing all sequencing quality thresholds. These restrictions were applied to define a normative menstrual resolution trajectory without pharmacologic suppression. This cohort supports IRS training, residualization, and sensitivity analyses, and serves as the source population for subsequent cycle day–specific and trajectory-based resolution analyses. For cycle day 3 resolved vs. persistent/unresolved states. Only cycle day 3 samples were analyzed from healthy, endometriosis confirmed, and autoimmune confirmed or highly suspected patients, yielding 113 cycle day 3 tampons from 91 individuals, including 24 healthy controls, 55 endometriosis cases, and 34 autoimmune cases. Using 486 tampons from 208 patients spanning cycle days 1–3, we examined how endothelial cell programs correlate with other cell-specific activity states as samples progress along the resolution trajectory. Patients who did not fall under the healthy, endometriosis confirmed, or autoimmune confirmed or highly suspected definitions were excluded from this analysis.

| **Metric** | **IRS** |
| --- | --- |
| Total number of seq libraries | 1154 |
| Number of unique kits | 562 |
| Number of unique patients | 255 |
| ***Cycle Day (kits)*** | |
| Cycle Day 1 | 167 (29.72%) |
| Cycle Day 2 | 264 (46.98%) |
| Cycle Day 3 | 131 (23.31%) |
| ***Age group (patients)*** | |
| 18–24 | 20 (7.8%) |
| 25–34 | 118 (46.3%) |
| 35–44 | 100 (39.2%) |
| 45+ | 15 (5.9%) |
| Unknown | 2 (0.8%) |
| ***Birth control (patients)*** | |
| No birth control | 247 (96.9%) |
| Estrogen+Progestin | 0 (0.0%) |
| Progestin-only | 0 (0.0%) |
| Non-hormonal IUD | 0 (0.0%) |
| Hormonal (unspecified) | 0 (0.0%) |
| Other/unspecified | 0 (0.0%) |
| Unknown | 8 (3.1%) |
| ***Bleeding phenotype (kits)*** | |
| Heavy | 237 (42.2%) |
| Medium | 241 (42.9%) |
| Light | 57 (10.1%) |
| Unknown | 27 (4.8%) |
| Other | 0 (0.0%) |

#### **Endometriosis Training and Validation**

#### Samples used for endometriosis diagnostic development were drawn from the broader IRS analysis cohort described above, enabling direct linkage between resolution biology and diagnostic modeling. However, diagnostic discovery, training, and validation applied **additional, prespecified restrictions** to ensure clinical specificity and generalizability. Endometriosis cases were defined using strict criteria, including laparoscopic confirmation, imaging-verified endometrioma, or incidental surgical findings. Controls included putatively healthy individuals as well as symptomatic controls presenting with infertility, chronic pelvic pain, heavy bleeding, or other gynecologic or inflammatory conditions that may clinically resemble endometriosis, in whom endometriosis was not found or not suspected. **Post-surgical tampons were excluded from diagnostic discovery, training, and validation** to avoid confounding by treatment-induced molecular changes; these samples were analyzed separately solely to assess pre- versus post-surgical shifts in classifier scores. For model development, patients were randomly assigned to discovery/training versus validation cohorts, with assignment deliberately weighted to **prioritize a larger independent validation cohort** over discovery sample size, reflecting a conservative design choice intended to maximize assessment of generalizability. Detailed sample counts and cohort composition for diagnostic analyses are provided in Supplementary Tables S5 and S6.

#### **Supplementary Table 5. SVM discovery/training cohort (Cycle days 1-3)**

This cohort was used exclusively for diagnostic feature discovery and model training. Endometriosis cases were defined by laparoscopic confirmation, imaging-verified endometrioma, or incidental surgical findings. Controls included putatively healthy individuals as well as symptomatic controls presenting with infertility, chronic pelvic pain, heavy bleeding, or related gynecologic or inflammatory conditions in whom endometriosis was not found or not suspected. Forty-six patients were randomly selected for this cohort, with preference given to maximizing the size of independent validation cohorts rather than discovery sample size, reflecting a conservative design prioritizing generalizability.

| **Metric** | **Total** | **Case** | **Control** |
| --- | --- | --- | --- |
| Total number of seq libraries | 220 | 121 | 99 |
| Number of unique kits | 136 | 65 | 71 |
| Number of unique patients | 46 | 25 | 21 |
| *Age group (patients)* | | | |
| 18–24 | 4 (8.7%) | 3 (12.0%) | 1 (4.8%) |
| 25–34 | 23 (50.0%) | 11 (44.0%) | 12 (57.1%) |
| 35–44 | 17 (37.0%) | 11 (44.0%) | 6 (28.6%) |
| 45+ | 2 (4.3%) | 0 (0.0%) | 2 (9.5%) |
| Unknown | 0 (0.0%) | 0 (0.0%) | 0 (0.0%) |
| *Birth control (patients)* | | | |
| No birth control | 41 (89.1%) | 21 (84.0%) | 20 (95.2%) |
| Estrogen+Progestin | 2 (4.3%) | 2 (8.0%) | 0 (0.0%) |
| Progestin-only | 0 (0.0%) | 0 (0.0%) | 0 (0.0%) |
| Non-hormonal IUD | 2 (4.3%) | 1 (4.0%) | 1 (4.8%) |
| Hormonal (unspecified) | 0 (0.0%) | 0 (0.0%) | 0 (0.0%) |
| Other/unspecified | 0 (0.0%) | 0 (0.0%) | 0 (0.0%) |
| Unknown | 1 (2.2%) | 1 (4.0%) | 0 (0.0%) |
| *Bleeding phenotype (kits)* | | | |
| Heavy | 39 (28.7%) | 38 (58.5%) | 1 (1.4%) |
| Medium | 75 (55.1%) | 15 (23.1%) | 60 (84.5%) |
| Light | 16 (11.8%) | 11 (16.9%) | 5 (7.0%) |
| Unknown | 6 (4.4%) | 1 (1.5%) | 5 (7.0%) |
| Other | 0 (0.0%) | 0 (0.0%) | 0 (0.0%) |
| Endometriosis (confirmed) | 25 (54.3%) | 25 (100.0%) | 0 (0.0%) |
| Adenomyosis (confirmed or suspected) | 12 (26.1%) | 9 (36.0%) | 3 (14.3%) |
| Fibroids (confirmed or suspected) | 5 (10.9%) | 2 (8.0%) | 3 (14.3%) |
| PCOS (confirmed or suspected) | 8 (17.4%) | 6 (24.0%) | 2 (9.5%) |
| Autoimmune disease (confirmed or suspected) | 11 (23.9%) | 10 (40.0%) | 1 (4.8%) |
| Thyroid disease | 3 (6.5%) | 3 (12.0%) | 0 (0.0%) |

#### **Supplementary Table 6A. SVM validation cohort (cycle days 2–3)**

This cohort represents the independent validation population for endometriosis classifier performance. Validation was performed at the patient level using one tampon per individual to simulate intended clinical deployment. The cohort includes all-comers as well as prespecified symptomatic and infertility subgroups, enabling evaluation under increasing clinical enrichment rather than healthy screening conditions. No samples from this cohort were used in feature discovery or model training. Performance metrics reported in the main manuscript derive from this cohort and its clinically defined subgroups.

| **Metric** | **Total** | **Case** | **Control** |
| --- | --- | --- | --- |
| Total number of seq libraries | 556 | 273 | 283 |
| Number of unique kits | 245 | 118 | 127 |
| Number of unique patients | 139 | 73 | 66 |
| ***Age group (patients)*** | | | |
| 18–24 | 15 (10.8%) | 7 (9.6%) | 8 (12.1%) |
| 25–34 | 62 (44.6%) | 29 (39.7%) | 33 (50.0%) |
| 35–44 | 55 (39.6%) | 35 (47.9%) | 20 (30.3%) |
| 45+ | 7 (5.0%) | 2 (2.7%) | 5 (7.6%) |
| Unknown | 0 (0.0%) | 0 (0.0%) | 0 (0.0%) |
| ***Birth control (patients)*** | | | |
| No birth control | 121 (87.1%) | 67 (91.8%) | 54 (81.8%) |
| Estrogen+Progestin | 10 (7.2%) | 3 (4.1%) | 7 (10.6%) |
| Progestin-only | 4 (2.9%) | 1 (1.4%) | 3 (4.5%) |
| Non-hormonal IUD | 2 (1.4%) | 1 (1.4%) | 1 (1.5%) |
| Hormonal (unspecified) | 0 (0.0%) | 0 (0.0%) | 0 (0.0%) |
| Other/unspecified | 0 (0.0%) | 0 (0.0%) | 0 (0.0%) |
| Unknown | 2 (1.4%) | 1 (1.4%) | 1 (1.5%) |
| ***Bleeding phenotype (kits)*** | | | |
| Heavy | 103 (42.0%) | 82 (69.5%) | 21 (16.5%) |
| Medium | 108 (44.1%) | 27 (22.9%) | 81 (63.8%) |
| Light | 26 (10.6%) | 2 (1.7%) | 24 (18.9%) |
| Unknown | 8 (3.3%) | 7 (5.9%) | 1 (0.8%) |
| Other | 0 (0.0%) | 0 (0.0%) | 0 (0.0%) |
| Endometriosis (confirmed) | 73 (52.5%) | 73 (100.0%) | 0 (0.0%) |
| Adenomyosis (confirmed or suspected) | 42 (30.2%) | 35 (47.9%) | 7 (10.6%) |
| Fibroids (confirmed or suspected) | 23 (16.5%) | 17 (23.3%) | 6 (9.1%) |
| PCOS (confirmed or suspected) | 29 (20.9%) | 17 (23.3%) | 12 (18.2%) |
| Autoimmune disease (confirmed or suspected) | 28 (20.1%) | 22 (30.1%) | 6 (9.1%) |
| Thyroid disease | 12 (8.6%) | 6 (8.2%) | 6 (9.1%) |

**Supplementary Table 6B. SVM validation Performance by subcohort (cycle days 2–3**)

Clinical cohorts are defined based on increasing alignment with diagnostic intent. “All-comers” includes healthy controls, symptomatic individuals with non-endometriosis gynecologic or inflammatory conditions, and endometriosis cases, reflecting the full biological heterogeneity of menstrual effluence. A broader “unhealthy” cohort includes individuals presenting with any condition that could reasonably prompt evaluation for endometriosis or directly impact menstruation, independent of classical symptom criteria. A “symptomatic” cohort is further defined by infertility or chronic pelvic pain (pain score ≥7/10), representing patients for whom endometriosis workup is clinically indicated. The infertility cohort constitutes the intended-use population for the diagnostic assay. These cohorts are not mutually exclusive, reflecting real-world clinical overlap.

| Cohort | Number of kits | Patients (median per draw) | Endometriosis cases | Controls | Kit level AUC | Patient level AUC | Patient level AUC 95% CI | Patient level MCC | Patient level MCC 95% CI |
| --- | --- | --- | --- | --- | --- | --- | --- | --- | --- |
| All comers | 219 | 139 | 73 | 66 | 0.83 | 0.812 | 0.79–0.83 | 0.554 | 0.51–0.58 |
| Unhealthy | 146 | 97 | 65 | 32 | 0.88 | 0.844 | 0.83–0.86 | 0.604 | 0.58–0.64 |
| Symptomatic | 123 | 78 | 55 | 23 | 0.92 | 0.893 | 0.88–0.90 | 0.648 | 0.62–0.67 |
| Infertility | 72 | 54 | 37 | 17 | 0.94 | 0.922 | 0.91–0.94 | 0.712 | 0.68–0.76 |

### **Analysis agnostic patient tables**

Exclusions applied per protocol (Internal dataset used for AX3/AX5 development not included in summary tables); final cohort: 308 patients, 732 kits, 1426 samples.

#### Table S7A. Race (patients)

| Race | n | % |
| --- | --- | --- |
| White | 227 | 73.7 |
| Asian | 25 | 8.1 |
| Black or African American | 23 | 7.5 |
| More than one Race | 21 | 6.8 |
| Missing | 8 | 2.6 |
| American Indian or Alaska Native | 2 | 0.6 |
| Some Other Race | 2 | 0.6 |

#### Table S7B. Ethnicity (patients)

| Ethnicity | n | % |
| --- | --- | --- |
| Non-Hispanic | 259 | 84.1 |
| Hispanic or Latino | 37 | 12.0 |
| Missing | 12 | 3.9 |

#### Table S7C. Age bucket (patients)

| Age bucket | n | % |
| --- | --- | --- |
| 18–24 | 32 | 10.4 |
| 25–29 | 64 | 20.8 |
| 30–34 | 82 | 26.6 |
| 35–39 | 72 | 23.4 |
| 40–44 | 39 | 12.7 |
| 45+ | 17 | 5.5 |
| Missing | 2 | 0.6 |

#### Table S7D. Bleeding phenotype (patients)

| Bleeding phenotype | n | % |
| --- | --- | --- |
| Medium | 134 | 43.5 |
| Heavy | 132 | 42.9 |
| Light | 31 | 10.1 |
| Missing | 11 | 3.6 |

Sampling intensity: mean kits per patient = 2.38 ± 2.16 (SD).

#### Table S8. Disease burden (patients)

| Diagnosis | Confirmed n | Confirmed % | Suspected n | Suspected % |
| --- | --- | --- | --- | --- |
| Endometriosis | 136 | 44.2 | 35 | 11.4 |
| Adenomyosis | 53 | 17.2 | 29 | 9.4 |
| Fibroids | 34 | 11.0 | 23 | 7.5 |
| PCOS | 32 | 10.4 | 27 | 8.8 |
| Autoimmune | 37 | 12.0 | 30 | 9.7 |
| Thyroid | 20 | 6.5 | 4.0 | 1.3 |

#### Table S9. CPP pain severity by diagnosis category

Confirmed endometriosis cases were confirmed by laparoscopic surgery or through imaging for endometriomas. Adenomyosis and fibroid cases were confirmed through imaging. PCOS cases were confirmed through clinical workups and clinical labs. Suspected cases were designated “suspected” if there was symptomatic evidence coupled with family history or history of disease, patient confirmation but no medical report to support the finding, or there was significant evidence from medical reports and patient reported data to merit a “suspected” designation. For adenomyosis, patients were also categorized as “suspected” in circumstances were patient informed medical history demonstrates probability of disease.

| Diagnosis / Status | n patients | N pain non-missing | CPP Pain mean | CPP Pain SD |
| --- | --- | --- | --- | --- |
| ('Endometriosis', 'Confirmed') | 136 | 112 | 7.08 | 2.03 |
| ('Endometriosis', 'Suspected') | 35 | 21 | 7.76 | 1.34 |
| ('Adenomyosis', 'Confirmed') | 53 | 46 | 7.46 | 1.77 |
| ('Adenomyosis', 'Suspected') | 29 | 23 | 6.22 | 2.04 |
| ('Fibroids', 'Confirmed') | 34 | 29 | 6.74 | 2.72 |
| ('Fibroids', 'Suspected') | 23 | 22 | 6.05 | 3.05 |
| ('PCOS', 'Confirmed') | 32 | 24 | 6.5 | 2.92 |
| ('PCOS', 'Suspected') | 27 | 27 | 5.87 | 2.67 |
| ('Autoimmune', 'Confirmed') | 37 | 29 | 6.03 | 2.63 |
| ('Autoimmune', 'Suspected') | 30 | 24 | 7.29 | 1.57 |
| ('Thyroid', 'Confirmed') | 20 | 0 | 5.04 | 2.9 |
| ('Thyroid', 'Suspected') | 4 | 4 | 4.25 | 3.3 |

#### Table S10. Autoimmune subtypes (patient)

Diagnoses were self-reported and classified as confirmed or suspected based on participant report of clinician diagnosis. Conditions with emerging or debated autoimmune etiology (e.g., POTS, hypermobile Ehlers–Danlos syndrome, fibromyalgia) were retained as immune-associated.

| Category | Diagnosis (standardized) | Diagnostic status | n (patients) |
| --- | --- | --- | --- |
| **Endocrine autoimmune** | Hashimoto’s thyroiditis | Confirmed | 15 |
|  | Graves’ disease | Confirmed | 4 |
|  | Addison’s disease | Confirmed | 2 |
|  | Type 1 diabetes mellitus | Confirmed | 2 |
| **Gastrointestinal autoimmune** | Celiac disease | Confirmed | 4 |
|  | Inflammatory bowel disease (Crohn’s disease) | Confirmed | 3 |
|  | Inflammatory bowel disease (ulcerative colitis) | Confirmed | 1 |
| **Systemic autoimmune / connective tissue disease** | Systemic lupus erythematosus | Confirmed | 6 |
|  | Mixed connective tissue disease | Confirmed | 2 |
|  | Rheumatoid arthritis | Confirmed | 1 |
|  | Ankylosing spondylitis | Confirmed | 1 |
|  | Sjögren’s syndrome | Confirmed | 1 |
| **Dermatologic autoimmune** | Psoriatic arthritis | Confirmed | 1 |
|  | Psoriasis | Confirmed | 1 |
|  | Alopecia areata | Confirmed | 1 |
|  | Wells syndrome (eosinophilic cellulitis) | Suspected | 1 |
| **Granulomatous / inflammatory breast disease** | Granulomatous mastitis | Confirmed (immune-mediated) | 1 |
| **Neurologic / immune-mediated** | Narcolepsy type 1 | Confirmed | 1 |
| **Autonomic / connective tissue (immune-associated)** | Postural orthostatic tachycardia syndrome (POTS) | Confirmed | 1 |
|  | Postural orthostatic tachycardia syndrome (POTS) | Suspected | 4 |
|  | Hypermobile Ehlers–Danlos syndrome | Confirmed | 1 |
|  | Hypermobile Ehlers–Danlos syndrome | Suspected | 2 |
|  | Orthostatic intolerance | Confirmed | 1 |
| **Chronic pain / immune-associated** | Fibromyalgia | Suspected | 2 |
| **Unspecified autoimmune / immune-mediated** | Autoimmune disease, unspecified (e.g., positive ANA, clinician suspicion without formal diagnosis) | Suspected | 9 |
| **Reproductive immune dysregulation** | Immune-mediated infertility / reproductive immunology | Suspected | 2 |

#### Table S11A. Birth control (kits)

| Birth control | n | % |
| --- | --- | --- |
| No birth control | 592 | 80.9 |
| Estrogen and Progestin pill | 60 | 8.2 |
| Progestin IUD | 17 | 2.3 |
| Progestin pill | 17 | 2.3 |
| Non-hormonal IUD | 16 | 2.2 |
| Missing | 10 | 1.4 |
| Estrogen and Progestin vaginal ring | 9 | 1.2 |
| Progestin implant | 4 | 0.5 |
| Yes, did not specify | 3 | 0.4 |
| Progestin injection | 3 | 0.4 |
| Estrogen and Progestin patch | 1 | 0.1 |

#### Table S11B. Flow type (kits)

| Flow type | n | % |
| --- | --- | --- |
| Heavy | 346 | 47.3 |
| Medium | 218 | 29.8 |
| Light | 140 | 19.1 |
| Not Recorded | 15 | 2.0 |
| Spotting | 13 | 1.8 |

#### Table S11C. Cycle day (kits)

| Cycle day | n | % |
| --- | --- | --- |
| 1 | 207 | 28.3 |
| 2 | 322 | 44.0 |
| 3 | 171 | 23.4 |
| 4 | 32 | 4.4 |

#### Table S11D. Sample-level QC metrics

| Metric | Mean | SD | N (non-missing) |
| --- | --- | --- | --- |
| Strandedness_Category_2 (0-1) | 0.92 | 0.032 | 1426.0 |
| AVG % ribosomal_RNA (0-100) | 0.257 | 0.408 | 1426.0 |
| DI_25 | 0.112 | 0.045 | 1426.0 |
| a333XY (male transcripts) | 0.014 | 0.122 | 1426.0 |
| AX3 | 6.224 | 1.31 | 1426.0 |
| AX5 | 5.736 | 1.254 | 1426.0 |
| j5r | 4.251 | 0.625 | 1426.0 |
